# The Global Epilepsy Needs Study (GENS): A mixed-methods, multi-country exploration of the unmet psychosocial and everyday needs of people with epilepsy

**DOI:** 10.1101/2025.09.10.25334585

**Authors:** Gus A Baker, Sandeep Kumar Bagga, Donna Walsh, Claire Nolan, Shivani Sharma, Charlotte Hooker, Pepa Gonzalez Parrao, Francesca Sofia, J Helen Cross, Ma. Marta Bertone, Ivana García, Adam Jallow Janneh, Alison Kukla, Elvira Vacas Montero, Ding Ding, Latica Friedrich, Lecio Figueira Pinto, Gagandeep Singh, Chahnez Charfi Triki, Leya Raj, Allan Reese, GENS Collaborators

## Abstract

**Objective:** While epilepsy research has largely focused on medical management and clinical outcomes, less attention has been given to the unmet psychosocial, and everyday needs of people with epilepsy (PWE), particularly in low- and middle-income countries. The Global Epilepsy Needs Study (GENS) aims to explore these needs, which are integral to quality of life, by capturing both shared and context-specific experiences.

**Methods:** GENS employed a patient-centered approach and mixed-methods design, integrating a cross-sectional survey and semi-structured interviews in 15 countries. The survey, available in 12 languages, captured experiences across 10 life domains (n=5296). Interviews, analysed thematically using a phenomenological approach and Colaizzi’s Method, explored lived experiences in depth (n=75). To ensure meaningful involvement and diverse representation, national patient associations, healthcare professionals, researchers and people with lived experience guided each stage of the research process, from study design to manuscript development.

**Results:** Quantitative and qualitative data were integrated using a joint display method. This analysis generated 5 Generalised Themes across all life domains: 1) Managing uncertainty and redefining daily life; 2) Living with risk, social exclusion, and misunderstanding; 3) Challenges in navigating inaccessible systems; 4) Consequences of inaccessible or inadequate information; and 5) Complex epilepsy needs demand more than standard approaches.

**Significance:** This first-of-its-kind global study offers a comprehensive picture of the psychosocial and everyday challenges faced by PWE. It establishes a critical evidence base for epilepsy organisations, highlights the need for healthcare systems to adopt holistic, multidisciplinary approaches, and calls on policymakers to invest in systemic reforms that safeguard dignity, inclusion, and life opportunities. Future research should explore the needs of underserved groups, including caregivers, individuals with complex epilepsy, women, and those in low-income or rural settings.

**Plain Language Summary:** This study looked at the everyday challenges faced by people with epilepsy in different parts of the world. It showed that many people struggle with fear, stigma, poor access to services, and a lack of clear information and support. Women, people in rural areas, and those in low-income settings often face the greatest challenges. The study calls for better education, more support for caregivers, and improvements across health, work, school, and transport systems. It also shows the need for more research to understand and respond to the real-life needs of people most impacted by epilepsy.

## 1. INTRODUCTION

Epilepsy is one of the most common neurological conditions, affecting over 50 million people worldwide, with nearly 80% of cases occurring in low- and middle-income countries.^1,2^ It is characterised not only by an enduring predisposition to generate epileptic seizures but also by its neurobiological, cognitive, psychological, and social consequences.^3^

To date, most research has focused on seizure control, medication adherence, and access to specialist care,^4^ whereas the psychosocial and everyday life needs of people with epilepsy (PWE) and their caregivers remain comparatively underexplored. Yet, needs related to autonomy, education, employment, interpersonal relationships, and community inclusion are integral to quality of life (QoL).^5–7^ Stigma, discrimination, and social exclusion remain widespread, compounding the psychological and emotional burden of epilepsy.^4,8^

In recognition of the wide-reaching impact of epilepsy, the World Health Organization (WHO), in partnership with the International League Against Epilepsy (ILAE) and the International Bureau for Epilepsy (IBE) - a non-profit umbrella organisation comprising 160 member organisations in over 100 countries worldwide working together to achieve a transformational social change for PWE - launched the Global Campaign Against Epilepsy (GCAE) in 1997. This initiative laid the foundation for sustained international advocacy, culminating in 2022 with the unanimous adoption of the Intersectoral Global Action Plan on Epilepsy and Other Neurological Disorders (IGAP) by all WHO Member States, marking a significant commitment to address, holistically, the needs of PWE.^9^

However, the wider needs and experiences of PWE remain underrepresented in research; impeding meaningful implementation of these global commitments. Most existing studies have been conducted in high-income countries,^6,10–14^ with limited representation from low- and middle-income regions,^15–17^ where the majority of the global epilepsy population resides. Moreover, much of the literature centres on clinician perspectives,^17–20^ or focuses on specific subgroups,^21,22^ limiting the reach and relevance of findings. Few studies have included both PWE and their caregivers,^15,23^ and fewer still have employed mixed-methods designs capable of capturing the complexity of lived experience across diverse settings.^24^ While several international surveys have addressed topics such as stigma, digital health, or QoL,^5,18,25^ none have comprehensively mapped the psychosocial and everyday needs of PWE at a global scale.

IBE launched the Global Epilepsy Needs Study (GENS) as a response to the pressing need for such research. This multi-county, mixed methods initiative aims to explore the needs of PWE; understanding the complexity of experiences and the variation in socio-economic and cultural contexts. GENS provides a unique opportunity to shape inclusive, patient-centred research, care and policy; informed by lived experience.

## 2. METHODS

### 2.1 Study design

The study employed a mixed-methods approach, focusing on the unmet needs of PWE across 15 countries. Eligible participants were adults (aged 18 or older) who identified as a PWE, or a caregiver responding on behalf of a PWE. Study participants were recruited through IBE Chapters (national patient associations affiliated with IBE) in each participating country.

This international collaboration engaged stakeholders across the epilepsy community, including IBE representatives, national Chapters, healthcare professionals (HCPs), researchers, and individuals with lived experience (PWE and their caregivers). A strong patient-centered approach guided the study. PWE and caregivers worked alongside the researchers to co-create methodology, shape data collection, validate results and author the manuscript.

The study consisted of 2 primary components: a cross-sectional survey to capture broad, quantifiable insights, and semi-structured interviews to explore lived experiences in greater depth. Figure 1 outlines the research process.

**Figure 1:**
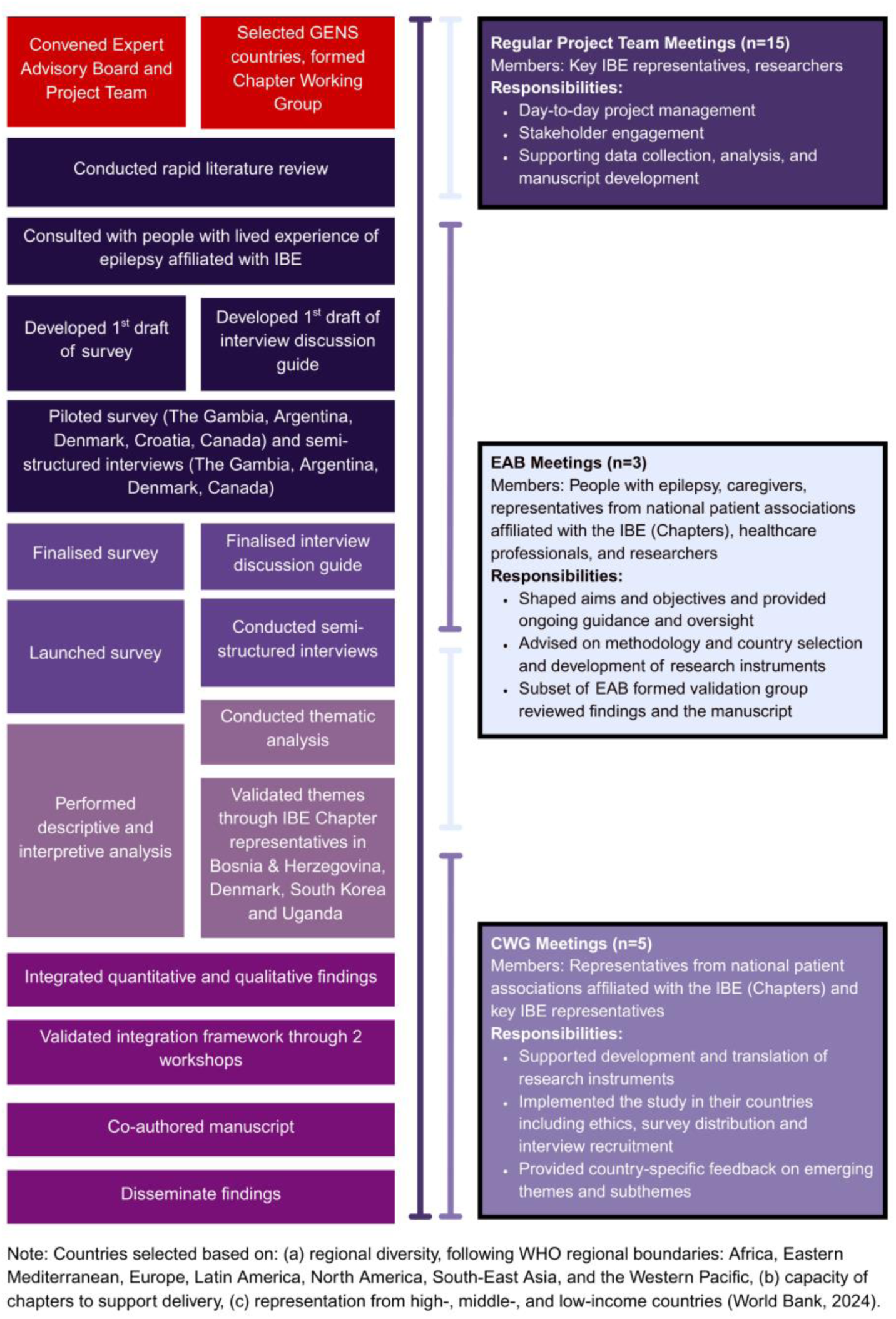
Flow chart of GENS research process, including working groups supporting the study.

### 2.2 Working Groups

The study was guided by three key working groups: the Project Team, the Expert Advisory Board (EAB), and the Chapter Working Group (CWG), with their respective responsibilities outlined in Figure 1.

### 2.3 Ethical Approval

Ethical approval or formal exemption was obtained in all 15 participating countries in collaboration with national ethics bodies between June and October 2024 (see Ethical Publication Statement).

### 2.4 Development of research instruments

The survey and interview guides were designed to explore the needs of PWE across 10 life domains. These areas were initially identified through a review of existing research, with the Quality of Life in Epilepsy Inventory-89 (QOLIE-89) serving as a key reference.^26^ These were refined with input from the EAB, CWG, and a focus group comprising people with lived experience affiliated with IBE. Their guidance ensured cultural relevance, accessibility, and grounding in lived experience (see Appendix S1 for further details on instrument development). The 10 domains were:

- Knowledge and Advice
- Safety and Survival
- Healthcare and Wellbeing
- Learning and Education
- Work and Income
- Transport and Driving
- Community and Relationships
- Mental health and Wellbeing
- Sexual and Reproductive Health
- Achieving Life Goals

### 2.5 Data collection

#### 2.5.1 Pilot testing

A pilot phase tested the survey and interview guides, leading to minor modifications. It also assessed the feasibility of data collection processes, including consent, moderator training, and secure data management. The survey was piloted in 5 countries (5-10 responses each), and single interviews were conducted in 4 countries to evaluate the feasibility of engaging local moderators.

#### 2.5.1 Survey

The survey was conducted online, via the Qualtrics survey platform between August and October 2024 with a target of approximately 250 responses per country (Figure 2). It included demographic questions and 10 dedicated sections, each corresponding to a specific domain, comprising both multi-select multiple-choice items and a limited number of open-text questions. For needs-based questions, participants selected relevant items from a predefined list and then ranked each selected item according to its level of priority (Appendix S2).

**Figure 2:**
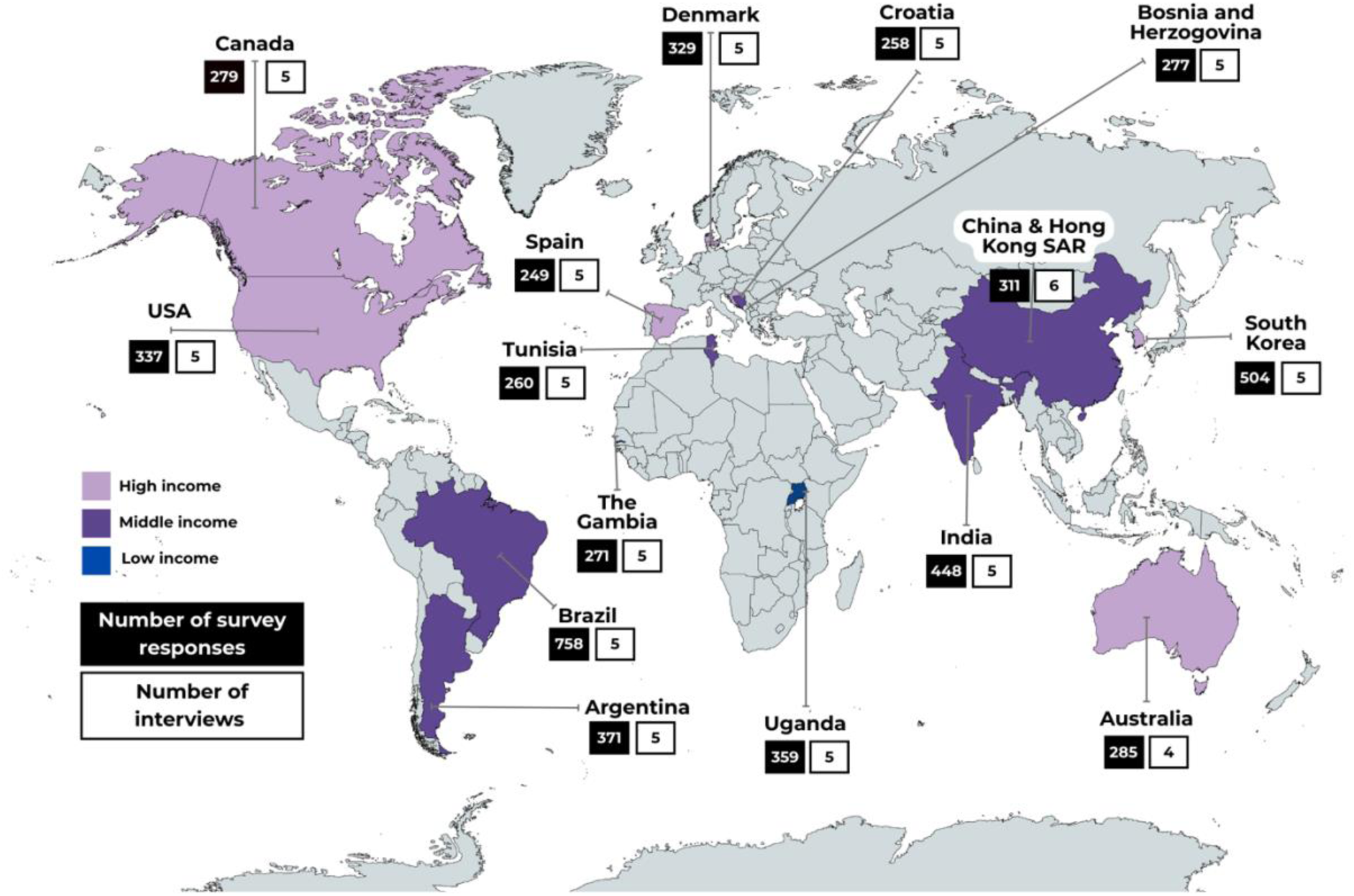
GENS focus countries.

Dissemination was coordinated by IBE Chapters through their networks, with additional promotion via IBE’s own channels, including paid advertisements in 4 countries, to meet minimum response rate. In several countries, Chapters made context-specific adaptations (e.g., engaging volunteers or leveraging local networks) to optimise reach and inclusion. These adaptations were guided by a shared methodological framework, ensuring coherence across settings. In 5 countries, trained individuals assisted participants to overcome barriers related to literacy or digital access.

The survey was developed in English and refined for accessibility using the Flesch-Kincaid readability scale,^27^ then translated into 11 languages: Arabic, Bosnian, Brazilian Portuguese, Chinese (simplified), Croatian, Danish, French (Canadian), Hindi, Korean, Spanish (Latin American), and Telugu. A forward–backward translation process was undertaken in collaboration with Chapter representatives and local moderators to ensure consistency and accuracy. Translations prioritised clear, jargon-free language to maximise accessibility across diverse linguistic and cultural contexts. Simplified Chinese, without adaptation, was used in Hong Kong SAR, China.

#### 2.5.2 Semi-structured Interviews

To ensure demographic and experiential diversity and enhance the transferability of findings, a purposive sample strategy was used to include: 1 male participant, 1 elderly participant, 1 rural-based individual, a caregiver of a person with complex epilepsy (CGC), and a caregiver of a person with non-complex epilepsy (CGN). Three exceptions occurred: in Tunisia, both caregivers supported individuals with complex epilepsy; and no rural PWE was identified (a female urban/suburban PWE replaced), though one CGC partly reflected the rural view. In Australia, no older participant was identified.

Participants received an information sheet and gave written informed consent. Interviews lasted 45 to 60 minutes, were digitally recorded, transcribed verbatim, and translated into English. Participants were reimbursed in line with local norms and ethical best practices.^28^

Semi-structured interviews were conducted in local languages by interview moderators, either in person or via remote platforms such as Zoom, depending on participant preference and local context. All moderators completed one-on-one training with the research team to review the discussion guide, confirm informed consent procedures, and ensure secure data handling.

### 2.6 Thematic analysis

A phenomenological stance guided the qualitative analysis, recognising the importance of understanding participants’ subjective experiences. To reduce potential bias, researchers (CH and SB) produced reflexive accounts to acknowledge and set aside personal assumptions – a process known as bracketing. Thematic analysis was guided by Colaizzi’s Method,^29,30^ selected for its structured rigour and focus on preserving the authenticity of participants’ voices (Figure S1). Consolidated Criteria for Reporting Qualitative Research (COREQ) was used to guide study reporting.^31^

Transcripts were coded using an inductive thematic approach. CH and SB independently coded the first 4 transcripts. Codes were discussed in detail and accepted, removed, combined or revised by consensus to ensure consistency and rigour.

### 2.7 Validation of findings

Rigorous validation processes were followed to ensure the findings accurately reflected the perspectives of PWE across diverse settings. Validation of themes from qualitative interviews was gathered via an online survey of IBE Chapter representatives in 4 countries, selected to reflect a balance of high-, middle-, and low-income settings and to avoid overlap with countries represented in the Validation Workshops (VWs). In addition, 2 VWs were held with a subgroup of the EAB and CWG, with equal representation of patients/caregivers and clinician/researcher perspectives, who provided feedback on the integrated findings to confirm, refine, or challenge the thematic interpretations.

### 2.8 Statistical analysis

Survey data were exported from Qualtrics and analysed in Stata 16.1 using scripted .DO files, ensuring transparency through an exploratory data analysis approach.^39^ Methods included frequency tables, cross-tabulations with chi-squared tests, t and F tests, scatter and box plots, and Spearman correlations. Bonferroni adjustments were applied to account for multiple comparisons, so the statement ‘significant’ after a comparison should be interpreted generally as ‘p<0.00005’ rather than the traditional 0.05. CHAID analysis^16^ was used heuristically to explore predictors of group membership, followed by logistic regression to estimate effects.

### 2.9 Integration of qualitative and quantitative findings

A joint display approach was used to compare qualitative themes with corresponding quantitative trends, highlighting areas of convergence, divergence, and complementarity. This method enabled the integration of statistical findings with lived experience insights, enriching the analysis and informing the development of the Integrated Framework (Table S1). By balancing analytical rigour with real-world perspectives, the approach provided a more comprehensive understanding of PWE needs.

## 3. RESULTS

### 3.1 Characteristics of study participants

We collected 5296 survey responses and conducted 75 semi-structured interviews across 15 countries, with China and Hong Kong SAR analysed as 1 entity. Participant characteristics are summarised in Figure 2 and Table 1.

**Table 1:**
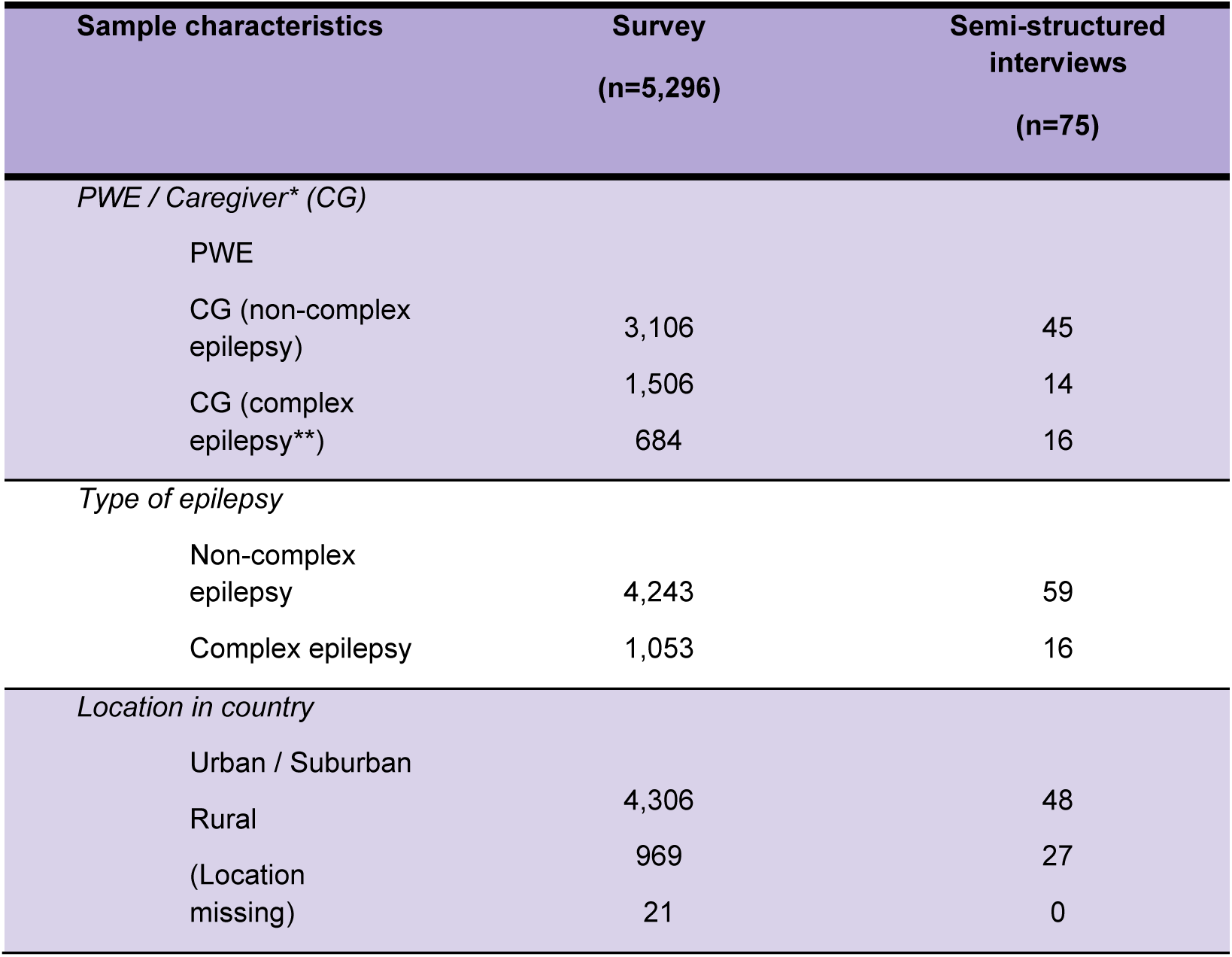
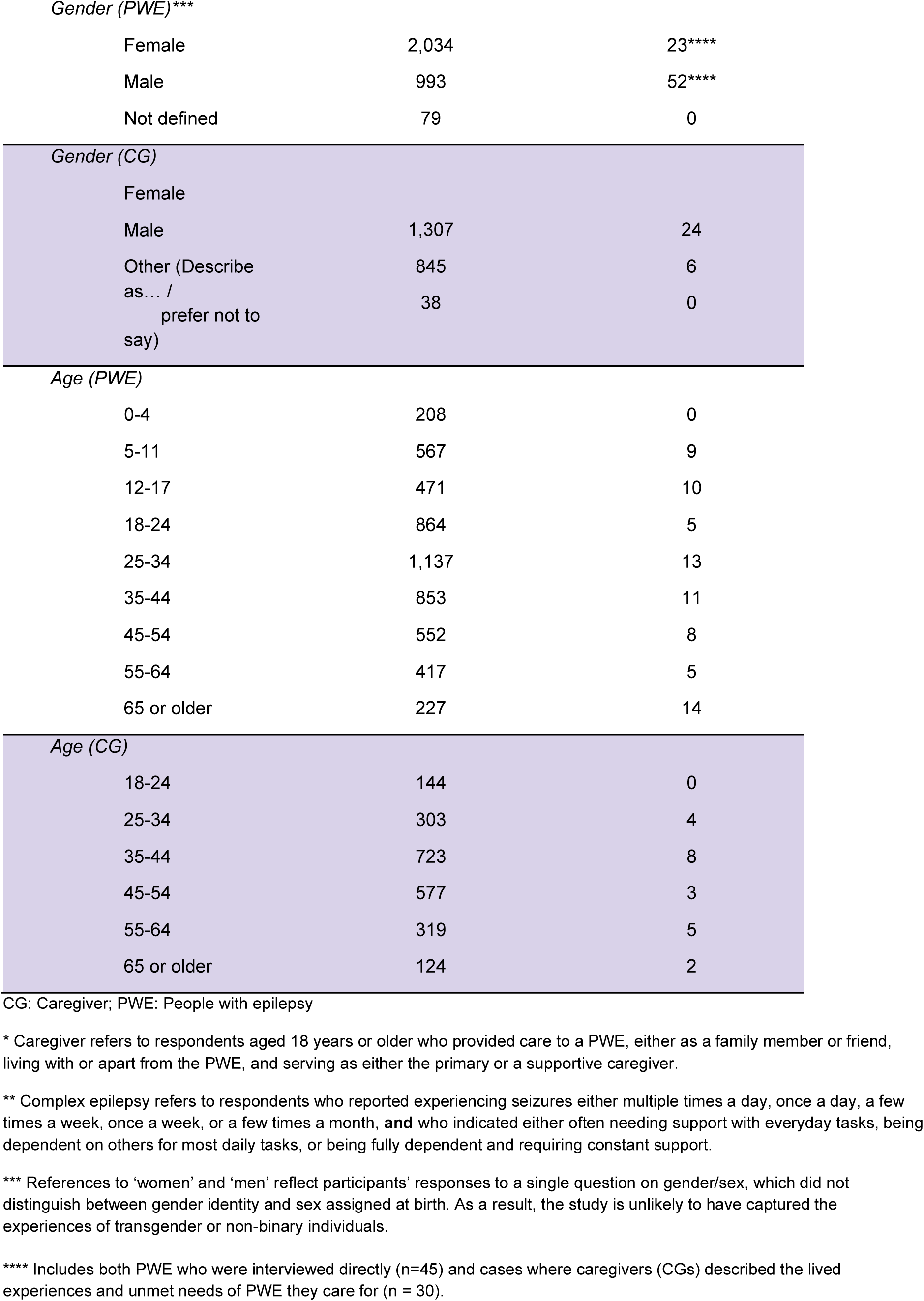
Sample characteristics.

### 3.2 Overview of findings

Only survey results that remained statistically significant after Bonferroni correction for multiple comparisons (p < 0.0001) are reported here. Figures 3 and 4 represent the top 10 most frequently selected needs, and top 3 most urgent needs across all domains, respectively.

**Figure 3:**
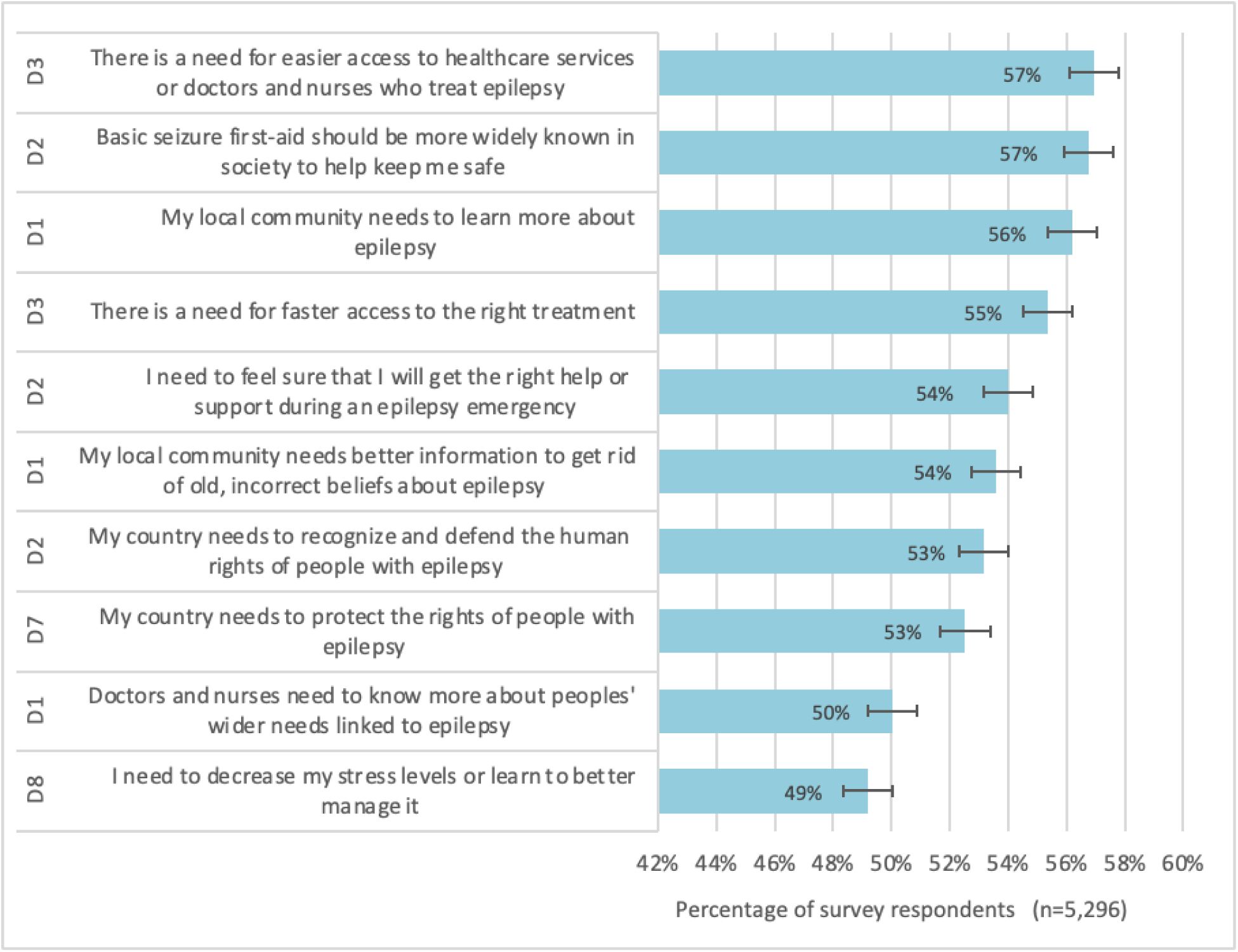
Top 10 most frequently selected needs across all survey domains (D = survey domain).

**Figure 4:**
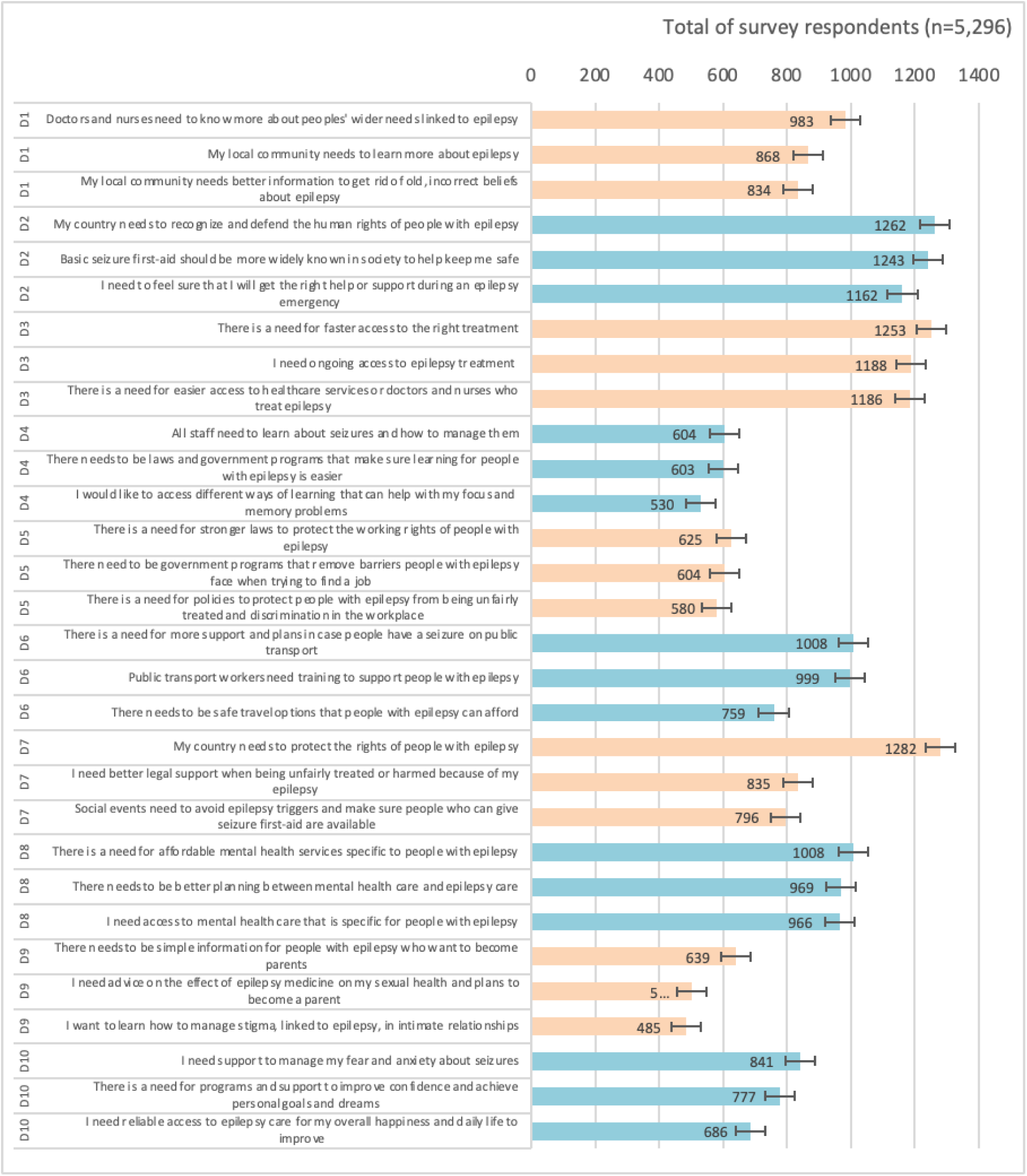
Top three needs most selected as ‘Urgent’ in each domain (D = survey domain).

Across the 10 domains, Safety and Survival (Domain 2) and Healthcare and Wellbeing (Domain 3) showed the highest average number of needs, standardised by domain items. In contrast, Sexual and Reproductive Health (Domain 9) had the lowest, with nearly half of respondents selecting “I do not have any of the above needs” - the most frequent response in this domain, and the least frequent across all others.

Response rates for Learning and Education (Domain 4) and Work and Income (Domain 5) were lower due to routing logic (Domain 4 shown only to respondents in, or planning, education; Domain 5 only to those employed or seeking work). These lower rates should be considered when interpreting findings.

Subgroup analyses showed greater needs among respondents from low- and middle-income countries, and among women, people with complex epilepsy, caregivers, those who viewed epilepsy as a disability, and self-identified minorities.

Thematic analysis of interviews generated 22 subthemes, grouped into 5 Generalised Themes (GTs) reflecting core realities across cultural and demographic settings (Table 2). As this study was designed to identify unmet needs, findings focus on gaps in support rather than existing strengths. This should be seen as reflecting the study’s aims, not an absence of positive experiences.

**Table 2:** Generalised themes and subthemes identified during thematic analysis.

| Generalised theme | Subthemes | Illustrative quotes |
| --- | --- | --- |
| **Managing uncertainty and redefining daily life**<br>The unpredictable nature of epilepsy increases vulnerability, disrupts routines, and impacts daily participation and future planning. This ongoing uncertainty creates instability in everyday life, and requires individuals and families to continually adapt their activities, plans and expectations. | Physical risk inherent to epilepsy | *"I've been recovering from a knee injury caused by a seizure I had, where my leg got stuck on the couch at home. It caused a medial collateral ligament strain."*<br>Brazil, Male |
|  | Emotional uncertainty and psychological strain | *"For those who haven't yet managed to control their seizures, there is a constant fear of having a seizure at an important moment".*<br>Bosnia and Herzegovina, Male |
|  | Barriers to everyday roles and recreation | *"I'm a little afraid of going somewhere alone...I really wanted to go to Japan in the winter, but now that I think about it, I'm a little scared...if I fall down on the street, can these people help me?... I don't think that's possible."*<br>South Korea, Rural |
|  | Parenthood, planning and uncertainty | *"If a pregnant woman has an epileptic seizure, can she lose her fetus or not? Does the medication have repercussions on fertility, especially on the fetus? Does the medication stop during pregnancy or not? All these questions were like a movie running in my head and I could not find an answer to them and could not ask them in front of anyone."*<br>Tunisia, Male |
| **Living with risk, social exclusion, and misunderstanding**<br>Stigma, judgement, societal misconceptions and safety risks shape participation and relationships, often leading to exclusion, withdrawal, strained connections and barriers to personal development. | Inadequate emergency response | *"I think people should have basic knowledge on how to behave if someone in their environment experiences a seizure....a few years ago, when a guy had a seizure in my town, and due to the inadequate help from those present, the guy unfortunately passed away."*<br>Bosnia and Herzegovina, Rural |
|  | Vulnerability and safety risks | *"In the beginning there were times when I would get a seizure and regain consciousness alone and sometimes with my property stolen."*<br>Uganda, Older |
|  | Intimacy and romantic relationships | *"I had a boyfriend when I was about 21 or 22 and his mother was bothered by my epilepsy and we broke up."*<br>Croatia, Rural |
|  | Barriers to personal growth and development | *"They dare not make friends or fall in love like normal people, always worrying that others will despise them once they know about their condition. They end up isolating themselves day by day, becoming more and more introverted and less confident."*<br>China, Older |
| **3. Challenges in navigating inaccessible systems**<br>People with epilepsy often face healthcare, education, employment, and transport systems that lack the | Barriers to accessible healthcare | *"When I go to the hospital and there is no medication, I run the risk of getting a seizure, to or from the hospital. There are extremes where I go even two months without the medication being available."*<br>Uganda, Male |
|  | Gaps in quality of care and support | *"I wish the medical staff could have a moment to exchange detailed information about the patient's* |
| flexibility, resources, or understanding required to accommodate their needs, leaving them to adapt within inflexible environments. |  | *illness. I can't even ask questions properly right now. Rather, when I try to speak as a patient carer, [HCP] sighs deeply and stops me from asking questions."*<br>*South Korea, CGC* |
|  | Barriers to mental health support | *"I was not referred for a psychiatrist by my neurologist despite the fact that I had shown symptoms of depression but it was my family who supported me and who demanded that I see a psychiatrist but late in life, I had already ruined my married life."*<br>*Tunisia, Older (Male)* |
|  | Barriers to equitable education | *"There is a lot of discrimination in the schools, because they do not know how to act. There are many cases, like the one that happened to a peer.. where they [the school] asked her to change schools because she didn't have the skills for that school and the school wasn't prepared."*<br>*Argentina, Rural* |
|  | Structural barriers to employment and retention | *"epilepsy excludes you from many public job competitions...in every competition for police positions, when you look at the list of medical conditions that disqualify candidates, epilepsy is almost always one of the first on the list under nervous system disorders...there isn't the same kind of inclusion policy that I see in other countries."*<br>*Brazil, Male* |
|  | Everyday challenges of epilepsy in the workplace | *"I have lower vitality because of the seizures my energy levels are lower than I'd typically expected to be. My ability to work is significantly reduced because of having epilepsy. I'm a barber but still I don't get as much work as I would have if I didn't have the condition."*<br>*Uganda, Male* |
|  | Health-related mobility challenges | *"I need to be well-rested and I don't travel alone. On longer trips, my wife and I take turns so that we don't drive for long periods at a time. I can only drive for three hours at a time, and it's becoming increasingly difficult."*<br>*Croatia, Male* |
|  | Transport accessibility and infrastructure gaps | *"Unfortunately, there aren't many conveniences available regarding transportation. While some buses have ramps, they aren't present in all buses or at all stops. Additionally, marked parking spaces exist but are often disrespected by others."*<br>*Bosnia & Herzegovina, CGN* |
| **4. Consequences of inaccessible or inadequate information**<br>A lack of timely, relevant, and accurate information about epilepsy, at individual, healthcare professional, and societal | Health information shortfalls in healthcare | *"I believe that doctors and nurses are taught how to manage seizures during their training, but these emotional and psychological aspects seem to be overlooked."*<br>*Brazil, Male* |
|  | Knowledge gaps in everyday and future planning | *"Maybe some [support] programs do exist, but even if they do, they are not sufficiently accessible to people...if there are programs for people with epilepsy, individuals must research* |
| levels, creates uncertainty, reinforces stigma, and limits the ability to make informed decisions and access appropriate support. |  | *and discover them on their own, which is a big problem.”*<br>*Bosnia & Herzegovina, Male* |
|  | Widespread misunderstandings of epilepsy | *“They think he has a bad disease and are afraid of being affected, so they don't want to have anything to do with him anymore.”*<br>*China, CGN* |
| **5. Complex epilepsy needs demand more than standard approaches**<br>People with rare or complex epilepsies face persistent challenges unmet by standard care, including fragmented healthcare pathways, unmet cognitive and emotional needs, gaps in specialist medical and psychosocial support and heavy caregiving burdens. | Access to epilepsy care | *“My biggest frustration...was the amount of work I had to do as an advocate to get his needs met...First up, he has a seizure...seven week wait...then a six month wait...just get passed along. Limited communication between everyone...we actually took him twice...to [US] for care to skip the six month wait times...”*<br>*Canada, CGC* |
|  | Cognitive, emotional and developmental needs | *“He's very impulsive and he doesn't have a concept of like, safe surroundings. He would cross the street without looking twice, comparatively, to kids his age.”*<br>*Canada, CGC* |
|  | Burden of caregiving | *“She picks her skin non-stop. You literally can't leave her for more than two minutes by yourself...They can't do anything themselves. I mean, the only thing they could do themselves is get their iPad and use it. Everything else they rely on people.”*<br>*Australia, CGC* |

CGC: Caregiver of a person with complex epilepsy; CGN: Caregiver of a person with non-complex epilepsy

### 3.3 Integration of qualitative and quantitative findings

The integrated findings are presented within 5 overarching GTs, synthesising qualitative and quantitative results. This section provides a high-level overview, with additional detail in Table S1 and further insights to follow in subsequent publications.

### GT1: Managing uncertainty and redefining daily life Physical risk inherent to epilepsy

Women were more likely than men to report a need to reduce ‘risks of life-changing injuries or dying from epilepsy’ (47% vs. 40%; OR = 1.4). However, Sudden Unexpected Death in Epilepsy (SUDEP) was rarely mentioned in interviews, suggesting a partial divergence between quantitative and qualitative data. Interview accounts focused on immediate risks, indicating that while fear of serious harm is widespread, awareness of SUDEP may vary across settings.

### Emotional uncertainty and psychological strain

The most frequently selected need in Domain 10 was ‘support to manage my fear and anxiety about seizures’ (44%). Interviews described seizures as unpredictable, fostering “constant fear” (China, Older). Burden of concealment was another source of psychological strain: “the stigma…is still very strong, so many are afraid to openly talk…which is mentally and emotionally exhausting.” (Bosnia and Herzegovina, Male).

Women were more likely than men to report needs to ‘cope with emotions’ (48% vs. 39%; OR = 1.4), ‘cope with feeling isolated’ (39% vs. 34%; OR = 1.3), and ‘decrease stress’ (53% vs. 44%; OR = 1.4). Both genders linked inconsistent or inaccessible care to disrupted routines, distress and overall burden. Women more often spoke of persistent worry, exhaustion, and need for psychological help, *“…I felt as if a heavy burden had been placed on my heart*” (China, Older). Men discussed the need for psychological help less, instead linking impacts to shifts in confidence, independence, or cognitive function. Across genders, participants described strategies to manage stress, highlighting it as a common need.

### Barriers to everyday roles and recreation

Respondents who self-identified as having a disability were more likely to report the need to feel ‘well enough to help with household chores’ (29% vs. 21%; OR = 2.0). Interviewees described how fatigue, risk of injury, and family restrictions limited contributions at home. Many shared examples of adapting their home environments to maintain independence, e.g., *“[buying an induction oven] to feel safe…”* (Canada, Rural). Cognitive difficulties, such as memory problems, also disrupted leisure: *“…the personal trainer tells us, ‘Well, we’re going to this, this, this’…For me, it was like he spoke in Japanese, because there was no way I can retain that.”* (Argentina, Male).

### Parenthood, planning and uncertainty

In Domain 9, 36% selected the need for ‘simple information for PWE who want to become parents,’ although nearly half reported no needs. Both datasets highlighted gaps in sexual and reproductive health information. When raised in interviews, decisions around parenthood were described as complex and emotionally charged, shaped by concerns about medication safety, genetic transmission, and parenting with epilepsy: *“…if I passed my epilepsy on to my child, I would be very guilty…”* (South Korea, Rural).

### GT2: Living with risk, social exclusion, and misunderstanding

### Inadequate emergency responses

Among the top 5 needs were: ‘basic seizure first-aid should be more widely known’ (57%), ‘community needs to learn more about epilepsy’ (56%), and ‘receiving the right help during an epilepsy emergency’ (54%). Women were more likely than men to prioritise seizure first-aid (60% vs. 52%; OR = 1.4), help during epilepsy emergencies (57% vs. 49%; OR = 1.4), and safety during emergencies such as conflicts or disasters (49% vs. 42%; OR = 1.4). Interviews described delayed or harmful responses during seizures, with women particularly expressing fear and trauma related to such incidents in public settings.

### Vulnerability & safety risks

Over half of respondents highlighted ‘harmful and deep-seated community beliefs’ as a challenge (see GT4). Interviews illustrated how such beliefs affected education and employment, leading to bullying, stigma, and withdrawal: “…I am afraid they will bully me, as they do with other students who have similar [conditions] to mine” (Tunisia, Male); “…[they] are afraid of being affected, so they don’t want to have anything to do with him anymore” (China, CGC).

Respondents from low-income countries were more likely to report a need to ‘feel safe from being physically hurt by others’ (low: 73% vs. middle: 31% vs. high: 22%; OR = 11.6). Qualitative accounts described theft, abuse, and in severe cases, fatal violence during seizures: “He was beaten up and he died of his injuries. I have a feeling that he might have been like me.” (Uganda, Male).

Women were also more likely to express this need (34% vs. 29%; OR = 1.3) and a need to feel safe from emotional harm (44% vs. 36%; OR = 1.4). Men more often described physical injuries; with stigma, shame, and withdrawal being common to all. Caregivers of women and other marginalised genders^1^ raised particular concerns about abuse in intimate and domestic contexts.

### Intimacy and romantic relationships

Respondents from low-income countries were more likely to need ‘advice on how to tell key people in their life’ about epilepsy (high: 68% vs. middle: 25% vs. low: 27%; OR = 7.9). Interviews revealed the disclosure dilemma, with many fearing rejection in romantic contexts: “If a child has it…people don’t tell about their condition as children have to get married later in life.” (India, Rural). Participants also described seizures undermining intimacy, anxiety about having seizures during sex or confusion between seizure symptoms and sexual responses, and rejection after seizures: “she couldn’t handle that…I never saw her again” (Denmark, Male).

Long-term relationships could also be impacted, with participants describing emotional strain and shifts in caregiving or financial roles: “I was supposed to be the trophy husband. She wanted to be the one that brought the money…But that was not what happened…She had to start counting on me…I had to take over leadership…that was not what she wanted” (US, CGN).

### Barriers to personal growth and development

Forty per cent of respondents reported a ‘need for programs and support to improve confidence and achieve personal goals’. Interviews echoed this, describing systemic exclusion that hindered personal growth (see GT3), including from leadership roles and civic participation: “…one of the community members informed the organiser that I am someone who lives with epilepsy and I cannot take this responsibility [Chair of community workshop]…I’ve never been selected” (The Gambia, Older). Throughout, participants voiced concern at the lack of government protections to foster inclusion.

### GT3: Challenges in navigating inaccessible systems

### Barriers to accessible healthcare and Gaps in quality of care and support

The most commonly selected need across all domains was ‘easier access to healthcare services’ (57%). Most interview participants described services as fragmented, under-resourced, with long waits and limited follow-up. Therefore, the burden of coordinating care often fell to PWE and caregivers. One caregiver, concerned about delays, stated, “We ended up paying out of pocket” (Canada, CGC).

‘Faster access to the right treatment’ ranked fourth across all domains and subgroups (55%) and was significantly more likely to be prioritised in low-income countries (low: 85% vs. middle: 56% vs. high: 48%; OR = 5.3). Similarly, ‘ongoing access to epilepsy treatment’ was more often selected in low-income settings (low: 83% vs. middle: 49% vs. high: 39%; OR = 6.2). Interview participants highlighted shortages, long travel distances, and unaffordable medicine: “Another major concern is medication…many times it was out of stock” (Brazil, CGC) and “…treatment took a long time because the medicine is not available in my country” (Tunisia, CGC).

### Barriers to mental health support

Most interview participants described a lack of coordination between epilepsy and mental health services, with support often unavailable until distress escalated. Women were more likely to report a need for ‘mental health care specific for PWE’ (47% vs. 40%; OR = 1.4) and ‘affordable mental health services’ (48% vs. 41%; OR = 1.4). Although gendered patterns were less evident in interviews.

Rural respondents were more likely to report needing to ‘decrease stress levels or learn to better manage it’ (rural: 57% vs. suburban: 50% vs. urban: 47%; OR = 1.5) and ‘support to manage fears or anxiety about seizures’ (rural: 46% vs. suburban: 43% vs. urban: 43%; OR = 2.0). While this rural pattern was less distinct qualitatively, some rural participants explicitly expressed such unmet needs: “…psychological support is more important than someone helping me if I had a seizure…” (Croatia, Rural).

### Barriers to equitable education

Respondents from lower-income countries were more likely to report the need for teachers to ‘learn about epilepsy so they don’t have unfair beliefs’ (low: 78% vs. middle: 59% vs. high: 60%; OR = 2.7). Interviews across all income groups described students being denied inclusive opportunities, lacking adjustments, or facing stigma and academic pressure that triggered seizures: “Once I had a seizure during an anatomy exam…there wasn’t much support from the institution…some professors thought I was pretending…” (Brazil, Male).

Caregivers described education as a foundational priority: “…most importantly, other than medication, is access to education” (The Gambia, CGN). A few of them across subgroups also noted concerns about absent seizure-response protocols and teachers misunderstanding epilepsy.

### Structural barriers to employment and retention **and** Everyday challenges of epilepsy in the workplace

Respondents from low-income countries were more likely to report a need for workplace inclusion and relationship-building with colleagues (low: 55% vs. middle: 24% vs. high: 19%; OR = 10.9). Interviews showed this concern was universal, with participants citing stigma, disclosure anxiety, and fear of negative reactions after seizures. Some described structural barriers such as exclusion from jobs due to driving restrictions, disrupted skill development, and fear of accidents. Participants also reported everyday challenges, including memory issues, post-seizure fatigue, and constant worry about safety in the workplace.

### Health-related mobility challenges and Transport accessibility and infrastructure gaps

The need for ‘training (public transport workers) to support PWE’ ranked among the top three priorities across all income groups. Women were more likely than men to express a need to ‘feel safe when travelling’ (46% vs. 38%; OR = 1.4). Rural respondents were more likely to report needing ‘support to travel for work’ (rural: 33% vs. suburban: 27% vs. urban: 25%; OR = 1.6). Some interview participants concurred, describing unsafe or inaccessible transport and anxiety about seizures in transit. Such fears contributed to withdrawal and reliance on others. Mobility challenges were compounded by driving restrictions, which reduced independence and increased reliance on caregivers or public systems. PWE were more likely than CGs to report that ‘driving restrictions limited their independence’ (34% vs. 24%; OR = 1.9), reversing the general trend of caregivers reporting higher needs. As 1 participant reflected: “…the biggest down…lost my independence…like a hammer blow.” (Canada, Older).

### GT4: Consequences of inaccessible or inadequate information Health information shortfalls in healthcare

The need for ‘doctors and nurses to know more about people’s wider needs’ was the third most-selected item within Domain 3. Interview participants frequently echoed this, citing a lack of holistic care and limited professional knowledge that left families without guidance or support: “We are holistic integral beings…I don’t think this is taken into account. We’re not just electrons in the brain.” (Argentina, Male); “…I went to a neurologist who asked ‘What is it? Tell me what it is?’ He wanted me to tell him what epilepsy is.” (Brazil, Rural). In the absence of clear, trusted guidance, participants often turned to online forums or peers, and sometimes paid out of pocket for second opinions.

Respondents from low-income countries were significantly more likely to report needing ‘access to high-quality information’ (low: 77% vs. middle: 46% vs. high: 35%; OR = 7.0) and to state that ‘doctors and nurses need to know more about people’s wider needs’ (low: 77% vs. middle: 46% vs. high: 49%; OR = 4.4). Qualitative accounts from these settings described basic gaps such as diagnoses or medications given without explanation or follow-up: “healthcare professional only prescribed…but I was not informed it was epilepsy” (The Gambia, CGN). By contrast, participants in higher-income contexts more often highlighted missing emotional support and broader aspects of care, rather than core diagnostic or treatment information.

### Knowledge gaps in everyday and future planning

Interviews identified gaps in personal knowledge related to, for example, symptom understanding and awareness of available services. Information to enable future planning was often deemed inadequate (e.g. parenthood - see GT1).

Survey data showed caregivers in low-income countries were significantly more likely to report a need for ‘training on epilepsy management’ (low: 88% vs. middle: 32% vs. high: 28%; OR = 20.2). Caregivers across settings more often described general gaps in family and community knowledge, with few explicitly mentioning ‘training,’ suggesting this is an emerging need rather than an established expectation

### Widespread misunderstanding of epilepsy

Respondents from low-income countries were more likely to report a need for families to ‘learn more about epilepsy’ (low: 80% vs. middle: 34% vs. high: 29%; OR = 11.7) and for ‘better information to get rid of old, incorrect beliefs’ (low: 80% vs. middle: 53% vs. high: 50%; OR = 5.5). The latter ranked as the sixth most frequently selected need across all subgroups.

Qualitative findings across low-, middle-income, and rural settings described inaccurate community beliefs and harmful practices, often reinforced by family or neighbours. Such myths frequently led to fear, overprotection, and isolation for PWE: “[participant’s mother] says make the person smell onion to stop their seizure…someone from the community might have told her” (India, Rural)

### GT5: Complex epilepsy needs demand more than standard approaches

### Access to epilepsy care

People with complex epilepsy were significantly more likely to report a need for ‘easier access to epilepsy healthcare services’ (66% vs. 55%; OR = 1.8) and dedicated ‘complex epilepsy services’ (53% vs. 38%; OR = 1.9). While healthcare addressing ‘specific needs’ was among the least selected overall, it was significantly more likely to be chosen by this group (47% vs. 37%; OR = 1.6). Caregivers described healthcare pathways as “archaic” and “disjointed” (Australia, Tunisia, CGC) and marked by “a clear lack of multidisciplinary approach” (Spain, CGC). Many reported coordinating care themselves, including chasing referrals or making treatment decisions with minimal guidance.

### Cognitive, developmental and emotional needs

Interviews highlighted the multifaceted nature of complex epilepsy, often accompanied by impairments in attention, memory, learning, speech, and emotional regulation. Despite these needs, caregivers described a striking lack of proactive support: “When [parents] go to a paediatrician and mention epilepsy or intellectual disability, they often just throw their hands up and say they can’t help.” (Bosnia and Herzegovina, CGC). Those with complex epilepsy (48% vs. 37%; OR = 1.6) and respondents viewing epilepsy as a disability (47% vs. 32%; OR = 2.0) were also more likely to report needing help managing ‘other health problems’. Many caregivers expressed frustration when HCPs lacked knowledge or confidence to address co-occurring or neurodevelopmental challenges, deepening gaps in care.

When asked about SUDEP directly, caregivers of individuals with developmental delays or impulsive behaviours more often described risks of immediate physical harm: “…probably more likely to choke than to die of SUDEP…” (Australia, CGC) and “jumping in the pool without thinking” (Canada, CGC) and the constant vigilance required to prevent these.

### Burden of caregiving

Caregivers of people with complex epilepsy were significantly more likely to report a need for ‘inclusive social activities for caregivers and those they care for’ (54% vs. 41%; OR = 2.2). Interviews illustrated how the demands of care often led to exhaustion and isolation, limiting participation in everyday life despite a strong desire for connection: “she does not go to a specialised association. How I wish I could find an association that can frame such cases. On the one hand the child finds specialists who take care of them in the right way, and on the other hand the mother finds an outlet to renew her energy” (Tunisia, CGC).

While the study’s primary focus was the needs of PWE, caregiver perspectives (some detailed in the Table S1) will be explored in future publications.

## 4. DISCUSSION

To our knowledge, GENS is the first large scale, mixed-methods, multi-country study to explore the unmet psychosocial and everyday needs of PWE. By capturing lived experience across diverse settings, the study provides a unique and comprehensive understanding of the challenges faced by PWE globally. It highlights shared experiences and also key variations across subgroups offering critical insights to inform more inclusive and person-centred epilepsy research, care and support.

The results of the study identified 5 major areas of need:

- Managing uncertainty and redefining daily life
- Living with risk, social exclusion, and misunderstanding
- Challenges in navigating inaccessible systems
- Consequences of inaccessible and inadequate information
- Complex epilepsy needs demand more than standard approaches

### Managing uncertainty and redefining daily life

Our findings reinforce the well-established body of research demonstrating that uncertainty is a defining feature of life with epilepsy; spanning social, emotional, and physical domains. This uncertainty stemmed not only from the unpredictability of seizures but also from the stigma that continues to surround the condition. While concealment was often adopted as a strategy of self-protection, it contributed to worsening isolation and emotional distress, echoing the observations of Bauer et al.,²¹ Sherlock et al.,¹³ and Strzelczyk et al.⁵ Tackling stigma, therefore, remains central if wellbeing is to be meaningfully improved.

Despite longstanding recognition that comorbidities form an integral part of comprehensive epilepsy care,¹⁸ participants consistently described this as a neglected need, with the emotional toll of navigating fragmented and inconsistent systems too often borne alone.

Gendered differences in our accounts of psychological strain offer important insights into how care must be tailored. Women more frequently described persistent worry, exhaustion, and the need for psychological support, whereas men linked their experiences to loss of confidence, independence, and cognitive functioning. Parenthood decisions, particularly concerns around pregnancy and genetic transmission, emerged as areas of uncertainty, compounded by fragmented advice and inconsistent guidance from HCPs - gaps that echo earlier reports and remain urgent to bridge.^11, 12^

Safety-related restrictions, though intended to protect, were often experienced as overbearing and stifling, eroding autonomy. The loss of driving privileges was described as a profound loss of independence, raised more frequently by PWE than by caregivers, suggesting that its daily significance may be underestimated even by those closest to them. Strzelczyk et al.⁵ similarly reported PWE feeling ‘suffocated’ by protective measures and identified driving as one of the most affected domains of QoL.

### Living with risk, social exclusion, and misunderstanding

Living with epilepsy means living with risk - not only the inherent risks of seizures but also those created by others’ actions, beliefs, and misunderstandings. The fact that more than half of participants identified improved public understanding of seizure first aid, greater community awareness, and the dispelling of outdated myths as priority needs, emphasises how urgently these misconceptions must be addressed. Participants described experiences of fear, avoidance, and even violence, particularly in low-resource settings, echoing earlier accounts of epilepsy being associated with contagion or insanity,²⁴˒³⁴ and reinforcing the pressing need to challenge such beliefs. While both men and women described risks of harm, caregivers of women with epilepsy more frequently raised concerns about vulnerability to abuse, particularly in domestic contexts, highlighting how gendered differences continue to shape the daily realities of living with epilepsy.

Cultural stigma emerged as a major barrier to wellbeing. Misbeliefs eroded self-esteem, limited access to care, and discouraged disclosure. Social exclusion - through both marginalisation and self-isolation - was widely reported by PWE and caregivers. Such experiences - which diminish participation in education, employment, and social life - mirror previous findings on the intersection of stigma, mental health, and reduced self-worth,⁵˒³⁵ and emphasise the need for healthcare and policy approaches that acknowledge cultural context.

Disclosure was a particular concern across settings. In low-income contexts, ingrained stigma affected openness in personal and professional relationships. Women were particularly affected, as also noted by Gosain and Samanta,²⁴ especially in relation to marriage and parenthood prospects. While some described supportive relationships, others spoke of strain or co-dependence, illustrating how epilepsy can reshape the balance of relationships. In high-income countries, fear of judgement in schools, workplaces, and social environments was evident - demonstrating once again how policy priorities must be adapted to context.

### Challenges in navigating inaccessible systems

Difficulties in accessing and navigating healthcare systems emerged as a dominant concern, with over half of participants identifying easier access as a top need. Accounts of fragmented systems, long waits, and limited specialist provision echo previous findings;^36,11^ illustrating how poorly coordinated, siloed pathways often leave families managing care themselves.

In low- and middle-income countries, treatment was further disrupted by medication shortages, long travel distances, and prohibitive costs, leading to irregular adherence, forced switches, and considerable anxiety, as also noted by Makasi et al.^15^ Ponza et al^36^ note that factors such as public sector shortages and financial barriers also compromise continuity of care in high income countries, highlighting that access challenges are not only a concern in low resource settings. That reliable access remains uncertain is concerning, particularly given its established role as an independent predictor of QoL in PWE.^37^

Mental health support was often disconnected from epilepsy care. Nearly half of participants called for greater integration, often describing psychological help as available only in moments of crisis, if at all, despite emotional needs being persistent and central to wellbeing.

Beyond healthcare, education emerged as a critical concern. Inadequate seizure protocols, stigma, and exclusion were widely reported, echoing global findings of bullying and isolation,^8^ and showing how schools - instead of being safe spaces - can be precarious settings for PWE. Such experiences compromise wellbeing and risk undermining academic attainment and social development in the long-term.

Similar barriers arose in employment and transport. Stigma, inadequate workplace accommodations, and in rural areas, long travel distances and high costs all restricted participation. Work by Souza et al. shows how unemployment can reinforce cycles of disadvantage, limiting access to medication, mental health support, and wider social inclusion.^38^ Together, these findings highlight the need for legislative and systemic reforms that protect dignity and inclusion in daily life for PWE.

### Consequence on inaccessible or inadequate information

Our findings highlight the gaps in both clinical and community knowledge about epilepsy, and it’s profound effects on everyday life. Reports of limited confidence and expertise among general practitioners, and even neurologists, highlights a critical weakness in the care pathway for PWE. This is especially concerning given that poor management of epilepsy medication (e.g. side effects) is linked to lower QoL.^39^ For women of childbearing age, concerns around inheritability and medication safety were especially pressing, reflecting the high stakes of these knowledge gaps. This aligns with earlier work by Çelik and Kaya,^19^ who found many physicians felt unprepared to manage epilepsy, particularly in the context of pregnancy.

Gaps in formal knowledge were compounded by widespread cultural misunderstandings described earlier. In low resource settings, as outlined by Makasi et al.; Gosain and Samantha; and Tarhini et al. ^15,24,24^, these ingrained beliefs about contagion and insanity not only delayed care but amplified stigma and exclusion. Yet, even in high-income settings, participants spoke of misinformation shaping their interactions and limiting their confidence in social support systems, showing that these issues are not confined by geography.

Caregivers, particularly those supporting individuals with complex epilepsy or in low-income settings, often felt untrained and unsupported, echoing earlier reports.^40,41^ Their accounts point to the urgent need for equipping caregivers with the knowledge and skills required to manage complex and multifaceted needs of PWE.

### Complex epilepsy needs demand more than standard approaches

The voices of caregivers predominantly informed our findings on complex epilepsy. Although intertwined with their own experiences, these accounts offered a valuable lens into the everyday realities of people living with complex epilepsy. The relentless treatment demands, high seizure frequency, and accompanying emotional exhaustion described by caregivers reflect the heavy and enduring burden borne by PWE. These challenges contribute to a broader psychosocial strain that extends beyond the individual to affect family dynamics, stability, and overall QoL - factors known to be central to the wellbeing of PWE.^35,42^

Concerns reported around long-term care, ageing caregivers, and the absence of structured support networks raise important questions about the future security and autonomy of this population. Within this context, the financial pressures reported, particularly in lower-resource settings, as supported by exiting literature,^40,41^ emerge not only as a strain on caregivers but as a critical determinant of the health, safety, and life opportunities of PWE.

Across settings, access to healthcare was consistently prioritised. Participants described ‘disjointed’ pathways, often lacking multidisciplinary input or attention to developmental and cognitive challenges. Together with reports of HCPs’ limited confidence and experience, these highlight the inadequacies of current systems in meeting the needs of this population.

Finally, caregiving itself was described as exhausting and isolating - a burden acknowledged in the epilepsy literature but still under-researched.^43^ The need for inclusive opportunities that build resilience and foster social connection was strongly expressed, reinforcing how complex epilepsy not only demands holistic care for the individual but also structured support for the families who care for them.

This paper presents the overarching results of GENS, with future work set to explore in greater depth the challenges faced by underserved groups, including caregivers, individuals with complex epilepsy, women, and those in low-income or rural settings. Yet even now, the priorities for action are clear. Strikingly, 2 of the 10 most frequently selected needs called directly for the recognition and protection of the human rights of PWE, a thread that ran consistently through the 5 themes. Our findings showcase the necessity of IGAP’s epilepsy-specific strategic objective, which calls on governments worldwide to strengthen the public health approach to epilepsy; with a focus on improving access to care, and safeguarding human rights. Beyond documenting unmet needs, these results provide an evidence base to inform and educate the epilepsy community and wider public, while guiding the work of organisations such as IBE, ILAE, and their national Chapters. Together, they signal a pressing call for healthcare systems to embrace holistic, multidisciplinary models of care, and for policymakers to invest in systemic reforms that ensure dignity, inclusion, and life opportunities for PWE.

## 5. LIMITATIONS

Methodological limitations exist, particularly in cross-context comparability. First, although the survey reached over 5000 participants, recruitment was limited to 15 countries which could limit the generalisability of findings globally. Second, core study materials were translated and locally adapted. This approach may have introduced subtle variations in meaning that could have influenced how participants understood and responded to questions. Third, the scope of the survey was limited by the need to balance comprehensiveness with feasibility. This may have resulted in the omission of relevant areas of need not captured in the final instruments. Fourth, qualitative interviews were conducted by local moderators, potentially leading to variations in interviewing style or probing. Finally, although reflexivity and patient community validation were built in, the interpretative nature of phenomenological analysis means the researchers’ perspectives inevitably influenced the interpretation of qualitative data.

## 6. CONCLUSION

As the first comprehensive, mixed-methods global study of its kind, GENS provides robust evidence on the unmet needs of PWE, drawing on the voices of more than 5000 participants worldwide. The findings make clear that epilepsy is defined not only by seizures but by the daily challenges of uncertainty, isolation, and diminished autonomy, with stigma and exclusion shaping lived experience. Across settings, care remains fragmented, reactive, and poorly integrated; with systemic challenges extending into education, employment, and transport. Personal and public misinformation and limited expertise of HCPs further exacerbate these challenges, especially for those living with complex epilepsy. Taken together, these findings serve both as a mirror and a map - reflecting the realities of life with epilepsy, and signaling clear priorities for action.

## Data Availability

The original contributions generated for this study are included in the article and its Supporting Material (the Integration Framework developed for the GENS study). Further inquiries can be directed to the corresponding author.

## ACKNOWLEDGMENTS

The following pharmaceutical companies have provided financial support to the development of Phase 1 of the Global Epilepsy Needs Study – documented in this manuscript – through grants to the International Bureau for Epilepsy: Angelini Pharma, Jazz Pharmaceuticals, Takeda Pharmaceuticals and UCB.

These companies have had no editorial influence on this research article.

Funds were used to contract the research consultancy (MediPaCe), cover project management fees (for IBE and its chapters), reimburse contributors, support translation and dissemination costs, and offset other miscellaneous expenses.

During the preparation of this work the author(s) used ChatGPT (V.5) to improve grammar and sentence clarity. The author(s) reviewed, edited, and verified all AI generated suggestions and bear full responsibility for the final interpretation and presentation of the findings.

We would also like to sincerely acknowledge the contribution of a number of people not included in the authorship or collaborator group. These people have contributed across the different phases of the project and we offer our genuine appreciation. The IBE Community Council, whose encouragement, involvement and counsel has continually shaped the GENS project. The nine focus group members with lived experience of epilepsy from around the world, whose advice and guidance shaped the development of the research tools. Members of the IBE International Executive Committee; Graeme Shears and Man Mohan Mehndiratta; and staff team Niamh O’Neill, Marie Ennis O’Connor & Elizandra Cripps for input, direction and editing. And Sebastian Winter, our former colleague, whose input formed the foundations of our project plan.

We know that our chapters often had volunteers or staff members who supported delivery of this project on the ground. Please know this project could not have happened without you, and we remain truly grateful.

Finally, to the wider IBE community - these results are for you and we hope they will drive progress and impact for people with epilepsy around the world.

## Footnotes

1 One of the PWE referred to in this context was non-binary.

