## Supplemental Appendix 1 for "The Global Epilepsy Needs Study (GENS): A mixed-methods, multi-country exploration of the unmet psychosocial and everyday needs of people with epilepsy"

### METHODS

#### Domain Development Process

The development of these domains followed a structured and iterative process. An initial literature review was conducted to identify existing studies examining the unmet needs of PWE and the impact of epilepsy on quality of life. The Quality of Life in Epilepsy Inventory-89 (QOLIE-89)[29] was identified as a key resource. Its seven domains (cognitive functioning, emotional wellbeing, social functioning, seizure worry, medication effects, energy/fatigue, and overall quality of life) served as a conceptual foundation.

Building on these established areas, the Project Team developed 10 new domains that extended and deepened the coverage of life areas identified in QOLIE-89. Draft domains were subsequently presented to members of the Expert Advisory Board (EAB) for feedback and refinement, through online consultation. Additional online input was gathered from representatives of the Chapter Working Group (CWG). Further consultation was conducted via a dedicated focus group with a range of people with lived experience of epilepsy affiliated with IBE (members of a lived experience council, IBE's Global Youth Team, and other chapter representatives from different regions of the world) seeking final feedback on the domains and early identification of priority areas within the domains that would later inform the development of questions and 'needs statements' in the survey and interview discussion guides.

#### Survey Design Considerations

In parallel, detailed feedback on survey design was gathered from EAB and CWG members to enhance accessibility, inclusivity, and cultural relevance. Key design considerations included the need to accommodate a wide range of subgroups, including caregivers, individuals with rare or complex epilepsies, those with lower literacy levels, and underrepresented or vulnerable demographics (e.g. older adults, people identifying their epilepsy as a disability).

Stakeholders emphasised the importance of:

- Using plain and culturally appropriate language
- Asking simple, unambiguous questions
- Including definitions where needed to aid comprehension
- Ensuring flexibility in administration (e.g. self-completion or proxy response)
- Keeping the survey as brief as possible as long surveys were seen as burdensome for respondents, particularly those with cognitive difficulties

The final version of the survey was developed in English and refined to improve readability using the Flesch-Kincaid readability scale. For translated and adapted versions, emphasis was placed on using clear, jargon-free, and easy-to-understand language to support accessibility across all contexts. The Spanish version was used in both Spain and Argentina with moderators given discretion to adapt wording as appropriate.

#### Feedback on Draft Instruments

Following domain and survey structure development, extensive rounds of feedback and review were undertaken. Early drafts of the survey and interview discussion guide were shared with EAB and CWG members. Their feedback informed refinements across several areas, including:

- Streamlining the overall structure of the survey
- Further simplification of language for clarity
- Neutralising phrasing to avoid negative or leading tones
- Broader considerations for demographic questions

- Clarifying the distinction between ‘impact’ and ‘need’ within needs statements
- Including illustrative examples, where helpful, to support participant understanding

These improvements informed the final draft of survey items and the development of ‘needs statements’ within each domain.

**Reimbursement of interview participants**

In line with ethical best practices, participant compensation was provided as a token of appreciation for their time, ensuring it was not of such value as to be considered coercive. Compensation amounts were determined with consideration of cost of living, purchasing power parity, and typical reimbursement rates for participation in similar health research within each country, and were finalised in the participant’s local currency using the exchange rate applicable at the time of transfer.
