## Supplemental Appendix 2 for "The Global Epilepsy Needs Study (GENS): A mixed-methods, multi-country exploration of the unmet psychosocial and everyday needs of people with epilepsy"

[SURVEY DICTIONARY - Participants will see the following words as link, when clicking their will see the definitions]

**Awareness programs**
Content that aims to increase what people know and improve attitudes linked to epilepsy and the people who live with it. Usually, it may talk about what epilepsy is, what actions can be taken to protect patients, changes to remove challenges, address unfair ideas or old beliefs around epilepsy.

**Co-morbidity**
Means having two or more diseases or health conditions at the same time. For example, a person might have both epilepsy and depression together, which can affect their health in different ways.

**Reasonable adjustments**
Changes employers or teachers make to remove or reduce a disadvantage related to someone's epilepsy. Examples at work: making changes to the workplace, changing someone's working patterns, finding a different way to do something or providing equipment, services or support (source: ACAS).

**Stigma**
When some people treat others unfairly because of something about them, like epilepsy. This happens because of unfair ideas or common, wrong beliefs and a lack of understanding.

**SUDEP**
Sudden Unexpected Death in Epilepsy (SUDEP) happens when a person with epilepsy dies suddenly, and no other cause of death is found. SUDEP is rare, but it's important to be aware of the danger because it may sometimes be preventable (source: NHS).

**Unmet needs**

Something important that people want very much but do not have. In the GENS survey, we want to understand those unmet needs of people with epilepsy, that can make life easier or better.

### What is GENS and why is it important?

The International Bureau for Epilepsy (IBE) is **starting a new research study**: The Global Epilepsy Needs Study (GENS).

IBE and our members (national epilepsy support groups) want to **better understand the needs of people with epilepsy**. GENS will help us to do this.

At the end of the study, IBE and its members will be able to **share the needs of people with epilepsy across the world**. IBE hopes this will lead to **improved support and services**.

A patient research company called **MediPaCe is working with IBE** to run this study.

### Why YOUR voice matters

We need to hear from as many people with epilepsy and caregivers as possible. Your own personal experience will give us a **clear picture of where you need most help**.

### How do I get involved: The GENS survey

The GENS survey is a major part of the research study.
It aims to explore the **unmet needs** of people with epilepsy in their daily lives.

In the survey, we will:

1. Ask some basic questions about **who you are**.
2. Ask questions about your **current everyday unmet needs**.
3. Give you a chance to share any **other unmet needs**.

### Who should answer the survey?

You should answer this survey if you are 18 years old or over:

- You are a person with epilepsy, **OR**
- You care for someone with epilepsy.

People **caring for someone** with epilepsy should:

- First: Complete three short screening questions.
- Then: Answer the survey **on behalf of the person with epilepsy**, which can be a child or adult.
- Later: Answer some brief questions about you and your needs as a caregiver.

Instructions are provided throughout the survey.

Our research will go deeper into the unmet needs of caregivers by doing interviews and focus groups at a later stage.

### How long will it take?

The survey should take around **30 minutes to complete**.

### Where can I find more information?

You can read more about GENS here:
https://www.ibe-epilepsy.org/initiatives/global-epilepsy-needs-study-gens/.

The study results will be presented on the IBE website (www.ibe-epilepsy.org) in early 2025 and will also be shared with all IBE members (160 national epilepsy associations across the world).

Please read the points below before you start the survey:

- MediPaCe will make sure **no one can connect answers** with survey respondents (anonymous).
- The (anonymous) results will be used by MediPaCe, IBE and involved national organizations to share the **collected results (aggregated)** nationally and globally.
- IBE will use the survey to learn about the **unmet needs** of people with epilepsy.
- IBE will **share the learnings of this study** in different ways. For example, in reports, journals, conferences, or websites for everyone to see.

If you have any questions about the survey or your data, please contact. Thank you for your interest in this survey.

Please choose from the two options below:

□ Yes, I agree (consent) with all the above.

□ No, I do not agree (consent) with all the above. [skip to no consent - thank you page]

// NEW WEBPAGE //

[Progress bar with % will be displayed at the top of the screen to visually indicate progress of the respondent]

### General questions about you

Q4. Please select from the following:

☐ I am a person with epilepsy

☐ I am a caregiver of a person with epilepsy

Q5. Please choose the country you live in.

*Select your country from the dropdown menu.*

[Dropdown menu with 'Other'.]

[Those that select 'Other' will see the following]

Please use the space below to indicate which country you live in:

Please select your age group.

☐ Under 18 years old [Skip to "Do not meet criteria" webpage]

☐ 18-24

☐ 25-34

☐ 35-44

☐ 45-54

☐ 55-64

☐ 65 or older

[NOTE ONLY FOR CAREGIVERS]

### General questions about the person you care for

It is time to answer from the perspective of the **adult or child with epilepsy you care for**. This will help us learn about their daily life and unmet needs.

Please select the age group of **the person you care for**.

☐ 0-4

☐ 5-11

☐ 12-17

☐ 18-24

☐ 25-34

☐ 35-44

☐ 45-54

☐ 55-64

☐ 65 or older

[END OF NOTE FOR CAREGIVERS]

[REMINDER NOTE ONLY FOR CAREGIVERS]
***REMINDER – Please answer on behalf of the person you care for.***
[REMINDER NOTE ONLY FOR CAREGIVERS]

Which best describes your gender?
☐ Female
☐ Male
☐ I prefer to describe as: ________
☐ I prefer not to say

[Only for those caring for someone 18 years or above] What is your relationship status?

☐ Single

☐ Married

☐ Living with partner

☐ Widowed

☐ Divorced/separated

Do you consider yourself part of any minority group?

☐ Yes

☐ No

☐ I prefer not to say

If yes, please use the space below to state which minority group(s) you belong to:
*(e.g., racial or ethnic minority, LGBTQ+, religious minority, person with a specific disability, etc.)*

_______________________________________________

What is your current work status?

☐ [only for caregivers] The person is a child, so this question does not apply

☐ Full-time employed

☐ Part-time employed

☐ Self-employed or freelance

☐ Unemployed or job seeking

☐ Student

☐ Retired

☐ Unable to work due to disability

☐ Other, please specify ____________________

What is the highest level of education you have completed?

☐ No formal education

☐ Early childhood education (e.g., nursery school, preschool)

☐ Primary/elementary school

☐ Secondary/high school completion or equivalent

☐ Some college, no degree

☐ Trade/technical/vocational training

☐ Standard undergraduate degree (e.g., bachelor's degree)

☐ Graduate or professional degree

☐ Other, please specify ______________________

Which type of area do you now live in?

☐ Urban (city)

☐ Suburban (outside a city, but not rural)

☐ Rural (countryside)

☐ Other, please specify ______________________

When were you diagnosed with epilepsy?

*Select the year when this happened.*

[Dropdown menu with years from 2024 till 1900]

How often do you experience seizures?

☐ **Multiple times a day**

☐ **Once a day**

☐ **A few times a week**

☐ **Once a week**

☐ **A few times a month**

☐ Once a month

☐ Less than once a month

☐ Never

☐ Other, please specify ______________________

On a typical day, how would you rate your level of dependence on others due to epilepsy?

☐ Fully independent, no help required

☐ Sometimes require help with difficult tasks

☐ **Often need support with everyday tasks**

☐ **Dependent on others for most daily tasks**

☐ **Fully dependent, requiring constant support**

☐ Other, please specify ______________________

Please rate the following aspects of your **current** health from very poor to excellent.
*Please note, the health aspects below might be connected or might not apply to you.*

| | Very poor | Poor | Fair | Good | Excellent | Not applicable |
|---|---|---|---|---|---|---|
| **General health** (e.g., how you feel overall, energy levels, fitness) | | | | | | |
| **Epilepsy health** (e.g., seizure control, medication management, side effects) | | | | | | |
| **Thinking and memory** (e.g., remembering things, ability to concentrate, problem-solving) | | | | | | |
| **Behaviors** (e.g., in rare and complex epilepsies, developmental delays, hyperactivity, impulsivity, mood disorders) | | | | | | |
| **Overall wellbeing** (e.g., mood, stress levels, enjoyment of life) | | | | | | |
| **Mental health** (e.g., anxiety, depression, bipolar, psychosis) | | | | | | |

Do you think of your epilepsy as a disability?

☐ No, I don’t

☐ I’m not sure

☐ Yes, I do

☐ Prefer not to say

Do you feel epilepsy is thought of as a disability where you live?

☐ No, it is not

☐ Some people do, some people don’t

☐ Yes, it is

☐ I’m not sure

☐ Prefer not to say

Who have you told about your epilepsy?

*Select all that apply*

☐ My partner/spouse

☐ My family members (like parents, guardians, siblings)

☐ My relatives (like grandparents, aunts, uncles, cousins)

☐ My friends or peers

☐ School / preschool / training center staff (like teachers or nurses) – where I attend as a student

☐ My employer

☐ My co-workers

☐ I haven’t told anyone

☐ Other, please specify _____________________

// NEW WEBPAGE //

[Entire section to be displayed for respondent]

### Your life with epilepsy

We will now ask about your unmet needs. All questions relate to **your daily life with epilepsy currently (during the last three months).** If you have questions about the survey, contact:

[REMINDER NOTE ONLY FOR CAREGIVERS]

***REMINDER – Please answer on behalf of the person you care for.***

[REMINDER NOTE ONLY FOR CAREGIVERS]

// NEW WEBPAGE //

[Entire section to be displayed for respondent]

### Section 1: Knowledge and advice

This section is about knowledge and access to advice about epilepsy.

Think about your **current daily life** with epilepsy. Select all the statements that you consider are **unmet needs** for you.

*If the needs listed below do not relate to you, please select the last option to go straight to the next section.*

[List to be randomized]

- ❑ I need access to more high-quality information about epilepsy (e.g., seizure control, triggers, treatment options, side effects)
- ❑ My family and those who care for me need to learn more about epilepsy (e.g., tips to help me stay safe, seizure first-aid, seizure control)
- ❑ My local community needs to learn more about epilepsy (e.g., seizure first-aid, why seizures happen, famous people that have it)
- ❑ Doctors and nurses need to know more about peoples' wider needs linked to epilepsy (e.g., emotions and mental health needs of people with epilepsy at different stages of life)
- ❑ I need advice on how to have useful conversations with doctors and nurses
- ❑ My local community needs simple and easy to access content about providing help during a seizure

- ❑ I want advice on how to tell key people in my life about my epilepsy (e.g., about my diagnosis, how it affects my life)
- ❑ My local community needs better information to get rid of old, incorrect beliefs about epilepsy
- ❑ I do not have any of the above needs

We now want to learn what will most improve the quality of your daily life. We want to know which unmet needs you believe should be addressed **more urgently than others**.

The table below lists your selected unmet needs. Please rate these from ***Not a priority*** to ***Urgent priority***.

| | Not a priority | Low priority | Medium priority | High priority | Urgent priority |
|---|---|---|---|---|---|
| [Statements auto filled from above selections] | o | o | O | o | O |

OPTIONAL

Have we **missed any unmet needs** related to knowledge and access to information about epilepsy? Please let us know by using the space below.

// NEW WEBPAGE //

[Entire section to be displayed for respondent]

### Section 2: Safety and survival

This section is about understanding when you feel at risk and how you could be made to feel safer.

Think about your **current daily life** with epilepsy. Select all the statements that you consider are **unmet needs** for you.

*If the needs listed below do not relate to you, please select the last option to go straight to the next section.*

[List to be randomized]

- ❑ I need to feel safe from the risk of injuries linked to seizures in a public space
- ❑ There is a need to reduce the risks of life-changing injuries or dying from epilepsy (e.g., SUDEP)
- ❑ I need to feel safe when traveling to different places (e.g., visiting family, holidays)
- ❑ There needs to be advice on how to cope with changes in weather that may impact people with epilepsy (e.g., extreme heatwaves)
- ❑ Basic seizure first-aid should be more widely known in society to help keep me safe
- ❑ My country needs to recognize and defend the human rights of people with epilepsy (e.g., through laws, nationwide campaigns, etc.)
- ❑ I need to feel safe from being physically hurt by others because of my epilepsy
- ❑ I need to feel safe from being emotionally hurt by others because of my epilepsy
- ❑ I need to feel sure that I will get the right help or support during an emergency (e.g., during conflicts, natural disasters, pandemic and other crises)
- ❑ I need to feel sure that I will get the right help or support during an epilepsy emergency (e.g., prolonged seizure, serious injuries suffered during a seizure in a public place)
- ❑ I do not have any of the above needs

We now want to learn what will most improve the quality of your daily life. We want to know which unmet needs you believe should be addressed **more urgently than others**.

The table below lists your selected unmet needs. Please rate these from ***Not a priority*** to ***Urgent priority***.

| | Not a priority | Low priority | Medium priority | High priority | Urgent priority |
|---|---|---|---|---|---|
| [Statements auto filled from above selections] | o | O | o | o | O |

OPTIONAL

Have we **missed any unmet needs** related to your safety and survival? Please let us know by using the space below.

// NEW WEBPAGE //

[Entire section to be displayed for respondent]

### Section 3: Healthcare and wellbeing

This section is about healthcare services, treatments, and wellbeing.

Think about your **current daily life** with epilepsy. Select all the statements that you consider are **unmet needs** for you.

*If the needs listed below do not relate to you, please select the last option to go straight to the next section.*

[List to be randomized]

- ❑ There is a need for easier access to healthcare services or doctors and nurses who treat epilepsy
- ❑ There is a need for faster access to the right treatment (e.g., medicine, procedures)
- ❑ I need to get health care that addresses my own specific needs (e.g., having a personal care plan)
- ❑ I need ongoing access to epilepsy treatment (e.g., seizure medicines)
- ❑ I need help dealing with the side effects of my epilepsy medicine
- ❑ I need help taking care of my other health problems along with my epilepsy (e.g., co-morbidity)
- ❑ There is a need for easier access to rare or complex epilepsy services (e.g., access to specialists and special care, help with treatment decisions)
- ❑ I need easier access to good medical insurance coverage to afford epilepsy treatment
- ❑ I do not have any of the above needs

We now want to learn what will most improve the quality of your daily life. We want to know which unmet needs you believe should be addressed **more urgently than others.**

The table below lists your selected unmet needs. Please rate these from ***Not a priority*** to ***Urgent priority***.

| | Not a priority | Low priority | Medium priority | High priority | Urgent priority |
|---|---|---|---|---|---|
| [Statements auto filled from above selections] | O | o | o | o | O |

OPTIONAL

Have we **missed any unmet needs** related to your health and wellbeing? Please let us know by using the space below.

// NEW WEBPAGE //

[Entire section to be displayed for respondent]

### Section 4: Learning and education

This section is about learning and the educational setting.

Are you **currently attending or planning to attend** formal education? E.g., preschool, nursery, school, college, training, university, etc.

- ☐ Yes
- ☐ No, skip to the next section [Skip to section 5]

Think about your **current daily life** with epilepsy. Select all the statements that you consider are **unmet needs** for you.

*If the needs listed below do not relate to you, please select the last option to go straight to the next section.*

[List to be randomized]

- ❑ My teachers need to learn about epilepsy so that they don't have unfair or wrong beliefs about it
- ❑ I need to feel free from stigma and negative attitudes from fellow students
- ❑ During my studies or training, I would like to have access to reasonable adjustments (e.g., support to avoid triggers, rest space, extra time, etc.)
- ❑ All staff need to learn about seizures and how to manage them
- ❑ I would like to access different ways of learning that can help with my focus and memory problems (e.g., visual learning, recorded lessons, assorted color paper, etc.)
- ❑ I would like to feel more included in the learning setting (e.g., teachers letting all students know that differences are normal)
- ❑ There needs to be laws and government programs that make sure learning for people with epilepsy is easier

- ❑ I do not have any of the above needs

We now want to learn what will most improve the quality of your daily life. We want to know which unmet needs you believe should be addressed **more urgently than others**.

The table below lists your selected unmet needs. Please rate these from ***Not a priority*** to ***Urgent priority***.

| | Not a priority | Low priority | Medium priority | High priority | Urgent priority |
|---|---|---|---|---|---|
| [Statements auto filled from above selections] | O | O | o | o | O |

OPTIONAL

Have we **missed any unmet needs** related to your learning and education? Please let us know by using the space below.

---

// NEW WEBPAGE //

[ONLY FOR THOSE THAT SAID THEY ARE EMPLOYED, SELF-EMPLOYED OR JOBSEEKING
Entire section to be displayed for respondent]

### Section 5: Work and income

This section is about job seeking, the workplace and having an income.

Think about your **current daily life** with epilepsy. Select all the statements that you consider are **unmet needs** for you.

*If the needs listed below do not relate to you, please select the last option to go straight to the next section.*

[List to be randomized]

- ❑ I would like access to reasonable adjustments specific to me (e.g., flexible working hours, no bright lights or screens, flexibility for medical appointments)
- ❑ There need to be government programs that remove barriers people with epilepsy face when trying to find a job
- ❑ I need more support to make sure epilepsy does not affect my job security and career progress (e.g., dealing with seizures or going to health checks)
- ❑ I need to feel more included in the workplace and be able to build relationships
- ❑ My work colleagues need training on what epilepsy is and how to deal with seizures
- ❑ I need to feel free from stigma in the workplace and negative attitudes of colleagues
- ❑ I would like to have a stable income and learn how to manage my money so that I can better provide for myself
- ❑ There is a need for stronger laws to protect the working rights of people with epilepsy
- ❑ There is a need for policies to protect people with epilepsy from being unfairly treated and discrimination in the workplace
- ❑ I do not have any of the above needs

We now want to learn what will most improve the quality of your daily life. We want to know which unmet needs you believe should be addressed **more urgently than others**.

The table below lists your selected unmet needs. Please rate these from ***Not a priority*** to ***Urgent priority***.

| | Not a priority | Low priority | Medium priority | High priority | Urgent priority |
|---|---|---|---|---|---|
| [Statements auto filled from above selections] | o | o | o | O | O |

OPTIONAL

Have we **missed any unmet needs** related to your work and income? Please let us know by using the space below.

// NEW WEBPAGE //
[Entire section to be displayed for respondent.]

### Section 6: Transport and driving

This section is about safe transport and driving rules in your country.

Which of the transport options below do people with epilepsy have in your country?
*Please select all that apply.* [List to be randomized]

☐ Specialized transport services (i.e., specifically for people with more transport needs or people with epilepsy)

☐ Public transport with general adjustments (e.g., special safety measures for people with disabilities or epilepsy, free public transport)

☐ Private (paid for) services with trained drivers (e.g., where drivers are trained in seizure first-aid)

☐ Only standard public transport options without any adjustments (i.e., no special safety measures for people with disabilities or epilepsy)

☐ I don't know

☐ Other, please specify _____________________

Think about your **current daily life** with epilepsy. Select all the statements that you consider are **unmet needs** for you.
*If the needs listed below do not relate to you, please select the last option to go straight to the next section.*
[List to be randomized]

❑ I would like public transport to be easier for me to access (e.g., through special fares, lifts, ramps etc.)

- ❑ There is a need for more support and plans in case people have a seizure on public transport
- ❑ Public transport workers need training to support people with epilepsy (e.g., spotting seizures, giving first aid, etc.)
- ❑ There needs to be safe travel options that people with epilepsy can afford
- ❑ Driving restrictions on people with epilepsy in my country limit my independence
- ❑ I need more support to travel to work and earn a living
- ❑ I do not have any of the above needs

We now want to learn what will most improve the quality of your daily life. We want to know which unmet needs you believe should be addressed **more urgently than others**.

The table below lists your selected unmet needs. Please rate these from ***Not a priority*** to ***Urgent priority***.

| | Not a priority | Low priority | Medium priority | High priority | Urgent priority |
|---|---|---|---|---|---|
| [Statements auto filled from above selections] | o | o | o | o | o |

OPTIONAL
Have we **missed any unmet needs** related to safe transport and driving? Please let us know by using the space below.

// NEW WEBPAGE //

[Entire section to be displayed for respondent]

### Section 7: Community and household

This section is about your relationship with others, feeling part of your community and life at home.

Are you aware of any epilepsy support groups you could access?

☐ Yes, I am aware and have attended

☐ Yes, I am aware but have **not** attended [Skip next question]

☐ No, I am not aware but am interested [Skip next question]

☐ No, I am not aware and am **not** interested [Skip next question]

[for those that answered 'Yes, I am aware and have attended']

If you have attended an epilepsy support group, how would you rate your experience?

| **Very negative** I felt alone and disconnected from the group | **Negative** I did not feel a strong sense of belonging | **Neutral** I didn't feel strongly one way or the other | **Positive** I felt welcomed and included | **Very positive** I felt a strong sense of belonging and connection |
|---|---|---|---|---|

Think about your **current daily life** with epilepsy. Select all the statements that you consider are **unmet needs** for you.

*If the needs listed below do not relate to you, please select the last option to go straight to the next section.*

[List to be randomized]

❑ I would like to feel welcomed when connecting with others in my community

❑ Social events need to avoid epilepsy triggers and make sure people who can give seizure first-aid are available

- ❑ I need advice on how, who and when to confidently tell others about my epilepsy
- ❑ I need to feel well enough to help with household chores
- ❑ I would like to have opportunities to find a life partner, wife, or husband
- ❑ My country needs to protect the rights of people with epilepsy
- ❑ I need better legal support when being unfairly treated or harmed because of my epilepsy
- ❑ I do not have any of the above needs

We now want to learn what will most improve the quality of your daily life. We want to know which unmet needs you believe should be addressed **more urgently than others**.

The table below lists your selected unmet needs. Please rate these from ***Not a priority*** to ***Urgent priority***.

| | Not a priority | Low priority | Medium priority | High priority | Urgent priority |
|---|---|---|---|---|---|
| [Statements auto filled from above selections] | o | o | o | o | o |

OPTIONAL
Have we **missed any unmet needs** related to your relationship with others, feeling part of your community and life at home? Please let us know by using the space below.

// NEW WEBPAGE //

[Entire section to be displayed for respondent]

### Section 8: Mental health and wellbeing

This section is about mental health and wellbeing.

Think about your **current daily life** with epilepsy. Select all the statements that you consider are **unmet needs** for you.

*If the needs listed below do not relate to you, please select the last option to go straight to the next section.*

[List to be randomized]

- ❑ I need access to mental health care that is specific for people with epilepsy
- ❑ There needs to be better planning between mental health care and epilepsy care
- ❑ I need help with keeping my focus and remembering things in my daily life
- ❑ There is a need for affordable mental health services specific to people with epilepsy
- ❑ I want to learn how to be aware of, and cope with, my emotions
- ❑ I need support to better cope with feeling isolated and lonely
- ❑ I need to decrease my stress levels or learn to better manage it
- ❑ I would like to learn about mindfulness and other self-care practices
- ❑ I do not have any of the above needs

We now want to learn what will most improve the quality of your daily life. We want to know which unmet needs you believe should be addressed **more urgently than others**.

The table below lists your selected unmet needs. Please rate these from ***Not a priority*** to ***Urgent priority***.

| | Not a priority | Low priority | Medium priority | High priority | Urgent priority |
|---|---|---|---|---|---|
| [Statements auto filled from above selections] | o | o | o | O | o |

OPTIONAL

Have we **missed any unmet needs** related to your mental health and wellbeing? Please let us know by using the space below.

// NEW WEBPAGE //

[Entire section to be displayed for respondent.]

### Section 9: Sexual and reproductive health

This section is about intimacy in your life and having children.

Think about your **current daily life** with epilepsy. Select all the statements that you consider are **unmet needs** for you.

*If the needs listed below do not relate to you, please select the last option to go straight to the next section.*

[List to be randomized]

- ❑ I need access to doctors and nurses specialized in sexual health and reproduction and epilepsy
- ❑ I need advice on the effect of epilepsy medicine on my sexual health and plans to become a parent
- ❑ I want to learn how to manage stigma, linked to epilepsy, in intimate relationships
- ❑ There needs to be simple information for people with epilepsy who want to become parents (e.g., risks from epilepsy medicines)
- ❑ I would like expert help to make the best choice about becoming a parent
- ❑ I do not have any of the above needs

We now want to learn what will most improve the quality of your daily life. We want to know which unmet needs you believe should be addressed **more urgently than others**.

The table below lists your selected unmet needs. Please rate these from ***Not a priority*** to ***Urgent priority***.

| | Not a priority | Low priority | Medium priority | High priority | Urgent priority |
|---|---|---|---|---|---|

| | | | | | |
|---|---|---|---|---|---|
| [Statements auto filled from above selections] | o | o | o | O | o |

OPTIONAL

Have we **missed any unmet needs** related to intimacy in your life and having children? Please let us know by using the space below.

// NEW WEBPAGE //

[Entire section to be displayed for respondent]

### Section 10: Achieving your goals

This section is about quality of life and reaching your full potential in life.

Think about your **current daily life** with epilepsy. Select all the statements that you consider are **unmet needs** for you.

*If the needs listed below do not relate to you, please select the last option to go straight to the next section.*

[List to be randomized]

- ❑ I feel I need more time and support than others to be able to reach my goals and ambitions
- ❑ There is a need for programs and support to improve confidence and achieve personal goals and dreams
- ❑ I need reliable access to epilepsy care for my overall happiness and daily life to improve
- ❑ I need better housing or accommodation
- ❑ I need support to manage my fear and anxiety about seizures
- ❑ I would like to learn more about self-care to support more independent living
- ❑ I need to learn how to identify, handle and/or protect myself from stigma around epilepsy that exists in my country
- ❑ I do not have any of the above needs

We now want to learn what will most improve the quality of your daily life. We want to know which unmet needs you believe should be addressed **more urgently than others**.

The table below lists your selected unmet needs. Please rate these from ***Not a priority*** to ***Urgent priority***.

| | Not a priority | Low priority | Medium priority | High priority | Urgent priority |
|---|---|---|---|---|---|
| [Statements auto filled from above selections] | O | o | O | o | o |

OPTIONAL

Have we **missed any unmet needs** related to your quality of life and reaching your full potential in life? Please let us know by using the space below.

---

[START Caregivers only]

### Caring for someone with epilepsy: General questions about you

**REMINDER – Now please answer for yourself.**

Which best describes your gender?

☐ Female

☐ Male

☐ I prefer to describe as: ________

☐ I prefer not to say

What is your current work status?

☐ Full-time employed

☐ Part-time employed

☐ Self-employed or freelance

☐ Unemployed or job seeking

☐ Student

☐ Retired

☐ Unable to work due to disability

☐ Other, please specify ______________________

What is the highest level of education you have completed?

☐ No formal education

☐ Primary/elementary school

☐ Secondary/high school completion or equivalent

☐ Some college, no degree

☐ Trade/technical/vocational training

☐ Standard undergraduate degree (e.g., bachelor's degree)

☐ Graduate or professional degree

☐ Other, please specify ______________________

Please rate the following aspects of your **current** health from very poor to excellent.

*Please note, the health aspects below might be connected or might not apply to you.*

| | Very poor | Poor | Fair | Good | Excellent | Not applicable |
|---|---|---|---|---|---|---|
| **General health**<br>(e.g., how you feel overall, energy levels, fitness) | | | | | | |
| **Overall wellbeing**<br>(e.g., mood, stress levels, enjoyment of life) | | | | | | |

Do you live with the person with epilepsy you care for?

☐ Yes

☐ No

What is your connection to the person with epilepsy that you care for?

*Select your connection from the list below.*

☐ Parent

☐ Husband or wife

☐ Partner

☐ Grandchild

☐ Grandparent

☐ Child

☐ Brother or sister

☐ Friend/Family friend

☐ Other relative

☐ Other, please specify ____________________

☐ I'd prefer not to say

Please think about the amount of care you provide for the person with epilepsy. Which of the following best describes your situation?

☐ I am the primary caregiver.

☐ Someone else is the primary caregiver.

☐ I share caregiving responsibilities equally with someone else.

☐ Other, please specify ____________________

☐ I'd prefer not to say

// NEW WEBPAGE //

[Entire section to be displayed for respondent]

### **Caring for someone with epilepsy:** Your experiences

Your needs as a caregiver matter. In this section, we will ask you about what might improve **your** daily life.

Think about your **current daily life** with epilepsy. Select all statements that you consider are **unmet needs for you** as a caregiver**.**

[List to be randomized]

- ❑ I need money to cover the costs related to caring for someone (e.g., medicines, reduced work hours)
- ❑ There needs to be rules and laws that support caregivers (e.g., adjustments in the workplace, time to rest for caregivers)
- ❑ I need training for caregivers on epilepsy management
- ❑ There should be awareness programs to tackle stigma
- ❑ I need help with the problems the person I care for has with remembering things, focusing, and solving problems (known as cognition)
- ❑ I need help with the issues the person I care for has with behavior
- ❑ There needs to be inclusive social activities for caregivers and those they care for
- ❑ I need help balancing my own goals (ambitions) and my caregiving role
- ❑ I do not have any of the above needs

We now want to learn what will **most improve** the quality of your daily life. We want to know which of your unmet needs you believe should be addressed more urgently than others.

The table below lists your selected unmet needs. Please rate these from ***Not a priority*** to ***Urgent priority***.

| | Not a priority | Low priority | Medium priority | High priority | Urgent priority |
|---|---|---|---|---|---|
| [Statements auto filled from above selections] | O | O | o | o | o |

OPTIONAL

Have we **missed any unmet needs** related to your life as a caregiver? Please let us know by using the space below.

---

[END Caregivers only]

// NEW WEBPAGE //

Thank you, you are almost there! **And finally…**

We really want to hear what you think. If there's something about living with epilepsy that you haven't told us yet, please let us know. What you say helps us learn more about real-life unmet needs and concerns people with epilepsy have in their daily life.

| [word limit of 300 words] |
|---|

// NEW WEBPAGE //
[For respondents that selected 'Yes, I consent to take part in the survey' and completed the survey]

**Thanks a lot for filling out our survey.**

What you've shared with us is important because it helps us understand the different ways epilepsy affects people. Your input is key to helping us make things better for everyone with epilepsy everywhere.

// NEW WEBPAGE //
[**For people that selected 'No, I don't consent to take part in the survey'**]

**We appreciate your interest and thank you for considering participating in our survey.**

[Optional question to capture reasons for non-responders]
If you don't mind sharing, could you tell us why you chose not to take part? This will help us improve our studies in the future. Thank you.
*Select all that apply.*

☐ I don’t have enough time right now.

☐ I’m not comfortable sharing personal information about my epilepsy.

☐ I don’t think the survey is relevant to my experiences.

☐ I need help to complete the survey.

☐ I have concerns about my privacy.

☐ Other, please specify ____________________

// NEW WEBPAGE //

[**For people that do not meet the survey’s selection criteria**]

We appreciate your interest and thank you for considering participating in our survey. Stay in touch with your local epilepsy patient association for other opportunities to participate in research.
