## Supplementary figures and images for "The Global Epilepsy Needs Study (GENS): A mixed-methods, multi-country exploration of the unmet psychosocial and everyday needs of people with epilepsy"

### Supplemental Figure 1

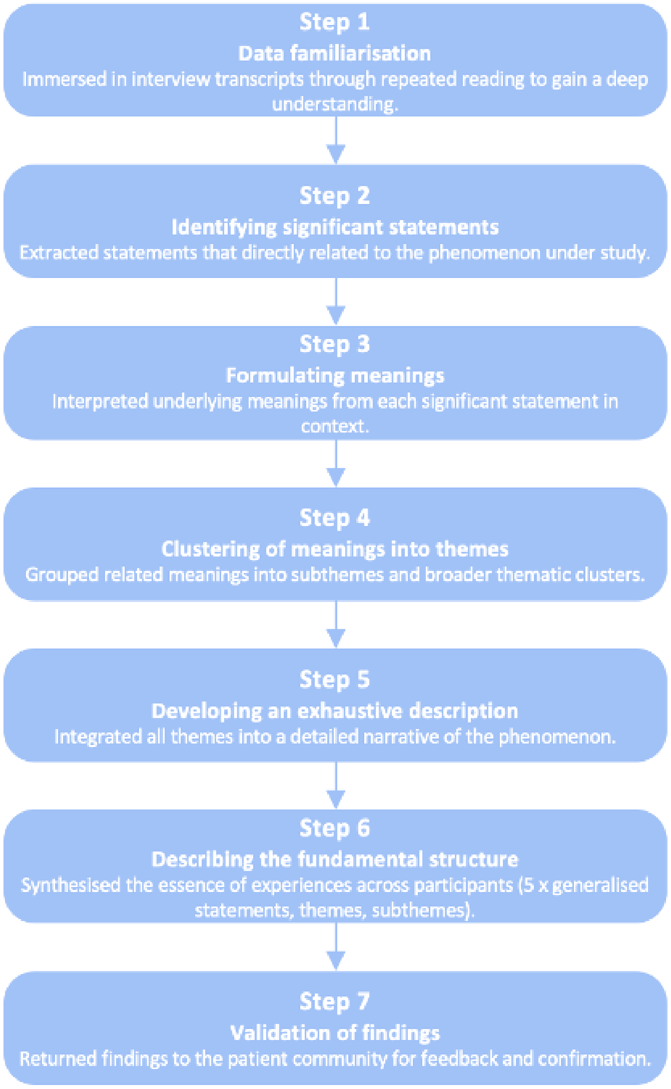


Figure: Steps in Colaizzi's phenomenological data analysis method.
