## Supplemental Table 1 for "The Global Epilepsy Needs Study (GENS): A mixed-methods, multi-country exploration of the unmet psychosocial and everyday needs of people with epilepsy"

| | Qual | | | Quant | |
|---|---|---|---|---|---|
| **Generalised theme** | **Subtheme** | **Codes** | **Supporting quote(s)** | **Statistic** | **Integration comment** |
| **1. Managing uncertainty and redefining daily life** | Physical risk inherent to epilepsy | Living with everyday physical dangers | "I've been recovering from a knee injury caused by a seizure I had, where my leg got stuck on the couch at home. It caused a medial collateral ligament strain."<br>Brazil, Male, Participant 1<br><br>"I also bought an induction oven...It's for my safety because I love cooking and baking...if I feel like I'm having an off day, I just put things in a slow cooker and I let it cook all day. See I think ahead like that and... the induction oven will turn off [in the event that the participant has a seizure and is unable to turn off the oven themselves]."<br>Canada, Rural (female), Participant 2 | **Divergence:** Female respondents were significantly more likely than male respondents to report a need to reduce the risks of life-changing injuries or dying from epilepsy (e.g., SUDEP) (47% vs. 40%; OR = 1.4, p < 0.0001). | Although survey data showed that female respondents were more likely than male respondents to express a need to reduce the risks of serious harm or death from epilepsy (e.g. SUDEP), this emphasis was not reflected in the qualitative interviews. While seizure-related injuries and more immediate, tangible risks (such as falling during a seizure) were mentioned, these references were largely descriptive and not accompanied by an explicit desire to reduce such risks. Instead, they were treated as an expected part of living with epilepsy, prompting practical adjustments aimed at maintaining autonomy and keeping daily life running smoothly, such as installing induction ovens or reorganising household routines.SUDEP was mentioned only once and was quickly framed as something to be aware of but not actively worried about, given the perceived lack of control over its occurrence. Overall, while physical risks were acknowledged, they were not a central focus, with everyday, visible hazards shaping how risk was perceived and managed. |
| | Emotional uncertainty and psychological strain | Living with the unpredictability of seizures | "[Society] don't take into account how bad we feel, how painful it is to have a seizure, the fear we have. I live in fear."<br>Argentina, Male (urban/suburban), Participant 2<br><br>"'Occasionally, I feel nervousness, anxiety, and restlessness. Epileptic seizures frustrate me because they happen daily. I don't currently have therapy for my anxiety, but I feel that I need it."<br>Bosnia and Herzegovina, Rural, Participant 4<br><br>"I need stability. Financial and career stability, in terms of work, is crucial for me. A lack of stability creates discomfort, which can even trigger seizures...companies don't understand. They say 'we agree' but then...they don't."<br>Brazil, Rural, Participant 3 | **Complementary:** People living in rural areas were significantly more likely to report needing support to manage fears and anxiety about seizures compared to those in suburban or urban areas (46% vs. 43% vs. 43%; OR = 2.0, p<0.0001). | Survey findings show that people in rural areas were more likely to report needing support to manage fears and anxiety about seizures. Qualitative data demonstrated that fear and emotional strain, particularly due to the unpredictability of seizures, were common across all locations, not uniquely intensified in rural areas. However, the few explicit requests for support captured in the qualitative data came only from rural participants. This limited alignment suggests a somewhat complementary relationship, where qualitative insights provide partial support to the survey trend by illustrating how and where these support needs are voiced, rather than reinforcing a broader rural-urban difference in emotional impact. |

| | | | | | |
|---|---|---|---|---|---|
| | | Emotional impact of stigma | “Every time I heard this disease referred to as "sheep madness" in our area, I couldn't help but have wild thoughts, feeling as if there was something wrong with my mental state, and I was filled with a sense of shame...The thought of having a seizure in public and being seen by others makes me feel extremely embarrassed and afraid, to the point where I'm hesitant to even leave the house”<br>China, Older, Participant 5<br><br>"The stigma surrounding epilepsy...is still very strong, so many are afraid to openly talk about their condition. They try to act as if they don’t have epilepsy, which is mentally and emotionally exhausting.”<br>Bosnia and Herzegovina, Male, Participant 5 | No corresponding quantitative item. | No integration required. |
| | | Emotional distress beyond seizures | "I spent 2 years asking for [virtual cognitive rehabilitation]... The rehabilitation was only in the capital, 160 km away. The insurance, if I did it in in person, they would cover it, but not virtually... I went [to the capital] to request it...they printed it there for me, I couldn't believe it, I started crying on the spot"<br>Argentina, Rural, Participant 3<br><br>"We live in a rural town, and some epilepsy medications are not available at the local hospital. For instance, the medication I take...is an imported drug...It would be great if the community health centers could manage patients with epilepsy, understand their needs, and even purchase some of the medications that are not available at the local hospital on a centralized basis...The imported medication I take has to be paid for entirely out of pocket, and it's a significant expense every year. This financial burden causes me a lot of psychological stress."<br>China, Older, Participant 5 | **Complementary:** Respondents who self-identified as having a disability were significantly more likely to report a need for reliable access to epilepsy care in order to improve their overall happiness and daily life than those who did not self-identify as disabled (45% vs 32%; OR = 2.0, p<0.0001). | Survey data indicated that people who self-identified as having a disability placed greater emphasis on the need for reliable access to epilepsy care to support their overall happiness and daily life. Qualitative accounts complement this by illustrating how inconsistent or inaccessible care disrupts daily routines, imposes emotional and logistical burdens, and contributes to broader feelings of distress or neglect. Participants described long travel distances to receive treatment, medication shortages, delays in accessing services, and the stress this creates for both individuals and caregivers. |

| | | | | | |
|---|---|---|---|---|---|
| | | Emotional distress beyond seizures | “Before the epilepsy, I thought everything was possible. After epilepsy, I have realized that there are many things that are impossible.”<br>Argentina, Older (Male), Participant 1<br><br>"I think [epilepsy] makes you a bit angrier because your patience for things that annoy you becomes smaller."<br>Uganda, Older (male), Participant 3<br><br>"After getting epilepsy, he is even less likely to talk about his inner feelings.”<br>China, Caregiver (non-complex epilepsy, male PWE), Participant 3<br><br>“At the beginning of my diagnosis, I was very depressed. I was worried about it. I felt like I couldn't do well no matter how I did it...i really felt like the world had collapsed”.<br>South Korea, Rural (female), Participant 5<br><br>"After being diagnosed with epilepsy, it has had a significant impact on my psychology and emotions. When I first found out about my condition, it felt as if a heavy boulder had been placed on my heart, creating an immense psychological burden”<br>China, Older (female), Paricipant<br><br>“conversation is important, psychological support is more important than someone helping me if I had a seizure."<br>Croatia, Rural (female), Participant 3 | **Complementary:** Female respondents were more likely than male respondents to report: wanting to learn how to be aware of, and cope with, their emotions (48% vs. 39%; OR = 1.4), support to better cope with feeling isolated and lonely (39% vs. 34%; OR = 1.3), and decrease their stress levels or learn to better manage them (53% vs. 44%; OR = 1.4, p<0.0001). | Quantitative findings revealed that female respondents were significantly more likely than male respondents to express a need for emotional and psychosocial support across three areas: emotional awareness and coping; managing isolation and loneliness ; and stress reduction. Qualitative data complemented the first two areas; while the emotional impact of epilepsy was evident across genders, female participants were more likely to speak about the emotional and psychological toll of living with epilepsy or caregiving, often describing persistent worry, exhaustion, and the need for support. Emotional impacts were also commonly described by male participants, but often in relation to a shift in how they see themselves e.g., changes in confidence, independence, and cognitive function, and sometimes mood instability. They were however less likely to discuss seeking psychological help. When it came to stress management, a central concern for both men and women, participants across genders described concrete strategies to cope with stress (see supporting quotes in row 11). This suggests a complementary finding: while quantitative data showed women were more likely to express a need for stress reduction support, qualitative findings indicate that managing stress is a shared priority regardless of gender. |

| | | | | | |
|---|---|---|---|---|---|
| | | Emotional distress beyond seizures | "I believe that regular support from my family, combined with learning how to manage stress and frustration, would help improve my mental health."<br>Bosnia, Rural, Participant 4<br><br>"Reducing stress would help me the most. For example, when I don't have many school obligations, I feel much better. Homework, studying, and other tasks create significant stress, and stress, as psychologists often say, acts as a "silent killer" that negatively impacts mental and overall health. There should be ways for people with epilepsy to better manage stress, as it greatly affects their condition and quality of life."<br>Bosnia and Herzegovina, Male (urban/suburban), Participant 5<br><br>"One of my main triggers is stress, so that's why I get into the volunteering. I just love the better mental health aspects of it. Watching that stress, I do yoga right before I go to bed. Just a a simple one. That's more flexibility than actually working out and it just relaxes the muscles help you release stress right before you go to bed, and that helps me sleep a lot better."<br>US, Rural, Participant 3<br><br>"[To take care of their epilepsy] I try to get enough sleep to reduce stress...It all comes down to stress. I like to meditate"<br>Croatia, Male (urban/suburban), Participant 2 | **Divergence:** Respondents from rural areas were significantly more likely than those from suburban or urban areas to report that they need to decrease their stress levels or learn to better manage them (57% vs. 50% vs. 47%; OR = 1.5, p<0.0001). | Quantitative data highlighted a stronger reported need among rural respondents to reduce or better manage stress, but this location-based difference was not clearly reflected in the qualitative findings. Participants across rural, urban, and suburban settings consistently identified stress as a major challenge, linking it to seizure control, emotional wellbeing, and daily functioning. They described various coping strategies, such as family support, volunteering, and relaxation techniques, but did not frame their experiences in relation to geographic context. |
| | | Emotional distress beyond seizures | "I must be very careful when leaving home because I have been hit by three cars. Because when I have a seizure I have no control over anything.... I can be walking or I can stop in the middle of the road or I can be lucky that nothing happens to me. I fell and broke, no I got stuck with a shard of glass here (interviewee indicates back of head), I mean I went three fingers. The things that for others are very banal, aren't for me. Yesterday I fell. This is going to last me three days...If I'm really anxious I don't leave the house anymore. If I have a seizure in the morning I no longer leave the house that day. Because it's irresponsible. I don't play that game."<br>Spain, Male, Participant 4<br><br>"there was other times where I did have a seizure on my bike and I had a bike accident, messed up my knee a little bit"<br>Canada, Male, Participant 5<br><br>"I will proactively search for information on actions, preparations or follow-up works as well as first-aid measures to prevent me from injury if I have a seizure."<br>China, Male, Participant 2 | **Divergence:** Female respondents were significantly more likely than male respondents to report a need to feel safe from the risk of injuries linked to seizures in a public space (49% vs. 42%; OR = 1,4, p < 0.0001). | Survey results indicated that women were more likely than men to express a need to feel safe from injury during seizures in public spaces. However, this gendered concern was not clearly reflected in the qualitative data. Men offered more detailed accounts of physical harm linked to seizures, including injuries from falls, bike accidents, and other incidents, some of which occurred in public, with additional examples noted at home. Female participants rarely, if at all, explicitly described seizure-related injuries. While participants of all genders expressed concern about having seizures in public, these concerns were generally tied to stigma and social discomfort rather than fear of physical harm. This divergence suggests that women's heightened reporting of injury-related safety concerns in the survey may reflect a different way of internalising or anticipating vulnerability in public settings, one that was not spontaneously articulated in qualitative interviews. |

| | | | | | |
|---|---|---|---|---|---|
| | Barriers to everyday roles and recreation | Limited freedom of travel | "I'm a little afraid of going somewhere alone, and I think it'll be okay if it's in Korea, but actually, I really wanted to go to Japan in the winter, but now that I think about it, I'm a little scared. Because if I fall down on the street, can these people help me? In Korea, I get in touch and the people around me can come and help me, but if it's overseas, I don't think that's possible. So I thought about whether I can go on this trip or not."<br>South Korea, Rural (female), Participant 5<br><br>"my safety isn't a concern of mine anymore before, like even when I got my permit, I wouldn't drive throughout college because I was so scared of it, but the more I've gone through life. And the longer I've been seizure free, the more comfortable I've been.""<br>US, Male, Participant 4<br><br>"I need to be well-rested and I don't travel alone. On longer trips, my wife and I take turns so that we don't drive for long periods at a time. I can only drive for three hours at a time, and it's becoming increasingly difficult."<br>Croatia, Male, Participant 2<br><br>"Actually, I don't drive, so I only use public transportation. I was a little worried about having a seizure while using public transportation at first."<br>South Korea, Male, Participant 1" | **Complementary:** Female respondents were significantly more likely than male respondents to report a need to to feel safe when traveling to different places (e.g., visiting family, holidays) (46% vs. 38%; OR = 1.4, $p < 0.0001$). | While survey findings indicated that women were more likely to report a need for safety while traveling, qualitative insights from both men and women show that concerns about seizure-related risks while away from home are widely shared. Participants described anxiety about having seizures in unfamiliar settings, challenges with driving, and reliance on companions for travel. |
| | | Barriers to recreational activities | "the personal trainer tells us: "well, we're going to do these, it's this, this and this exercise in these repetitions". For me, it was like he had spoken to me in Japanese, because there's no way I can retain that...fortunately, now I was going with this gym buddy, I can tell him to please remember it."<br>Argentina, male, Participant 2<br><br>"I love going to concerts, they are great, but stroboscopes bother me...they bother me so much that I have to cover my eyes with my hands...I regret not going to concerts as often"<br>Croatia, Male, Participant 2<br><br>"I go to the gym, do CrossFit, run, and practice martial arts...but for about a month and a half now, I've been recovering from a knee injury caused by a seizure I had. It caused a medial collateral ligament strain. So, I'm going through recovery. I'm doing way less than usual"<br>Brazil, Male, Participant 1 | No corresponding quantitative item. | No integration required. |

| | | | | | |
|---|---|---|---|---|---|
| | | Constraints on household roles | "I also bought an induction oven, to feel safe. It’s for my safety because I love cooking and baking...if I feel like I'm having an off day, I just put things in a slow cooker and I let it cook all day. See I think ahead like that and I have a plug-in steamer the induction oven will turn off...I feel like some days I'm not as strong, so, my husband brings the laundry downstairs and stuff."<br>Canada, Rural (svd), Participant 2<br><br>"Before falling ill, I was the backbone of the household, taking care of all the chores such as grocery shopping, cooking, and cleaning...After I fell ill in 2017, my family adamantly refused to let me do any household chores""<br>China, Older (not svd), Participant 5" | **Divergence:** People who view epilepsy as a disability were significantly more likely to report that they need to feel well enough to help with household chores (29% vs. 21%; OR = 2.0, $p < 0.0001$). | Survey data showed that individuals who self-identified as having a disability were more likely to prioritise feeling well enough to help with household chores. However, this specific focus did not emerge strongly in qualitative interviews. While some participants described limitations with domestic tasks, these were often discussed only when prompted and framed within a broader pursuit of independence, with practical adjustments made to maintain daily routines. This divergence suggests that household chores may be just one expression of a wider goal not always articulated in the same terms during interviews. |
| | Parenthood, planning and uncertainty | Concerns around having children | “I thought if I passed my epilepsy on to my child, I would be very guilty, so I thought, I’d rather not have children.”<br>South Korea, Rural, Participant 5<br><br>I’m terrified that my child will also have epilepsy because I know the pain of epilepsy."<br>China, Rural, Participant 1<br><br>“it is their spouse who usually carries almost the entire burden of caring for the child or children… [wife] would have carried the burden of caring for the children. I would have done what I could. They probably would have given me more seizures"<br>Spain, Rural, Participant 5<br><br>"I often think about how I would handle taking care of children. I worry that the child might inherit epilepsy or develop another neurological condition. Taking care of children is already very challenging, and it becomes even harder if both the parent and the child have neurological issues."<br>Bosnia and Herzegovina, Male, Participant 5 | No corresponding quantitative item. | No integration required. |
| | | Gaps in knowledge and support on sexual and reproductive health | “If a pregnant woman has an epileptic seizure, can she lose her foetus or not? Does the medication have repercussions on fertility, especially on the foetus? Does the medication stop during pregnancy or not? Can I give birth or not? All these questions were like a movie running in my head and I could not find an answer to them and could not ask them in front of anyone.”<br>Tunisia, Male, Participant 2<br><br>“The sexuality of such people is a complete taboo. Some of us parents talk about it, others don’t. I don’t know what will happen when she hits puberty, whether she will be immature or want to have a boyfriend. It’s not discussed at all. So far, I’ve found out a little about contraceptives...”<br>Croatia, Caregiver (complex epilepsy), Participant 1<br><br>"We were negotiating [having children]. I risked myself...I said I'll just take one of the drugs at night and that's it. I said, I know I shouldn't say it, but a small seizure doesn't really affect me...That’s when I was with the doctor I didn’t like and didn’t trust them...I didn’t tell him [that PWE reduced medication during pregnany]....Doctors didn’t listen to what patients were saying."<br>Canada, Rural, Participant 2 | **Convergence:** While nearly half (46%) of survey respondents reported no needs related to sexual and reproductive health, 36% selected “simple information for people with epilepsy who want to become parents”, making it the most frequently selected need within this domain. | A need for clearer, more accessible reproductive information emerged in both datasets. Qualitative interviews described sexual and reproductive health as poorly addressed and difficult to navigate, often complicated by stigma, cultural taboos, and limited opportunities for open discussion with healthcare providers. Participants raised unanswered questions about pregnancy, medication safety, fertility, and contraceptives, with some expressing a lack of trust in medical advice or feeling unheard by clinicians. Although these concerns appeared less frequently in interviews than in the survey, they pointed to significant gaps in information and support for people with epilepsy and their families when making decisions about relationships, family planning, and pregnancy. |

| | | | | | |
|---|---|---|---|---|---|
| 2. **Living with risk, social exclusion, and misunderstanding** | Inadequate emergency response | Stigma and misunderstanding prevent safe responses | "I sincerely hope that more people around me can learn about epilepsy first aid knowledge. Just think about it, if one day you really encounter someone having a seizure, how much time would be wasted if they don't know what to do!"<br>China, Older (female), Participant 5<br><br>"my biggest concern is if one day I should get a blood clot or similar. Then people would think I had an epileptic seizure... Then I might die in that case, because it would be a heart attack and not an epileptic seizure...I'm nervous about that, it's my life at stake...I think this is a very overlooked issue"<br>Denmark, Male, Participant 2<br><br>"I think people should have basic knowledge on how to behave if someone in their environment experiences a seizure. They should know how to provide first aid to that person. I remember a few years ago, when a guy had a seizure in my town, and due to the inadequate help from those present, the guy unfortunately passed away."<br>Bosnia, Rural (female), Participant 4<br><br>"Once other people see that physical contact with someone who has a seizure isn't contagious then this gives other people more confidence to actually be supportive at the same time...A wider dissemination of knowledge and a dispelling of myths around epilepsy would go a long way. This would make people less cautious or apprehensive. Also, it would also make them less helpless and unable to help when they see one of us going through a seizure moment."<br>Uganda, Male, Participant 1 | **Complementary:** Female respondents were significantly more likely than male respondents to prioritise that 'basic seizure first-aid should be more widely known' (60% vs. 52%; OR = 1.4) as well as report a need to feel sure they will 'get the right help or support during an epilepsy emergency' (e.g., prolonged seizure, serious injuries suffered during a seizure in a public place) (57% vs. 49%; OR = 1.4, $p < 0.0001$) and other emergencies '(e.g., during conflicts, disasters, pandemic and other crises)' (49% vs. 42%; OR = 1.4, $p < 0.0001$). | Survey data showed that female respondents were significantly more likely than male respondents to prioritise the need for reliable support during epilepsy-related emergencies and broader crisis situations. In qualitative interviews, participants of all genders described feelings of urgency and fear when recounting such emergencies. Accounts varied: some emphasised misinformation or inadequate public awareness, while others focused on the emotional toll of past incidents or frustrations with systemic inaction. While a few female participants spoke in more personal terms about vulnerability and emotional burden, and some male participants highlighted public knowledge gaps or myths, these tendencies were not universal, and concerns about emergency response were broadly shared across genders. |
| | | Vulnerability to harm from others during seizures | "I heard of an incident where a man was killed because they thought he was a night dancer. He went onto the balcony of a woman's house in the night, facing the house, she kept asking him who he was and what he wanted but he just kept staring. She raised an alarm and people rushed to her rescue, he was beaten up and he died of his injuries. I have a feeling that he might have been like me and that worries me."<br>Uganda, Male, Participant 5<br><br>"In the beginning there were times when I would get a seizure and regain consciousness alone and sometimes with my property stolen."<br>Uganda, Older, Participant 3<br><br>"[Describing incident from approximately 30-40 years ago]. My school days were ruined by my epilepsy. I was bullied victims from the time I started school to the end. It was really serious bullying, my parents had windows smashed and I got beaten up and death threats. It was every day. It was because of my epilepsy. The others called me 'the crooked one'."<br>Denmark, Rural, Participant 1 | **Complementary:** Respondents from low-income countries were significantly more likely to report the need to 'feel safe from being physically hurt by others' (low: 73% vs. middle: 31% vs. high: 22%; OR = 11.6) | Survey results indicated that respondents from low-income countries were more likely to express a need to feel safe from physical harm by others. Qualitative accounts reinforced this concern, describing theft, assault, and fatal incidents arising from public misunderstanding and stigma. While such risks were most often described in low-income settings, participants from high-income countries also shared past experiences of violence alongside more frequent accounts of bullying and social exclusion. Together, the findings suggest that the perceived need for safety is shared, but the nature and prevalence of risk differ by context, with public misunderstanding emerging as a central driver across both datasets. |

Vulnerability and safety risks

| | | | | | |
|---|---|---|---|---|---|
| | | Vulnerability to harm from others during seizures | "I heard of an incident where a man was killed because they thought he was a night dancer. He went onto the balcony of a woman's house in the night, facing the house, she kept asking him who he was and what he wanted but he just kept staring. She raised an alarm and people rushed to her rescue, he was beaten up and he died of his injuries. I have a feeling that he might have been like me and that worries me."<br>Uganda, Rural (male), Participant 2<br><br>"[Describing incident from approximately 30-40 years ago]. My school days were ruined by my epilepsy. I was bullied victims from the time I started school to the end. It was really serious bullying, my parents had windows smashed and I got beaten up and death threats. It was every day. It was because of my epilepsy. The others called me 'the crooked one'."<br>Denmark, Rural (female), Participant 1<br><br>"I find it embarrassing or tense when some people behave with him. They either hit him or belittle him."<br>Tunisia, Caregiver (complex epilepsy, male PWE), Participant 4<br><br>" Another challenge when she gets older is that she can become a target for abusers."<br>Croatia, Caregiver (complex epilepsy, female PWE), Participant 1 | **Divergent:** Female respondents were more likely than male respondents to report a need to feel safe from being physically hurt by others because of my epilepsy (34% vs. 29%; OR = 1.6, $p < 0.0001$). | Survey data showed that women were more likely than men to report a need to feel safe from being physically hurt by others because of their epilepsy. This gendered difference was not clearly reflected in the qualitative data, where both male and female participants described similar experiences of harm, including violence, bullying, and other abuse. However, two caregivers (one caregiver of a female PWE and one caregiver of a non-binary partner with epilepsy), raised gender-specific risks related to domestic abuse, including physical, emotional, and coercive forms, as well as intimate partner violence. This suggests that some gendered vulnerabilities and identity-specific risks may exist but are less frequently voiced directly in first-person accounts. Overall, the qualitative findings did not mirror the broader gender gap observed in the survey, instead highlighting a more general concern about harm that cut across genders. |
| | | Mistreatment and exploitation in healthcare settings | "I think more measures should be taken to crack down on unlicensed hospitals as well as hospital scalpers. My wife and I once met a scalper who pretended to be a patient and tried to take my wife to other unlicensed hospitals".<br>China, Male, Participant 2<br><br>"At the beginning, my son's condition was very serious. The doctor was afraid that he would move around and affect the treatment, so he tied him to the bed."<br>China, Caregiver (non-complex epilepsy), Participant 3 | No corresponding quantitative item. | No integration required. |
| | | Humiliation, ridicule, and emotional harm | "My school days were ruined by my epilepsy. I was bullied victims from the time I started school to the end. It was really serious bullying... It was every day. It was because of my epilepsy. The others called me 'the crooked one'. It was actually so bad that I was shouted at in the corridors of the school. Everyone at the school shouted at me."<br>Denmark, Rural (female), Participant 1<br><br>"I have two close friends who have pointed out that I was much slower to respond during my seizures, making me seem less intelligent to them. Their sarcastic comments made me feel uncomfortable and self-conscious. It was as if they were mocking my condition, which I found hurtful."<br>China, Male, Participant 2<br><br>"Because of these misconceptions, I'm even more hesitant to talk about my condition. I'm particularly afraid that once others find out I have epilepsy, they will look at me with strange eyes, gossip behind my back, and distance themselves from me."<br>China, Older (female), Participant 5" | **Divergent:** Female respondents were more likely than male respondents to report a need to feel safe from being emotionally hurt by others because of my epilepsy (44% vs. 36%; OR = 1.5, $p < 0.0001$). | Survey data showed that women were more likely than men to report a need to feel safe from being emotionally hurt by others because of their epilepsy. This gendered difference was not clearly reflected in the qualitative data. In qualitative interviews, participants of all genders described experiences of stigma, exclusion, and misunderstanding, including bullying, mockery, gossip, and social distancing. Some accounts placed particular emphasis on the emotional impact of these encounters, describing feelings such as shame, fear, and withdrawal from social situations. While these perspectives were present across genders, the qualitative findings provide context for the higher reported need among women in the survey, highlighting the pervasive role of stigma and misunderstanding in shaping emotional safety concerns. |

| | | | | | |
|---|---|---|---|---|---|
| | | Sexual anxiety and loss of intimacy | "I get dizzy. So, I must be mindful of even this detail. Is that dizziness part of an aura or is it product of ejaculation [...] Is it an orgasm or is it a crisis?"<br>Argentina, Male, Participant 2<br><br>"Romantic and sexual life is affected. May be because of my irritation due to medicines I speak vulgar words so [wife] doesn't show interest."<br>India, Man, Participant 1<br><br>"I once had a partner where I had a seizure while we were lying in bed. She couldn't handle that at all and I never saw her again"<br>Denmark, Male, Participant 4 | No corresponding quantitative item. | No integration required. |
| | Intimacy and romantic relationships | Strains and shifts in existing relationships | "When we got married, I was supposed to be the trophy husband. She wanted to be the one that brought the money... But that was not what happened… She had to start counting on me… I had to take over leadership… that was not what she wanted."<br>US, Caregiver (non-complex epilepsy), Participant 5<br><br>"I had a boyfriend when I was about 21 or 22 and his mother was bothered by my epilepsy and we broke up."<br>Croatia, Rural, Participant 3<br><br>"After you find a partner who accepts you, agrees to marry you, down the line even when you start out positively in what might have been a good relationship, when things go wrong [epilepsy] is then used as a weapon against you and a reference or cause of failure. It's unfortunate but it's true."<br>Uganda, Male, Participant 1<br><br>"The first shock was with my wife who rejected my illness and insulted me a lot with her hurtful words which was the reason for the depression I reached...The period did not last long and we separated."<br>Tunisia, Older, Participant 5<br><br>"…we try to talk about our boundaries a lot…to make sure that one person or another, especially [partner], isn't feeling like I'm completely drowning [them]…Sometimes I try to take over…there's a fine, fine line there. I feel like it could easily go into domestic abuse if you have someone who is angry and doesn't realize they're channeling that anger in a wrong way…I could easily see that being a blurred line and being a problem."<br>US, Caregiver (non-complex epilepsy), Participant 5 | No corresponding quantitative item. | No integration required. |

| | | | | | |
|---|---|---|---|---|---|
| | | Romantic rejections and barriers to future partnerships | "If a child has it, then people don't tell about their condition as children have to get married later in life."<br>India, Rural, Participant 2<br><br>"Guys avoided me and said I was disabled."<br>Bosnia and Herzegovina, Rural, Participant 4<br><br>"there were a few times where you, where you mention it and they just all of a sudden don't want to talk to you anymore but it all depends."<br>US, Male, Participant 4<br><br>"throughout the years I put a damper on like romantic partners just because like for a long time I, you know, people avoided me because my [port-wine stain] birthmark, you know, and like back then like I was really down and out because I thought I was ugly and you know."<br>Canada, Male, Participant 5<br><br>"I had a romantic relationship before and he was watching me take the medicine. He didn't ask me about it, but I felt from his actions that he had looked for it and who was the person required to use it after seeing the medicine box. After a long period of time with each other, I noticed that his actions had changed and we started to quarrel a lot for the most trivial reasons that he used to fabricate for the illness that I was living with. For this reason, I cut off the relationship with him... this is what makes me keep my illness a secret from everyone."<br>Tunisia, PWE (additional, female), Participant 2 | **Complementary:** Respondents from low-income countries were significantly more likely than those from middle- and high-income countries to report that they need advice on how to tell key people in their life about their epilepsy (e.g., about their diagnosis, how it affects their life) (68% vs. 25% vs. 27%; OR = 7.9, $p < 0.0001$). | Survey data showed that respondents from low-income countries were significantly more likely than those from middle- and high-income countries to report needing advice on how to tell others about their epilepsy. This pattern was not clearly reflected in the qualitative data overall. Conversations about disclosure in general (e.g. in schools, workplaces, or friendships) were more frequently raised by participants from middle- and high-income countries, suggesting divergence from the quantitative finding. However, in the context of romantic relationships, qualitative data complements the survey result. Participants from low-income countries described secrecy, fear of rejection, and discomfort when disclosing their epilepsy to potential partners. In contrast, participants from middle- and high-income countries more often reflected on having disclosed, and being rejected as a result. |
| | | Loss of autonomy and daily independence | "Before I ate the world [full of confidence], and today I am more dependent."<br>Argentina, Older, Participant 1<br><br>"...for me, that was the biggest down. Down issue was the fact I lost my driver driver's license last Monday. Lost my independence, my mobility and again after 60 years of driving at will anywhere kind of saying it really hit me like a hammer blow."<br>Canada, Older, Participant 4<br><br>"Before falling ill, I was the backbone of the household, taking care of all the chores such as grocery shopping, cooking, and cleaning...After I fell ill in 2017, my family adamantly refused to let me do any household chores"<br>China, Older, Participant 5 | No corresponding quantitative item. | No integration required. |

Barriers to personal growth and development

| | | | | | |
|---|---|---|---|---|---|
| | | Missed opportunities for learning and career development | “Another incident was of a child who was sitting her primary leaving exams. When the child didn’t perform to expectation the parents got really ashamed of her performance. They halted her education because of that. She thereafter requested that she be allowed to take up tailoring and was denied saying she might make a repeat of her earlier disappointment."<br>Uganda, Caregiver (complex epilepsy), Participant 2<br><br>“epilepsy excludes you from many public job competitions. My dream profession since I was a teenager was to become a federal police officer. But in every competition for police positions, when you look at the list of medical conditions that disqualify candidates, epilepsy is almost always one of the first on the list under nervous system disorders"".<br>Brazil, Male, Participant 1<br><br>“The community organized a workshop, and I was invited as one of the chairs. Then one of the community members informed the organizer that I am someone who lives with epilepsy and I cannot take this responsibility. And this happened on two occasions, and I’ve never been selected”.<br>The Gambia, Older, Participant 5<br><br>"Even by my former boss. I can see that he did not count the work I did on an equal footing with others. I wasn't allowed to use the skills that I actually had. I was quickly broken down and told that you can't.”<br>Denmark, Rural, Participant 1 | No corresponding quantitative item. | No integration required. |
| | | Erosion of social participation | "They dare not make friends or fall in love like normal people, always worrying that others will despise them once they know about their condition. They end up isolating themselves day by day, becoming more and more introverted and less confident.”<br>China, Older, Participant 5<br><br>"the social perception of this disease is not that good. And I just keep saying that I don't drink because I don't want people to ask me questions about my disease anymore. I also think that my relationships are narrowing a little bit. Because I have to hang out with people who don't drink."<br>South Korea, Rural, Participant 4<br><br>“Some friends directly blocked him on [social media] and deleted his phone number as soon as they heard he had this disease. It's obvious that they are distancing themselves from him. They think he has a bad disease and are afraid of being affected, so they don't want to have anything to do with him anymore.”<br>China, Caregiver (non-complex epilepsy), Participant 3 "<br><br>"For example, 'Oh, he has epilepsy, so let’s not invite him to an event with flashing lights,' without even knowing if that’s a trigger. Or sometimes someone makes a really tasteless joke.”<br>Bosnia and Herzegovinia, Male, Participant 1 | No corresponding quantitative item. | No integration required. |

| | | | | | |
|---|---|---|---|---|---|
| | | Gaps in inclusion and support systems | “Our association, which has around 60 members, tries to break the stigma and provide support to those affected. Others often don’t join because of the fear of stigma.”<br>Bosnia and Herzegovina, Caregiver (non-complex epilepsy), Participant 1<br><br>“some years ago I came across of a support group... but that did not last long and their procedures are so tedious we could not proceed with that. I also came to find out that that support group is helping themselves more than the patient.”<br>The Gambia, Caregiver (complex-epilepsy), Participant 3<br><br>“I have observed that our government has not yet implemented programs that fully protect people living with epilepsy or prioritise their rights within the community”.<br>Uganda, Male, Participant 1 | **Complementary:** 40% of survey respondents identified a need for programs and support to improve confidence and achieve personal goals. | Survey findings showed that 40% of respondents identified a need for programs and support to improve confidence and achieve personal goals. Qualitative accounts did not explicitly link these gaps to confidence-building, but described barriers to inclusion such as limited awareness of available services, short-lived or inaccessible initiatives, and government inaction in protecting the rights of people with epilepsy. Stigma also discouraged participation, with some avoiding support groups for fear of being judged. Together, these findings suggest that while the survey highlights aspirations for personal development, interview narratives frame the issue more in terms of systemic, informational, and social barriers to equitable participation. |
| **3. Challenges in navigating inaccessible systems** | Barriers to accessible healthcare | Barriers to accessing care and information | "Medication outages, when I go to the hospital and there is no medication, I run the risk of getting a seizure, to or from the hospital. There are extremes where I go even two months without the medication being available."<br>Uganda, Male, Participant 1<br><br>"Sometimes when I go there it’s not available. So, I have to return or send someone to bring it."<br>Uganda, Older, Participant 3<br><br>"If you go to public health, for example, in speech therapy, it is months and I would say that even years that you can wait for an appointment."<br>Argentina, Caregiver (non-complex epilepsy), Participant 5<br><br>"I can even say that it’s almost impossible to get feedback on whether my lifestyle is good, or if there’s anything I could change. I need someone who can provide advice regarding epilepsy and my lifestyle. I need someone who is immediately available, not someone I have to wait six months to see."<br>Bosnia & Herzegovina, Older, Participant 3" | **Complementary:** Respondents from low income countries were significantly more likely than those from middle- and high-income countries to report that there is a need for faster access to the right treatment (e.g., medicine, procedures) (low: 85% vs. middle: 56% vs. high: 48%; OR = 5.3, p < 0.0001). Similarly, ‘ongoing access to epilepsy treatment’ was selected more often in low-income settings (low: 83% vs. middle: 49% vs. high: 39%; OR = 6.2). Furthermore, the most commonly selected need across all domains was ‘easier access to healthcare services’ (57%).‘’Faster access to the right treatment’ ranked fourth across all domains and subgroups (55%). | While survey data showed that respondents in low-income countries were significantly more likely to report a need for faster access to the right treatment, interviews revealed that this need is often rooted in the basic availability of treatment itself. Participants described repeated medication shortages, long travel distances, and being instructed to purchase medicine privately, often at unaffordable prices. In contrast, participants from higher income countries more frequently cited long waiting times (e.g. for diagnostic tests) or the unavailability of timely feedback from specialists. These differences suggest that in lower income settings, access challenges are more fundamental, whereas in higher income contexts, delays may reflect system inefficiencies or gaps in care coordination. |
| | | Barriers to accessing care and information | "When he was one year old that’s when I witnessed him having seizures. In that situation, I tried to seek help from people so that he’ll be rushed to the health centre at [town]. The healthcare professional only prescribed medication like carbamazepine, but I was not informed that it was epilepsy."<br>The Gambia, Caregiver (complex epilepsy), Participant 3<br><br>"Previously we were given medication but without explicit guidelines on never changing or sharing medication, timeliness of dosage."<br>Uganda, Male, Participant 1<br><br>“I’ve noticed that doctors… are often not very well-informed about epilepsy. Most of my knowledge comes from my personal experience and research.”<br>Bosnia & Herzegovina, Male, Participant 1 | **Complementary:** Respondents from low-income countries were significantly more likely to report needing access to high-quality information about epilepsy (e.g., seizure control, triggers, treatment options, side effects) compared to those from middle- and high-income countries (77% vs. 46% vs. 35%; OR = 7.0, p < 0.0001). | The high demand for better information among respondents from low-income countries is reinforced by interview accounts that highlight serious gaps in accessing even the most 'basic' medical information such as receiving a diagnosis without explanation or being prescribed medication without clear instructions. In contrast, interviews from higher-income countries more often revealed concerns around more advanced / nuanced information needs, such as support for emotional wellbeing or being signposted to patient advocacy groups. These findings suggest that while epilepsy information gaps are a global concern for PWE, their nature and depth vary by context ranging from fundamental communication failures in low-resource settings to unmet needs around holistic care in higher-resourced ones. |

| | | | | | |
|---|---|---|---|---|---|
| | | Barriers to accessing care and information | "The silo here is that, first up, he has a seizure, you bring him to the family doctor, the family doctor refers to the only paediatric neurologist in the city, which is a seven week wait, and you go and you see her. And then she says, oh, this is a complicated case. You need to go to [location] and then it's a six month wait. So you wait again. Just get passed along. There's limited communication between everyone and then in the meantime, for us anyway, we're a border city to [neighbouring country]. So we actually took [our son] twice now over to [neighbouring country] for care to skip the six month wait times down here for testing."<br>Canada, Caregiver (complex epilepsy), Participant 1<br><br>""Unfortunately, not all medications available in the EU are available in the Republic of Croatia. Some medications are on the list, but you can't get them without medical committees. Automatic medications that go to committees are not prescribed because doctors don't want complications. That's completely bad.""<br>Croatia, Caregiver (complex epilepsy), Participant 1<br><br>""Medication outages, when I go to the hospital and there is no medication, I run the risk of getting a seizure, to or from the hospital. There are extremes where I go even two months without the medication being available.""<br>Uganda, Male, Participant 1" | **Complementary:** Respondents who identified as belonging to a minority group were significantly more likely than non-minority respondents to report needin ongoing access to treatment (55% vs. 47%; OR = 6.2, $p < 0.0001$), and tailored healthcare (46% vs. 37%; OR = 7.3, $p < 0.0001$). | Across Domain 3 (Healthcare and wellbeing), survey respondents from minority backgrounds were more likely to report unmet needs across nearly all items. The most significant findings indicate consistent disparities in access to rare or complex epilepsy services, ongoing access to treatment,and receiving tailored healthcare. While interview participants did not explicitly self-identify as belonging to a minority group, we explored the experiences of those who described exclusion or rejection directly linked to their epilepsy. What unites these accounts is not ethnicity, migration status, or cultural identity, but the social marginalisation that arises specifically because of epilepsy. As such, the healthcare barriers described long wait times, lack of tailored care, medication shortages, and systemic inefficiencies are best understood as consequences of epilepsy-related exclusion. While this does not offer a complete picture of all minority group experiences (e.g., it does not capture the impact of language barriers or cultural dissonance that may affect some ethnic minority or immigrant groups), it does offer insight into how epilepsy itself can lead to 'minority-like' status in the healthcare system. |
| | | Barriers to accessing care and information *and* Economic impact of epilepsy | "The government should focus on providing better access to healthcare services for people with epilepsy, including specialized treatment and medications"<br>Bosnia & Herzegovina, Rural, Participant 4<br><br>The truth that breaks my soul when I hear that among their families, they collect the money to be able to buy anti-epileptic medication. And that doesn't happen in Argentina, except when we are prescribed an epilepsy medication that is very expensive that the public service does not cover it.<br>Argentina, Male (urban/suburban), Participant 2<br><br>"I mean, we pay $12,000 a year just to have insurance for both of us. It's $12,000. That doesn't include all the co-pays... $12,000 just in paying for insurance every year so that she can make sure she gets the meds. We still have to pay for meds a little bit every month. We still, anytime we go to an appointment, we're paying 50 bucks. It's a lot of money and and that is awful for someone with epilepsy who has to go in a lot."<br>US, caregiver (non-complex epilepsy, urban/suburban), Participant 5<br><br>""...I don't think so the government is doing much in this aspect... The number one priority is medication. This is very common in the healthcare setting not only anti-epileptic drugs"<br>The Gambia, Caregiver (complex epilepsy, rural), Participant 3" | **Divergence:** Respondents from rural areas were significantly more likely than those from suburban or urban areas to report that their country needs to protect the rights of people with epilepsy (59% vs. 54% vs. 50%; OR = 1.5, $p < 0.0001$). | Respondents from rural areas were significantly more likely than their suburban or urban counterparts to report that their country needs to better protect the rights of people with epilepsy). However, this distinction was less apparent in the qualitative data. While rural participants did express concerns particularly around access to medication and financial burden, frustrations with government protection and support were voiced across all settings. Participants described a lack of government action to uphold the rights of people with epilepsy, poor access to specialised healthcare and essential medications, and significant financial strain caused by the need to pay for treatment out of pocket. These accounts suggest that while the survey data shows a clear statistical difference by location, the lived experiences captured in interviews reflect a more widespread sense of dissatisfaction with existing rights and government intervention. |

| | | | | | |
|---|---|---|---|---|---|
| | | Barriers to accessing care and information *and* Economic impact of epilepsy | [see also quotes above]<br><br>"We live in a rural town, and some epilepsy medications are not available at the local hospital...the medication I take...is an imported drug, so I have to purchase it from online pharmacies...delivery time is not guaranteed... possibility of counterfeit drugs online...I hope that the local government can provide more policy support for epilepsy patients... Additionally, I wish there could be more reimbursement coverage for epilepsy-related expenses. The imported medication I take has to be paid for entirely out of pocket...This financial burden causes me a lot of psychological stress."<br>China, Older (rural), Participant 5<br><br>"It is a priority for the government to provide public centres for patients to take care of this category in addition to the university hospital or local hospitals, and for a subscription fee that suits the individual's financial capacity."<br>Tunisia, PWE (additonal, female), Participant 2 | **Complementary:** Respondents from low-income countries were more likely to agree that their country needs to recognize and defend the human rights of people with epilepsy (e.g., through laws, nationwide campaigns, etc.) compared to those from middle- and high-income countries (78% vs. 59% vs. 42%; OR = 5.4, $p < 0.0001$). A similar pattern was observed in response to the statement "my country needs to protect the rights of people with epilepsy", with higher agreement among respondents from low-income countries (77% vs. 60% vs. 39%; OR = 7.3, $p < 0.0001$). | Respondents from low-income countries placed particular emphasis on the need to protect the rights of people with epilepsy, a view reinforced by qualitative accounts highlighting the effects of legal and structural neglect, such as lack of legislation, limited access to education, and economic exclusion. While this call for systemic reform was strongest in low-income settings, the belief that governments should reduce the financial burden of epilepsy and improve access to care was shared across all countries, and was especially acute in middle-income contexts. Civic advocacy was more visible in high-income settings like the USA and South Korea, reflecting context-specific strategies. |
| | | Healthcare services under-resourced | "In [my country], there are no specialists for epilepsy, and neurologists who are well-versed in epilepsy are very rare."<br>Bosnia & Herzegovina, Male, Participant 1<br><br>"To be honest, within our community we do not actually have diagnostics or machines to check for Epilepsy, we are dependent on the doctors knowledge and experience alone"<br>Uganda, Caregiver (non-complex epilepsy), Participant 1<br><br>"I would definitely say that it's a lot harder for other people. Because they're such a long waiting list and things like that, there's not enough epileptologists out there. To help these people so access to like. Like healthcare professionals is definitely harder. And. Insurance companies finding and gain insurance, so that your medicine can be covered is definitely harder. Like, that's something that has always been crazy. Like when I look for job, when I was looking for a job, I also had to look to see if they have insurance because as soon as soon as you turn 26 here, you're kicked off your parents insurance."<br>US, Male, Participant 4 | No corresponding quantitative item. | No integration required. |

| | | | | | |
|---|---|---|---|---|---|
| | | HCP knowledge and support | "...no other neurologist could understand... None of them... I went to a neurologist who asked, 'What is it? Tell me, what is it?' He wanted me to tell him what epilepsy is."<br>Brazil, Rural, Participant 3<br><br>"...epilepsy is considered [by healthcare professionals] to be only an irregularity in the electrical functioning of the brain...only a biological one and nothing else. And it despises the fact that we are holistic integral beings, and that I also have [seizure] when I fight with.... when I got divorced for example. Or, when they didn't pay me my salary...made me worry and I have a [seizure]."<br>Argentina, Male, Participant 1<br><br>"I've noticed that doctors… are often not very well-informed about epilepsy. Most of my knowledge comes from my personal experience and research."<br>Bosnia & Herzegovina, Male, Participant 1"<br><br>"I also believe that emotional changes after a seizure need more attention...I think this kind of information needs to be discussed more openly and made clearer to patients. I believe that doctors and nurses are taught how to manage seizures during their training, but these emotional and psychological aspects seem to be overlooked."<br>Brazil, Male, Participant 1<br><br>"The doctor said a lot of things I wasn't allowed to do. But I didn't want to put my life on hold either."<br>Denmark, Male, Participant 4 | **Complementary:** Respondents from low-income countries were more likely than those from middle- and high-income countries to report that doctors and nurses need to know more about people's wider needs linked to epilepsy (e.g., emotions and mental health needs of people with epilepsy at different stages of life) (77% vs. 46% vs. 49%; OR = 4.4, $p < 0.0001$). The need for 'doctors and nurses to know more about people's wider needs' was the third most-selected need within Domain 3. | While survey results showed that respondents from low-income countries were more likely to report that doctors and nurses need to better understand people's wider needs linked to epilepsy, interviews revealed what constitutes 'wider needs' may differ by context. Participants from low-income countries tended to focus on treatment-specific gaps (e.g. availability of medication), follow-up care, or basic clinical communication, rather than emotional or mental health needs. In contrast, participants from middle- and high-income countries more frequently mentioned the psychological and emotional dimensions of living with epilepsy. This difference may reflect variations in healthcare infrastructure, where in low-income settings the absence of basic medical support overshadows more nuanced psychosocial concerns. Together, these findings suggest that widening the clinical lens to include emotional wellbeing should be an approach adopted in all settings. |

Gaps in quality of care and support

| | | | | | |
|---|---|---|---|---|---|
| | | Quality of medical practice | “I wish the medical staff could have a moment to exchange detailed information about the patient's illness. I can't even ask questions properly right now. Rather, when I try to speak as a patient carer, he sighs deeply and stops me from asking questions.”<br>South Korea, Caregiver (complex epilepsy), Participant 1<br><br>"When he was one year old that’s when I witnessed him having seizures. In that situation, I tried to seek help from people so that he’ll be rushed to the health centre at [town]. The healthcare professional only prescribed medication like carbamazepine, but I was not informed that it was epilepsy."<br>The Gambia, Caregiver (complex epilepsy), Participant 3<br><br>"Previously we were given medication but without explicit guidelines on never changing or sharing medication, timeliness of dosage."<br>Uganda, Male, Participant 1<br><br>"[Speaking about other PWE and knowing the cause of epilepsy] ...and they don't know that...I tell them that they ask their doctors. Nothing, [doctors] don't inform, they don't inform them why. If I have stayed with the previous clinic, it was also the same. That's why I didn't want to stay. I wanted to continue understanding and looking for the why, what causes it for me."<br>Argentina, Rural, Participant 3<br><br>...they should have more information to tell us about treatment of seizures to tell us and also they should know and tell us more about the management of seizures. I feel that someone should tell me how to take medicine, for how long should I continue it and how will I become alright but no one says so. All of them just say, 'You have to take medicine, you have to take it compulsorily'."<br>India, Rural, Pariticpant 2 | **Complementary:** Respondents from low-income countries were significantly more likely than those from middle- and high-income countries to report that they need advice on how to have useful conversations with doctors and nurses (69% vs. 27% vs. 25%; OR = 7.9, p < 0.0001). | The survey finding that people in low-income countries are significantly more likely to want advice on how to have useful conversations with doctors and nurses (69% vs. 27% vs. 25%; OR = 7.9, p < 0.0001), is supported by qualitative insights. While there is no direct reference to the dialogue between PWE / CGs and HCPs in the interviews with participants from low-income countries, they do highlight how a lack of 'access' to meaningful dialogue with HCPs can result in relying on one's own expereinces to learn and understand their epilepsy. In these settings, access to HCPs itself was often limited (time and resource for long journeys frequently mentioned) leaving little opportunity to focus on the 'content' of the conversations. Across income groups, especially in rural areas, a similar challenge appears - not only about 'what' is said, but whether there is an 'accessible HCP (often specialist)'. In other income settings, missed opportunities 'within' the consultations were more commonly described. |
| | | Treatment challenges | "Then I had a second operation here in [city A] and that second operation was a failure, that is, it has a bad result."<br>Argentina, Older, Participant 1<br><br>"So he put me on a certain type of tablet, which changed my personality according to my wife, was didn't, didn't go well with that. So that was tegretol. And then I got changed over to Keppra and it's been heaps better since then."<br>Australia, Male, Participant 3<br><br>"Over the past few months, we have gradually reduced some of the epilepsy medications she was taking. We have streamlined the number of medications, so she is now on fewer drugs, which means fewer side effects."<br>Brazil, CGC, Participant 5 | No corresponding quantitative item. | No integration required. |

| | | | | | |
|---|---|---|---|---|---|
| | Barriers to mental health support | Barriers to accessing support and coping strategies | "I've taken 50 years to go to the psychiatrist. I needed a psychiatrist, I don't know how long ago, I've had a really bad time you know. This result, what I told you before, this is just a pill and 10 minutes of consultation. But if they can't coordinate, if they can't talk to each other then what do we do? Each one does his own thing. That is, it has to be "all together" like what the Germans do. It has to be a whole. You cannot just diagnose epilepsy and treat epilepsy. What about the rest? The frustration... The rest how do we treat it?"<br>Spain, Male, Participant 4<br><br>"I haven't noticed any specific [mental health] support services for people with epilepsy...Maybe they exist, but they are not well promoted or accessible."<br>Bosnia & Herzegovina, Male, Particpant 5<br><br>"[wiping away tears] No, no, they don't coordinate, they coordinate... I talk with my neurologist and spoke in turn with the psychologist... what would be beneficial is that you also have a psychiatrist. That balance the epileptic medication with the psychiatric medication that is pulling you back with something that lifts you up."<br>Argentina, Rural (female), Participant 3 | **Complementary:** Female respondents were significantly more likely than male respondents to report that there needs to be 'better planning between mental health care and epilepsy care' (51% vs. 42%; OR = 1.4, p < 0.0001). | Whilst quantitative data show that female respondents were significantly more likely than males to report a need for better planning between mental health care and epilepsy care (42% vs. 51%; OR = 0.7, p < 0.0001), this emphasis was not reflected in the qualitative interviews. |
| | | Barriers to accessing support and coping strategies | ""I don't currently have therapy for my anxiety, but I feel that I need it…. Maybe someone needs a psychologist…but unfortunaely such services are unavailable.""<br>Bosina & Herzegovina, Rural, Female, Participant 4<br><br>"...psychological support is more important than someone helping me if I had a seizure. Here on the island, I don't have anyone to talk to about epilepsy, but I don't want to bother them, and I don't have time to speak to a psychologist. I've never tried."<br>Croatia, Rural, Female, Participant 3<br><br>""...here, we are underserved. We don't have any psychiatric or psychological support, or any social assistance for that matter, that provides intensive or comprehensive care. There's no support in that sense. What happens is... I have medical follow-ups in the state capital. It's quite far from the city where I live, It's a bit more than 200 kilometres away. So… it's challenging, both in terms of transportation and cost...""<br>Brazil, Rural (Male), Participant 3<br><br>""...I was not referred for a psychiatrist by my neurologist despite the fact that I had shown symptoms of depression but it was my family who supported me and who demanded that I see a psychiatrist but late in life, I had already ruined my married life.""<br>Tunisia, Older (Male), Participant 5" | **Complementary:** Female respondents were significantly more likely than male respondents to report that they need access to 'mental health care that is specific for people with epilepsy' (47% vs. 40%; OR = 1.4, p < 0.0001), and 'affordable mental health services specific to people with epilepsy' (48% vs. 41%; OR = 1.4, p < 0.0001). | Quantitative data indicate that female respondents are significantly more likely than males to report a need for mental health care tailored to people with epilepsy and to report a need for more affordable mental health services. However, the qualitative data did not reveal clear gender differences in experiences related to access or affordability. Instead, both female and male participants described similar challenges, including a lack of referral pathways, absence of locally available services, and the emotional burden of managing epilepsy without adequate psychological support. Participants across genders shared feelings of being overlooked by the health system and described how support was often sought only after distress had escalated or when prompted by family members. These insights point to a broader, systemic issue: the need for more accessible, responsive, and epilepsy-specific mental health care for all, regardless of gender. |

| | | | | | |
|---|---|---|---|---|---|
| | | Barriers to accessing support and coping strategies | "...in my case...I was the one coordinating the psychiatrist with the psychologist...I talked with my neurologist and spoke in turn with the psychologist...in fact, with the medication, what would be beneficial is that you also have a psychiatrist. That balance - the epileptic medication with the psychiatric medication that is pulling you back with something that lifts you up."<br>Argentina, Rural, Participant 3<br><br>"I haven't noticed any specific support services for people with epilepsy in [participant's country]. Maybe they exist, but they are not well promoted or accessible....group therapy and meetings would be an excellent way to provide support, but such options currently do not exist."<br>Bosnia & Herzegovina, Male, Participant 5<br><br>"I was not referred for a psychiatrist by my neurologist despite the fact that I had shown symptoms of depression but it was my family who supported me and who demanded that I see a psychiatrist but late in life, I had already ruined my married life."<br>Tunisia, Older, Participant 5 | In Domain 8 (Mental health), respondents identifying as minorities were more likely to report needs across all 8 items (OR = 1.3 to 1.6, $p < 0.0001$). For example, more minorities reported needing access to epilepsy-specific mental health care (51% vs. 42%; OR = 1.5, $p<0.0001$), better coordination between mental health and epilepsy care (56% vs. 45%; OR = 1.6, $p<0.0001$), and affordable mental health services (52% vs. 43%; OR = 1.4, $p<0.0001$). | The survey found that respondents identifying as minorities were more likely to report mental health needs across all items in this domain. For example, more minorities reported needing access to epilepsy-specific mental health care, better coordination between mental health and epilepsy care, and affordable mental health services. While interview participants did not explicitly self-identify as belonging to a minority group, we explored the experiences of those who described social exclusion or rejection linked to their epilepsy. What unites these accounts is not ethnicity or cultural identity, but the social marginalisation that arises because of epilepsy. These interviews shed light on the nature of the mental health needs (included in the survey): they described a lack of coordination between mental and epilepsy care, limited or poorly promoted psychological services, absence of referrals, and reliance on families to push for support. These accounts suggest mental health needs are often unmet due to fragmented systems and low visibility of services. |
| | | Stress and environmental triggers | "I believe that regular support from my family, combined with learning how to manage stress and frustration, would help improve my mental health."<br>Bosnia, Rural, Participant 4<br><br>"Reducing stress would help me the most. For example, when I don't have many school obligations, I feel much better. Homework, studying, and other tasks create significant stress, and stress, as psychologists often say, acts as a "silent killer" that negatively impacts mental and overall health. There should be ways for people with epilepsy to better manage stress, as it greatly affects their condition and quality of life."<br>Bosnia and Herzegovina, Male (urban/suburban), Participant 5<br><br>"One of my main triggers is stress, so that's why I get into the volunteering. I just love the better mental health aspects of it. Watching that stress, I do yoga right before I go to bed. Just a a simple one. That's more flexibility than actually working out and it just relaxes the muscles help you release stress right before you go to bed, and that helps me sleep a lot better."<br>US, Rural, Participant 3<br><br>"[To take care of their epilepsy] I try to get enough sleep to reduce stress...It all comes down to stress. I like to meditate"<br>Croatia, Male (urban/suburban), Participant 2 | **Divergence:** Rural respondents were more likely to report a need to decrease stress levels or learn to better manage it (rural: 57% vs. suburban: 50% vs. urban: 47%; OR = 1.5). | Quantitative data highlighted a stronger reported need among rural respondents to reduce or better manage stress, but this location-based difference was not clearly reflected in the qualitative findings. Participants across rural, urban, and suburban settings consistently identified stress as a major challenge, linking it to seizure control, emotional wellbeing, and daily functioning. They described various coping strategies, such as family support, volunteering, and relaxation techniques, but did not frame their experiences in relation to geographic context. |

| | | | | | |
|---|---|---|---|---|---|
| | | Equitable access to education | Another important issue is the government did not have any facility or any educational program for these kids who were affected by epilepsy... government should have set up a place like a school, where they can enroll these kind of kids living with epilepsy to be educated because it's very important and this is a concern to me as a mother."<br>The Gambia, Caregiver (complex epilepsy), Participant 3<br><br>"most importantly, other than medication is access to education. The government should help those living with epilepsy to have equal opportunity to education as any other child this is very important and is my concern. I will be very happy if my children can go to school online.<br>The Gambia, Caregiver (non-complex epilepsy), Participant 4 | **Complementary:** Respondents from rural areas were significantly more likely than those from suburban or urban areas to report that their country needs to protect the rights of people with epilepsy (59% vs. 54% vs. 50%; OR = 1.5, $p < 0.0001$).<br><br>Also, respondents from low-income countries were more likely to agree that their country needs to recognize and defend the human rights of people with epilepsy (e.g., through laws, nationwide campaigns, etc.) compared to those from middle- and high-income countries (78% vs. 59% vs. 42%; OR = 5.4, $p < 0.0001$). A similar pattern was observed in response to the statement "my country needs to protect the rights of people with epilepsy", with higher agreement among respondents from low-income countries (77% vs. 60% vs. 39%; OR = 7.3, $p < 0.0001$) | Survey data showed that respondents from rural areasand low-income countries were significantly more likely than those from suburban/ urban areas and middle/high-income countries to say their country needs to protect the rights of people with epilepsy. While qualitative accounts reflected a broad dissatisfaction with existing rights and government action across all locations, rural Gambian participants specifically emphasised the right to equitable access to education for children with epilepsy. These examples from rural participants in a low-income country highlight education as a specific rights-related gap, partially complementing the broader quantitative finding but focusing on one concrete aspect rather than the full scope of rights referenced in the survey. |
| | Barriers to equitable education | Lack of institutional support | "There is a lot of discrimination in the schools, because they do not know how to act. There are many cases, like the one that happened to a peer.., where they [the school] asked her to change schools because she didn't have the skills for that school and the school wasn't prepared."<br>Argentina, Rural, Participant 3<br><br>"There is no general protocol for emergencies where some children in an educational institution would be given medicine to stop a seizure."<br>Croatia, Caregiver (complex epilepsy), Participant 1<br><br>"I had to drop out of college when I had a seizure in the computer lab. This was like 6 months before they passed the Americans with Disabilities Act. And I had to drop out of college because the professor would not take my final that I was trying to turn in at that time in the computer lab, I had a seizure, was taken by ambulance to an emergency room. And he just refused to accept it. He was like you went past the deadline, so I was like I was in the hospital. I was in this computer lab and stuff and he was like Nope, not gonna take it. And the final was worth half my grade, so."<br>US, Rural, Participant 3<br><br>"during an education class in [university[, I had a seizure. They called an ambulance, and everything. But later, the professor suggested to the program coordination that it might be better to remove me from the class because I was 'too stressed'."<br>Brazil, Male, Participant 1 | No corresponding quantitative item. | No integration required. |

| | | | | | |
|---|---|---|---|---|---|
| | | Limited awareness and stigma in education settings | Once I had a seizure during an anatomy exam…there wasn't much support from the institution…some professors thought I was pretending because I hadn't studied for the exam."<br>Brazil, Male, Participant 1<br><br>"I have been judged and misunderstood by many. I can give an example of a lecturer of mine at the university....one day, in his class while lecture was on I told him that I wanted to lay my head on the desk I am not feeling well, I am someone living with epilepsy, he did not take me serious and he did not accept me to lay my head. Then he said to me the way you are performing in my class you don't seem to be an epilepsy patient. He did not accept the fact that I live with epilepsy despite the explanation."<br>The Gambia, Male, Participant 1 | Survey respondents from lower-income countries were more likely to report the need for teachers to 'learn about epilepsy so that they don't have unfair beliefs' (low: 78% vs. middle: 59% vs. high: 60%; OR = 2.7). | Survey data showed that respondents from low-income countries were more likely than those from middle- or high-income countries to call for teacher education to counter unfair beliefs about epilepsy. Qualitative accounts, however, revealed that such stigma, disbelief, and lack of reasonable adjustments from educators occurred across all settings, suggesting the problem is more widespread rather than confined to low-income contexts. |
| | | Limited awareness and stigma in education settings | Once I had a seizure during an anatomy exam…there wasn't much support from the institution…some professors thought I was pretending because I hadn't studied for the exam."<br>Brazil, Male, Participant 1<br><br>"Initially he wasn't too bothered by the revelation of having the condition but it later hurt a bit when he was ostracised at school...there still remains the occasional incident but he's better now."<br>Uganda, Caregiver (non-complex epilepsy), Participant 1<br><br>"The teacher once said to me, 'What will I do if he has a seizure?' I explained what to do and that he hadn't had seizures for years... but during one incident, they called an ambulance... he was fine... but the teacher had a heart attack. Fortunately, the emergency services that had come for my child helped the teacher."<br>Bosnia & Herzegovina, Caregiver (complex epilepsy), Participant 2<br><br>"...at school, I don't have close friends. My relationships with schoolmates are mostly limited to standard school matters, like talking about assignments, group projects, and similar activities."<br>Bosnia & Herzegovina, Male, Participant 5 | **Complementary:** Respondents from low income countries were significantly more likely than those from middle- and high-income countries to report that they would like to feel more included in the learning setting (e.g., teachers letting all students know that differences are normal) (77% vs. 44% vs. 40%; OR = 7.3, $p < 0.0001$) and free from stigma and negative attitudes from fellow students (80% vs. 44% vs. 46%; OR = 5.0, $p < 0.0001$). | Survey data indicated that respondents from low-income countries were more likely than those from middle- and high-income countries to report needing greater inclusion in the learning environment and freedom from stigma. Qualitative findings echoed these needs but revealed they were common across all settings, not confined to low-income contexts. Participants described being denied opportunities, lacking necessary adjustments, or facing stigma and academic discrimination from both peers and educators, sometimes rooted in misconceptions about epilepsy or fear of seizures. Caregivers and students alike stressed the importance of better-informed, proactive educators to foster genuinely inclusive learning environments. |
| | | Need for adjustments | "He can't take long exams or sit down to study all afternoon. He sits for an hour or so and has to take a break because he loses concentration".<br>Spain, Caregiver (non-complex epilepsy), Participant 3<br><br>"He has an individualized approach at school due to reading problems, he doesn't understand what he reads, he can't concentrate for long"<br>Croatia, Caregiver (non-complex epilepsy), Participant 5<br><br>"My daughter struggles with attention, concentration, and is often less physically active. She reads and completes her tasks slowly. Because of these challenges, she has a teaching assistant."<br>Bosnia & Herzegovina, Caregiver (non-complex epilepsy),, Participant 1 | No corresponding quantitative item. | No integration required. |

| | | | | | |
|---|---|---|---|---|---|
| | | Barriers to employment for PWEs | "Historically, some believed epilepsy was of demonic origin. Because of this, many fail to grasp how difficult it is for people with epilepsy to establish social connections, form partnerships, start a family, or even find employment."<br>Bosnia, Caregiver (non-complex epilepsy), Participant 1<br><br>"My supervisor knew my condition, but they didn't care that I had tried my best to do my work and still fired me." "<br>Hong Kong, Male, 1" | No corresponding quantitative item. | No integration required. |
| | Structural barriers to employment and retention | Barriers to employment for PWEs | "[Describing a human rights issue at work, where necessary accommodations were not being made] I worte to human rights about it and they wrote to the union and that's how it all started. They're fighting now...I haven't worked since… this March, it'll be two years. I just feel like what's the point of fighting anymore? But I wrote the grievances and I'll just see what happens after. I've been told it's going through arbitration now....Now I just…I don't care about the job anymore. I just want, excuse my language, them to get shit for treating people with disabilities how they do.""<br>Canada, Rura (SVD)l, Participant 2<br><br>""She wants to become like everyone else. She wants to be active like others. She wants to work and do a job like my sister, brother and other girls of my age. But she can't do much. She only has to do this. She had to drop out of school in class 10th as she started having more seizures. She has completed class 9th but for the job an individual should have completed 10th or 12th class. There are no jobs in the market, not even a peon for someone who has only completed class 8th.""<br>India, Caregiver (complex epilepsy, SVD), Participant 2<br><br>""It is frustrating to be limited in the things you can do yet you used to. Now most of my work is closer to home yet before I used to travel to [small urban city], even [larger urban city]. I think it makes you a bit angrier because your patience for things that annoy you becomes smaller.""<br>Uganda, Older (SVD), Participant 3" | **Complementary:** People with epilepsy who view the condition as a disability were significantly more likely than those who do not to report that they would like to have a stable income and learn how to manage their money so they can better provide for themselves (45% vs. 34%; OR = 1.9, $p < 0.0001$). | The survey findings indicating that people who view epilepsy as a disability are more likely to express a desire for stable income and financial independence are supported by the qualitative insights. Participants described significant barriers to employment and personal development, including lack of workplace accommodations, disrupted education, and limited job opportunities particularly for those who had to leave school early due to their condition. The emotional and practical consequences of these challenges were evident, with individuals expressing frustration, loss of motivation, and feelings of exclusion from economic life. More generally, across all interviews (i.e. those who did not identify as having a disability) we heard fewer accounts of employment-related issues, though some still reported lack of empathy and accommodations at work, and difficulty securing employment (i.e. professions where disclosing epilepsy immediately disqualifies an applicant such as law enforcement). However, among those who self-identified as having a disability, the condition was often more limiting and challenging in nature, as reflected in their accounts. It should also be noted that caregivers are significantly impacted. Their comments tended to focus more on the financial burden of care and the need for financial assistance, rather than directly expressing a desire for their own stable income. This highlights the broader economic strain experienced by both PWE and their families. |

| | | | | | |
|---|---|---|---|---|---|
| | | Barriers to employment for PWEs | "Support should be coming from the top echelons of government where programs are put in place to enable people living with condition to access small funding for example to open up small businesses that do not necessarily require us to do work that is risky or might result in harm either to ourselves or the people with whom will be delivering the service... We are equally, if not more, passionate about financial freedom than some of our able-bodied counterparts. Within normal savings and credit societies a person living with the condition is not likely to get a loan because there's a fear that given that they get seizures, they're not likely to repay loan amounts."<br>Uganda, Male, Participant 1<br><br>"when it comes to social inclusion, I don't seem much progress. Honestly it feels like epilepsy lives in a grey zone - a forgetton zone. For example, epilepsy excludes you from many public job competitions...in every competition for police positions, when you look at the list of medical conditions that disqualify candidates, epilepsy is almost always one of the first on the list under nervous system disorders...there isn't the same kind of inclusion policy that I see in other countries."<br>Brazil, Male, Participant 1 | **Complementary:** Respondents from low-income countries were more likely to agree that their country needs to recognize and defend the human rights of people with epilepsy (e.g., through laws, nationwide campaigns, etc.) compared to those from middle- and high-income countries (78% vs. 59% vs. 42%; OR = 5.4, $p < 0.0001$). A similar pattern was observed in response to the statement "my country needs to protect the rights of people with epilepsy", with higher agreement among respondents from low-income countries (77% vs. 60% vs. 39%; OR = 7.3, $p < 0.0001$).<br><br>Additionally, female respondents were significantly more likely than male respondents to agree that their country needs to recognize and defend the human rights of people with epilepsy (e.g., through laws, nationwide campaigns, etc.) (47% vs. 57%; OR = 0.7, $p < 0.0001$). | Respondents from low-income countries were more likely than those from middle- and high-income countries to agree that their country needs to recognise and defend the rights of people with epilepsy. Female respondents were also more likely than male respondents to endorse this need.While this pattern was less distinct in the qualitative data, participants across settings and genders described exclusion from certain jobs, a lack of workplace accommodations, stigma from employers, and barriers to self-employment such as restricted access to loans. These accounts reflect a widespread experience of employment discrimination and structural barriers, complementing the survey's indication of a strong rights-based need, even if the same geographic and gender differences were not as prominent in interviews. |

| | | | | | |
|---|---|---|---|---|---|
| | Everyday challenges of epilepsy in the workplace | Disclosure dilemma *and* Workplace stigma and misconceptions | “My mistakes in my work performance will reveal my [diagnosis]...I feel embarrassed, worried and anxious about it.”<br>Hong Kong, Male, 2<br><br>"Mostly I don't want to tell anyone at work about my illness because the idea they have about me is that when I lose consciousness it is a condition related to diabetes...What if they knew about it? Maybe they would fire me from work because of the illness."<br>Tunisia, Older, Participant 5<br><br>"...at work [he] does not say that he has epilepsy because the second time he had a seizure, he was fired from work..."<br>Spain, Caregiver (non-complex epilepsy), Participant 3<br><br>"...I was with [a government official] as the communication officer...unfortunately I had an epileptic seizure. As soon as I fell, one of my colleagues mentioned that I was covered with sand...I overheard someone say 'He knows he is living with this condition, so why did he come to such places? He should have stayed in one place and allow others to work'."<br>The Gambia, Rural, Participant 2<br><br>"Unfortunately...in the last [job interviews], I was taking different pathways, because I asked myself, “Well, what do I do, do I say it or not during the next interview”. Because I went to a lot of interviews. I said, “Well, the next one I am going to say it in the interview. If you want me like this, that is, as I am, take it, and if no, not do it”. When I tried this pathway, I do tell them that this would come up in the medical assessment, that I had brain surgery, and they did not hire me."<br>Argentina, Rural, Participant 3<br><br>"At my old job, I was afraid that I would be labelled because of my diagnosis..."<br>Croatia, Male, Participant 2 | **Complementary:** Respondents from low-income countries were significantly more likely than those from middle- and high-income countries to report that they need to feel more included in the workplace and be able to build relationships (55% vs. 24% vs. 19%; OR = 10.9, $p < 0.0001$) as well as need advice on how to tell key people in their life about their epilepsy (e.g., about their diagnosis, how it affects their life) (68% vs. 25% vs. 27%; OR = 7.9, $p < 0.0001$). | Survey findings showed that respondents from low-income countries were significantly more likely than those from middle- and high-income countries to report needing greater inclusion and relationship-building in the workplace, as well as advice on how to talk to key people in their life about their epilepsy. While these needs were most strongly expressed in low-income settings in the survey, qualitative accounts indicated that concerns about workplace inclusion, disclosure, and fear of stigma were common across all contexts. Participants described anxiety about being misunderstood, dismissed, or treated differently after a seizure, reflecting both the immediate challenges of managing epilepsy at work and broader worries about social acceptance. Disclosure was discussed in multiple settings, with middle- and high-income participants more likely to raise it in relation to schools, workplaces, and friendships, with low-income participants more often framing it in romantic contexts. |
| | | Impact of epilepsy on work performance | "My sickness really affected my mental health. At any time I have an attacked, it will take two days before I leave my house to go to my business."<br>The Gambia, Older, Participant 5<br><br>"I have lower vitality because of the seizures my energy levels are lower than I’d typically expected to be. My ability to work is significantly reduced because of having epilepsy. I’m a barber but still I don’t get as much work as I would have if I didn’t have the condition."<br>Uganda, Male, Participant 1 | No corresponding quantitative item. | No integration required. |

| | | | | | |
|---|---|---|---|---|---|
| | | Workplace pressure and epilepsy management | " I think that I am a mildly anxious person at heart, it manifested itself more with my job which was stressful for me and that job was a trigger. I think I experienced burnout."<br>Croatia, Male, Participant 2<br><br>"When I got an apprenticeship as an office trainee. I got it in a place where I got to work under a boss who managed to destroy a lot of people. He has today gone bankrupt. When I was employed there, I had a lot of seizures, as I was under a lot of pressure both mentally and physically. With long working days and nose no breaks. So, there I was maxed out."<br>Denmark, Rural, Participant 1 | No corresponding quantitative item. | No integration required. |
| | Health-related mobility challenges | Physical strain of travelling | “I normally have to have scans before… that would be another trip… It really took me up to take pretty much three days off work.”<br>Australia, Rural, Participant 1<br><br>"I need to be well-rested and I don't travel alone. On longer trips, my wife and I take turns so that we don't drive for long periods at a time. I can only drive for three hours at a time, and it's becoming increasingly difficult."<br>Croatia, Male, Participant 2<br><br>"Just going to a big hospital in the provincial capital involved a two-hour train ride, leaving me feeling completely drained and increasingly tired. After getting off the train, I still had to take another vehicle to the hospital. By the time I arrived, I felt like all my energy was gone, and I didn't even want to talk."<br>China, Older, Participant 5 | No corresponding quantitative item. | No integration required. |
| | | Public transport safety concerns | “Actually, I don’t drive, so I only use public transportation. I was a little worried about having a seizure while using public transportation at first.”<br>South Korea, Male, Participant 1<br><br>“I really wanted to go to Japan in the winter, but now that I think about it, I'm a little scared. Because if I fall down on the street, can these people help me?"<br>South Korea, Rural (female), Participant 5 | **Complementary:** Women were more likely than men to express a need to 'feel safe when travelling to different places' (46% vs. 38%; OR = 1.4). | While survey findings indicated that women were more likely to report a need for safety while traveling, qualitative insights from both men and women show that concerns about seizure-related risks while travelling on public transport are widely shared. |
| | | Driving restrictions due to diagnosis | "...for me, that was the biggest down. Down issue was the fact I lost my driver driver's license last Monday. Lost my independence, my mobility and again after 60 years of driving at will anywhere kind of saying it really hit me like a hammer blow."<br>Canada, Older, Participant 4<br><br>"my father for example just they changed his tablet so he had to stop driving for six months until they were sure that it was controlled again so he wasn't allowed to allowed to drive again"<br>Australia, Male, Participant 3" | **Complementary:** PWE are generally less likely to select needs compard to CG (OR = 0.45-0.7, p<0.0001). There is one exception in the survey data. PWE were significantly more likely than caregivers to report that driving restrictions in their country limit their independence (34% vs. 24%; OR = 1.9, p < 0.0001). | The survey showed that PWE are more likely than caregivers (answering on behalf of PWE) to feel that driving restrictions limit their independence. Qualitative interviews add depth to this finding, revealing how the loss of a driver’s licence can feel like a profound personal blow impacting not just mobility, but autonomy, and daily freedom. Together, the findings highlight how epilepsy-related restrictions can extend far beyond safety, shaping a person’s sense of self and independence. |

| | | | | | |
|---|---|---|---|---|---|
| | | Bureaucratic hurdles in transport support | “We lose a lot of time on administrative tasks, such as obtaining transport certifications, which can be frustrating.”<br>Bosnia & Herzegovina, Caregiver (non-complex epilepsy), Participant 1<br><br>"I can't drive like the only transportation I have is the public transportation is my bike. And if the government covered public transportation for people that have epilepsy like that would be a bonus. And that's one of the biggest things that I struggle with is the public's transportation. You know, I mean, I can't file like, the receipts that I spend on public transport. And that's kind of like one of the things that would benefit. Everyone as a whole with epilepsy."<br>Canada, Male, Participant 5 | No corresponding quantitative item. | No integration required. |
| | | Cost of barriers to transport services | "Where I live it’s very far from the hospital and I must trek. when I go to the hospital, I must buy more medication at the pharmacy because the hospital doesn’t have the [medication]. Medication and transportation are challenges due to my limited source of income."<br>The Gambia, Older, Participant 5<br><br>"Not everyone in my community can afford it or is able to travel to [to capital city for appointments / scans]."<br>Uganda, Rural, Participant 2 | No corresponding quantitative item. | No integration required. |
| | | Inadequate accessible transportation infrastructure | “Transportation and public services are not adapted for people with disabilities or difficulties… I mostly have to drive him everywhere…”<br>Bosnia & Herzegovina, Caregiver (complex epilepsy), Participant 1<br><br>"Me living in a small town, we didn't have that kind of public transportation, but for people…living in like the bigger areas…they do have that public transport that they can rely on..."<br>USA, Male, Participant 4 | No corresponding quantitative item. | No integration required. |
| | Transport accessibility and infrastructure gaps | Inadequate accessible transportation infrastructure | "...if we have to go a little further out into the outlying districts, there are simply no options with public transport. It's as bad as it can be."<br>Denmark, Rural, Participant 1<br><br>"Also, transportation was difficult for us, as we had to travel long distances to reach urban areas to obtain transportation to keep medical appointments and obtain medication."<br>Tunisia, Caregiver (complex epilepsy), Participant 4<br><br>"That was probably the worst thing I think being in a rural area, we don't have public transport...the main trains only go three or four times a day and we only have a connecting bus from [location] to that train. And our next biggest town is [location - suburban], which is the population of probably 20,000.<br>But that's an hour away. So really that was the worst thing."<br>Australia, Rural, Participant 2 | **Complementary:** People living in rural areas were significantly more likely to report needing support to travel to work and earn a living compared to those in suburban or urban areas (33% vs. 27% vs. 25%; OR = 1.6, $p < 0.0001$). | People from rural areas were significantly more likely to report needing support to travel for work and income. While only one interview directly linked poor transport to work, interviews across all country income groups revealed widespread 'rural' transport challenges highlighting the need for improved infrastructure for PWE. |

| | | | | | |
|---|---|---|---|---|---|
| | | Inadequate accessible transportation infrastructure | "It definitely depends on where you live. Me living in a small town, we didn't have that kind of public transportation, but for people I do know that living in like the bigger areas, bigger cities, they do have that public transport that they can rely on, but it can definitely be hard for people in my situation living in smaller towns who may not have the type of family support that I was lucky enough to have."<br>USA, Male, Participant 4<br><br>"We've made some adjustments to suit her needs. For instance, when she cannot open a food package, she uses scissors to access the contents. Unfortunately, there aren't many conveniences available regarding transportation. While some buses have ramps, they aren't present in all buses or at all stops. Additionally, marked parking spaces exist but are often disrespected by others."<br>Bosnia & Herzegovina, Caregiver (non-complex epilepsy), Participant 1<br><br>"However, in [urban city], transportation and public services are not adapted for people with disabilities or difficulties. I mostly have to drive him everywhere, which creates significant challenges in my daily activities. For instance, I work from 8 AM, and the school starts at the same time, so I'm always about 10 minutes late for work every morning."<br>Bosnia & Herzegovina, Caregiver (complex epilepsy), Participant 2 | People who view epilepsy as a disability were significantly more likely than those who do not to report wanting public transport to be easier to access (e.g., through special fares, lifts, ramps etc.) (39% vs. 28%; OR = 1.8, $p < 0.0001$). | The survey finding that people who view epilepsy as a disability are more likely to want easier access to public transport is supported by qualitative accounts highlighting significant gaps in transport infrastructure and accessibility. Participants described practical challenges such as the absence of ramps, limited transport services, and poorly adapted public infrastructure. These issues are often exacerbated for those living in rural areas or smaller towns, where services may be more limited (e.g. buses stopping earlier in the evening). For people with epilepsy who also face driving restrictions, these transport barriers further compound their limitations, increasing reliance on CGs and adding stress to daily routines. The data reinforce the need for more inclusive, reliable, and flexible transport options that account for both geographic and disability-related challenges. |
| | | Evidence of misinformation | "At first [participant learned about epilepsy] in a chaotic way, searching on Google, searching on websites which I don't think they are always reliable."<br>Argentina, Male, Participant 2<br><br>"My [son] also searches for information about epilepsy on the internet at home. But the information on the internet is chaotic, with both true and false. The doctor told him not to look at those messy things casually and to find information from regular official accounts and video channels."<br>China, Caregiver (non-complex epilepsy) Participant 3<br><br>"videos on the Internet show that when seeing an epilepsy patient having a seizure, people put a towel or glove in the patient's mouth and pinch the philtrum. This incorrect knowledge is misleading the public."<br>China, Rural, Participant 1 | No corresponding quantitative item. | No integration required. |
| **4. Consequences of inaccessible or inadequate information** | Health information shortfalls in healthcare | | | | |

| | | | | | |
|---|---|---|---|---|---|
| | | Extensive burden to access epilepsy knowledge | "When he was one year old that's when I witnessed him having seizures. In that situation, I tried to seek help from people so that he'll be rushed to the health centre at [town]. The healthcare professional only prescribed medication like carbamazepine, but I was not informed that it was epilepsy."<br>The Gambia, Caregiver (complex epilepsy), Participant 3<br><br>"Previously we were given medication but without explicit guidelines on never changing or sharing medication, timeliness of dosage."<br>Uganda, Male, Participant 1<br><br>"I've noticed that doctors… are often not very well-informed about epilepsy. Most of my knowledge comes from my personal experience and research."<br>Bosnia & Herzegovina, Male, Participant 1<br><br>"...they should have more information to tell us about treatment of seizures to tell us and also they should know and tell us more about the management of seizures. I feel that someone should tell me how to take medicine, for how long should I continue it and how will I become alright but no one says so. All of them just say, 'You have to take medicine, you have to take it compulsorily."<br>India, Rural, Pariticpant 2 | **Complementary:** Respondents from low-income countries were significantly more likely to report needing access to high-quality information about epilepsy (e.g., seizure control, triggers, treatment options, side effects) compared to those from middle- and high-income countries (77% vs. 46% vs. 35%; OR = 7.0, $p < 0.0001$). | The high demand for better information among respondents from low-income countriesis reinforced by interview accounts that highlight serious gaps in accessing even the most 'basic' medical information such as receiving a diagnosis without explanation or being prescribed medication without clear instructions. In contrast, interviews from higher-income countries more often revealed concerns around more advanced / nuanced information needs, such as support for emotional wellbeing or being signposted to patient advocacy groups. These findings suggest that while epilepsy information gaps are a global concern for PWE, their nature and depth vary by context ranging from fundamental communication failures in low-resource settings to unmet needs around holistic care in higher-resourced ones. |

| | | | | | |
|---|---|---|---|---|---|
| | | HCPs miss educational opportunities | “I wish the medical staff could have a moment to exchange detailed information about the patient's illness. I can't even ask questions properly right now. Rather, when I try to speak as a patient carer, he sighs deeply and stops me from asking questions.”<br>South Korea, Caregiver (complex epilepsy), Participant 1<br><br>"When he was one year old that’s when I witnessed him having seizures. In that situation, I tried to seek help from people so that he’ll be rushed to the health centre at [town]. The healthcare professional only prescribed medication like carbamazepine, but I was not informed that it was epilepsy."<br>The Gambia, Caregiver (complex epilepsy), Participant 3<br><br>"Previously we were given medication but without explicit guidelines on never changing or sharing medication, timeliness of dosage."<br>Uganda, Male, Participant 1<br><br>“I’ve noticed that doctors… are often not very well-informed about epilepsy. Most of my knowledge comes from my personal experience and research.”<br>Bosnia & Herzegovina, Male, Participant 1<br><br>"[Speaking about other PWE and knowing the cause of epilepsy] ...and they don't know that...I tell them that they ask their doctors. Nothing, [doctors] don't inform, they don't inform them why. If I have stayed with the previous clinic, it was also the same. That's why I didn't want to stay. I wanted to continue understanding and looking for the why, what causes it for me."<br>Argentina, Rural, Participant 3<br><br>"...they should have more information to tell us about treatment of seizures to tell us and also they should know and tell us more about the management of seizures. I feel that someone should tell me how to take medicine, for how long should I continue it and how will I become alright but no one says so. All of them just say, 'You have to take medicine, you have to take it compulsorily."<br>India, Rural, Pariticpant 2 | **Complementary:** Respondents from low-income countries were significantly more likely than those from middle- and high-income countries to report that they need advice on how to have useful conversations with doctors and nurses (69% vs. 27% vs. 25%; OR = 7.9, $p < 0.0001$). | The survey finding that people in low-income countries are significantly more likely to want advice on how to have useful conversations with doctors and nurses is supported by qualitative insights. While there is no direct reference to the dialogue between PWE / CGs and HCPs in the interviews with participants from low-income countries, they do highlight how a lack of 'access' to meaningful dialogue with HCPs can result in relying on one's own expereinces to learn and understand their epilepsy. In these settings, access to HCPs itself was often limited (time and resource for long journeys frequently mentioned) leaving little opportunity to focus on the 'content' of the conversations. Across income groups, especially in rural areas, a similar challenge appears - not only about 'what' is said, but whether there is an 'accessible HCP (often specialist)'. In other income settings, missed opportunities 'within' the consultations were more commonly described. |

| | | | | | |
|---|---|---|---|---|---|
| | | Knowledge gaps in HCPs | "...no other neurologist could understand... None of them... I went to a neurologist who asked, 'What is it? Tell me, what is it?' He wanted me to tell him what epilepsy is."<br>Brazil, Rural, Participant 3<br><br>"...epilepsy is considered [by HCPs] to be only an irregularity in the electrical functioning of the brain...only a biological one and nothing else. And it despises the fact that we are holistic integral beings, and that I also have [seizure] when I fight with.... when I got divorced for example. Or, when they didn't pay me my salary...made me worry and I have a [seizure]."<br>Argentina, Male, Participant 1<br><br>"I also believe that emotional changes after a seizure need more attention...I think this kind of information needs to be discussed more openly and made clearer to patients. I believe that doctors and nurses are taught how to manage seizures during their training, but these emotional and psychological aspects seem to be overlooked."<br>Brazil, Male, Participant 1<br><br>"The doctor said a lot of things I wasn't allowed to do. But I didn't want to put my life on hold either."<br>Denmark, Male, Participant 4 | **Divergence:** Respondents from low-income countries were more likely than those from middle- and high-income countries to report that doctors and nurses need to know more about people's wider needs linked to epilepsy (e.g., emotions and mental health needs of people with epilepsy at different stages of life) (77% vs. 46% vs. 49%; OR = 4.4, $p < 0.0001$). The need for 'doctors and nurses to know more about people's wider needs' was the third most-selected need within Domain 3. | While survey results showed that respondents from low-income countries were more likely to report that doctors and nurses need to better understand people's wider needs linked to epilepsy, interviews revealed what constitutes 'wider needs' may differ by context. Participants from low-income countries tended to focus on treatment-specific gaps (e.g. availability of medication), follow-up care, or basic clinical communication, rather than emotional or mental health needs. In contrast, participants from middle- and high-income countries more frequently mentioned the psychological and emotional dimensions of living with epilepsy. This difference may reflect variations in healthcare infrastructure, where in low-income settings the absence of basic medical support overshadows more nuanced psychosocial concerns. Together, these findings suggest that widening the clinical lens to include emotional wellbeing should be an approach adopted in all settings. |
| | Knowledge gaps in everyday and future planning | Feelings of fear and worry arising from knowledge gaps | "... in the early stages, you have the seizure presented at A&E, you have to wait for a neurologist. So that's a bit of the unknown because you don't know. But now that they've put a diagnosis to it, you know what you're working with, I think. But there's still the unknown of maybe it was caused from something else. But I think once yeah, it's the same thing. Once you get a name and treatment, you can work with it."<br>Australia, Rural, Participant 4<br><br>"talking about it and sharing my experiences with [PO] members...It has helped me become less anxious and less ashamed of having this condition. I have met members whose challenges exceed mine by far and its these conversations that give all of us hope that we can make our situation better in the community."<br>Uganda, Rural, Participant 5 | No corresponding quantitative item. | No integration required. |
| | | Knowledge gaps in PWE | "My head keeps spinning. This time I think it is happening to me due to the cold weather. I doubt that as I feel cold, maybe because of that it happens. I can't handle the cold weather. I can handle heat but I cannot handle winter at all. Maybe, my head spins because of the cold."<br>India, Older (male), Participant 2<br><br>"She is so one of her triggers is thermosensitivity anything about anything above 75°? So, anything anything above 75° is is too hot for her...if it gets too hot in here, like during the summers, we really can't go out like here. It's OK, but in [different warmer state] where it was like 116°, she was stuck indoors"<br>US, Caregiver (non-compex epilepsy, male), Participant 5" | **Divergence:** Female respondents were significantly more likely than male respondents to report that there needs to be advice on how to cope with changes in weather that may impact people with epilepsy (e.g., extreme heatwaves) (35% vs. 42%; OR = 0.7, $p < 0.0001$) | While survey findings showed that women were more likely to express a need for advice on coping with weather-related changes that affect epilepsy, this issue was largely absent from the qualitative interviews. Notably, however, extreme changes in temperature were more commonly mentioned by caregivers of people with epilepsy, suggesting second-hand recognition of environmental impacts on seizure activity. The lack of direct discussion from individuals with epilepsy may reflect limited awareness or underestimation of these risks. This divergence underscores the importance of raising awareness about weather-related seizure triggers, particularly in the context of climate change, where rising global temperatures and extreme |

| | | | | | |
|---|---|---|---|---|---|
| | | | | | weather events are likely to pose increasing risks for people with epilepsy. |
| | | Knowledge gaps in caregivers | "Family members can help by learning about epilepsy, understanding the individual's needs, and being prepared to assist during a seizure."<br>China, Male, Participant 2<br><br>"Having more understanding and support from family, friends, and the community can make relationships better. This might include learning more about epilepsy, being patient, and offering emotional support and financial support."<br>India, Caregiver (non-complex epilepsy), Participant 2<br><br>"I got the impression from those meetings that there's still not enough [resources]. Like there's a lot of very, very confused caregivers... there's not enough awareness."<br>Canada, Caregiver (non-complex epilepsy) Participant 3" | **Divergence:** Caregiver respondents from low-income countries were significantly more likely than those from middle- and high-income countries to report that they need training for caregivers on epilepsy management (88% vs. 32% vs. 28%; OR = 20.2, $p < 0.0001$). | While CG respondents from low-income countries indicated a strong perceived need for CG training in epilepsy management, this need was not explicitly voiced in the qualitative data. Instead, participants across all income settings spoke more generally about gaps in knowledge and support within families and communities. References to caregiver needs were indirect and lacked a clear call for structured training. This divergence suggests that while caregiver training may emerge as a priority when prompted in surveys, it is not yet fully recognised or articulated in everyday narratives possibly reflecting limited awareness of what such training involves or how it could help. |
| | | Low awareness of government support | “As far as I know, such services [government support or resources for PWE] either don’t exist or are very rare. Maybe some programs do exist, but even if they do, they are not sufficiently accessible to people. Even if these programs exist, they need to be better promoted so that people know about them. Currently, even if there are programs for people with epilepsy, individuals must research and discover them on their own, which is a big problem.”<br>Bosnia & Herzegovina, Male, Participant 5 | No corresponding quantitative item. | No integration required. |
| | | Unfamiliar with epilepsy patient organisations | "I've never been to those support groups and those gatherings in the city for Epilepsy Day. I'm not involved in that. Maybe it would be good to somehow enable contact with other sufferers…I don't know their names. I didn't do any research on my own". | No corresponding quantitative item. | No integration required. |
| | | False cultural beliefs | "People usually don’t want to visit because they find the situation scary or even disgusting. Some people actually think that we offered her up as a sacrifice.”<br>Uganda, Caregiver (complex epilepsy), Participant 2<br><br>“Some friends directly blocked him on [social media] and deleted his phone number as soon as they heard he had this disease. It's obvious that they are distancing themselves from him. They think he has a bad disease and are afraid of being affected, so they don't want to have anything to do with him anymore.”<br>China, Caregiver (non-complex epilepsy), Participant 3 "<br><br>" She also says to make the person having a seizure smell onion to stop his/her seizure and make them fine. I had seen her making that kid smell onion when she was having a seizure...someone from the community might have told her"<br>India, Rural, Participant 2 | **Complementary:** Respondents from low-income countries were significantly more likely than those from middle- and high-income countries to report that their local community needs better information to get rid of old, incorrect beliefs about epilepsy (80% vs. 53% vs. 50%; OR = 5.5, $p < 0.0001$). | Respondents from low-income countries were significantly more likely than those from middle- and high-income countries to report that their local community needs better information to get rid of outdated or incorrect beliefs about epilepsy. Qualitative accounts confirmed that such beliefs exist across locations, particularly in low- and middle-income settings, but the ways they affect people with epilepsy vary. In all contexts, participants described impacts such as fear-based distancing and social isolation. However, in low-income countries, these misconceptions were more often linked to severe consequences, including violence and the prioritisation of herbal or spiritual remedies over medical treatment. |

Widespread misunderstanding of epilepsy

| | | | | | |
|---|---|---|---|---|---|
| | | Knowledge gaps in families | "Their father (respondent's husband) doesn’t allow me to travel alone. He tells me not to go close to roads as I might start having pain though I have not had a seizure for a long time. There is still fear in his heart and says I shouldn't go anywhere alone."<br>India, Rural, Participant 2<br><br>"I don't have a job. My family is in the countryside and they don't allow me to do physical work."<br>China, Rural, Participant 1<br><br>"it has happened to me with direct, close family that they say: “No, no. Don’t go, don't leave me alone because I don't know what to do....Another incident was of a child who was sitting her primary leaving exams. When the child didn’t perform to expectation the parents got really ashamed of her performance. They halted her education because of that. She thereafter requested that she be allowed to take up tailoring and was denied saying she might make a repeat of her earlier disappointment. She once went for burial and got a seizure so when more recently she lost an uncle she was denied the opportunity to go for the burial again for fear of a repeat."<br>Uganda, Caregiver (non-complex epilepsy), Participant 4 | **Complementary:** Respondents from low-income countries were significantly more likely to report that their family and those who care for them need to learn more about epilepsy (e.g tips to help me stay safe, seizure first-aid, seizure control), compared to those from middle- and high-income countries (80% vs. 34% vs. 29%; OR = 11.7, $p < 0.0001$). | In low-income countries, the survey revealed a strong need for families to better understand epilepsy, and this was reinforced in interviews that showed how limited knowledge often manifests as overprotection, fear, and restriction. Rather than enabling, this gap in understanding can undermine the independence and autonomy of people with epilepsy, particularly in settings where stigma and misinformation persist within the family context. While such dynamics were observed across all income settings, interviews suggested they were more pronounced and deeply rooted in low-income and middle-income contexts. |
| | | Knowledge gaps at work | "Now that the end of the year is approaching... we have to celebrate everything, drink as much alcohol as possible, to be with as many people as possible, go to places, to bars, to places that are hyper stimulating. This ones bothers me, when there are many sounds, many noises... I get desperate, and I need to leave."<br>Argentina, Male, Participant 2<br><br>"I find it doesn’t matter where you work, people just don't understand disabilities when they can't see them...It's the information, it doesn't get out properly.... And employers do not understand disabilities are epilepsy because a lot of epileptic people do not express themselves because they not might be as bad as you know mine or, they might experience a different way."<br>Canada, Rural, Participant 2 | No corresponding quantitative item. | No integration required. |
| | | Knowledge gaps in educational settings | "the teachers do not know CPR for epilepsy. In fact, I thought many times... to give a talk on CPR for epilepsy, but it's kind of complicated bureaucratically. Not because the school doesn't want to - the school is excellent; I don't have anything bad to say; from the leadership to the one who opens the door to you. Their support to me and my little boy ticks all the boxes - but it is a reality that no, they do not have that knowledge [CPR for epilepsy}. They do not have it."<br>Argentina, Caregiver (complex epilepsy), Participant 1<br><br>"There is no general protocol for emergencies where some children in an educational institution would be given medicine to stop a seizure."<br>Croatia, Caregiver (complex epilepsy), Participant 1<br><br>"Imagine that in my daughter's school, the teacher did not understand anything, she knew absolutely nothing, and I thought: “How can I leave my daughter with you, if you don't know how to help her?” So, it was good to sit down, talk to them, explain to them, to provide them with tools, because society does not have knowledge."<br>Argentina, Caregiver (non-complex epilepsy), Participant 5 | No corresponding quantitative item. | No integration required. |

| | | | | | |
|---|---|---|---|---|---|
| | | Knowledge gaps in healthcare | " I believe that doctors and nurses are taught how to manage seizures during their training, but these emotional and psychological aspects seem to be overlooked."<br>Brazil, Male, Participant 1<br><br>"I keep saying it everywhere, I think there is a lot of research missing in the subject. Pharma needs to take responsibility, to really mention all the side effects that we have, and that research goes further. Because it seems that we only have [medicines] to avoid the seizures and that's it, and that's a success. But what happen with the sleep that medication produces in us and the irritability that medication produces in us. And the stomach problems caused by the medication, or weakness or others. It seems that they do not exist. In truth that makes me angry."<br>Argentina, Male, Participant 2 | No corresponding quantitative item. | No integration required. |
| | | Knowledge gaps in society | "…there is a lack of knowledge. Most often, people think, when you have epilepsy, you fall over and convulse.”<br>Denmark, Caregiver (complex epilepsy), Participant 4"<br><br>“I think society is unaware. They should at least know how to react when a person has a seizure and, of course, recognize the symptoms when it first occurs. People are not educated about epilepsy and often do not take it seriously, not realizing that it can lead to loss of life.”<br>Bosnia & Herzegovina, Older, Participant 3<br><br>“Some friends directly blocked him on [social media] and deleted his phone number as soon as they heard he had this disease. It's obvious that they are distancing themselves from him. They think he has a bad disease and are afraid of being affected, so they don't want to have anything to do with him anymore.”<br>China, Caregiver (non-complex epilepsy), Participant 3 " | No corresponding quantitative item. | No integration required. |

| | | | | | |
|---|---|---|---|---|---|
| **5. Complex epilepsy needs demand more than standard approaches** | Access to epilepsy care | Barriers to accessing care and information | "...my biggest frustration and disappointment in this entire process other than just the shit show of the diagnosis was the the amount of work I had to do as an advocate to get his needs met. That should have been much more quick and fluid... first up, he has a seizure, you bring him to the family doctor...[who] refers to the only paediatric neurologist in the city, which is a seven week wait...then she says, 'oh, this is a complicated case. You need to go to [another location], and then it's a six month wait. So you wait again. Just get passed along. There's limited communication between everyone...we actually took [PWE] twice now over to [US] for care to skip the six month wait times down there for testing. So silos for sure. Lack of communication. Each has their own purpose but definitionely not a lot of cross communication<br>Canada, Caregiver (complex epilepsy), Participant 1<br><br>"Accessing rare epilepsy services is difficult in urgent cases you want for specialist doctors to come to you to examine the patient and solve his problem."<br>Tunisia, Caregiver (complex epilepsy), Participant 1<br><br>Unfortunately, not all medications available in the EU are available in the Croatia. Some medications are on the list, but you can't get them without medical committees [otherwise] not prescribed because doctors don't want complications. That's completely bad."<br>Croatia, Caregiver (complex epilepsy), Participant 1 | **Convergent:** People affected by complex epilepsy were significantly more likely than those affected by non-complex epilepsy to report that there is a need for easier access to 'healthcare services or doctors and nurses who treat epilepsy' (66% vs. 55%; OR = 1.8, $p < 0.0001$), 'complex epilepsy services' (53% vs. 38%; OR = 1.9) and 'health care that addresses their own specific needs (e.g., having a personal care plan) (47% vs. 37%; OR = 1.6, $p < 0.0001$). | Survey and interview data both highlighted the greater challenges faced by people with complex epilepsy in accessing tailored, coordinated care. Participants described fragmented health systems, poor infrastructure, and a lack of specialist input, which often left caregivers to take on the burden of navigating healthcare alone. Across settings, these barriers were compounded by difficulties obtaining clear, relevant information, limited continuity of care, and poor communication between providers. Such gaps in coordination and support contribute to delays in treatment, unmet care needs, and increased strain on families, underscoring the need for more integrated and specialist-led care pathways. |

| | | | | | |
|---|---|---|---|---|---|
| | | Barriers to accessing care and information | "The silo here is that, first up, he has a seizure, you bring him to the family doctor, the family doctor refers to the only paediatric neurologist in the city, which is a seven week wait, and you go and you see her. And then she says, oh, this is a complicated case. You need to go to [location] and then it's a six month wait. So you wait again. Just get passed along. There's limited communication between everyone and then in the meantime, for us anyway, we're a border city to [neighbouring country]. So we actually took [our son] twice now over to [neighbouring country] for care to skip the six month wait times down here for testing."<br>Canada, Caregiver (complex epilepsy), Participant 1<br><br>""Unfortunately, not all medications available in the EU are available in the Republic of Croatia. Some medications are on the list, but you can't get them without medical committees. Automatic medications that go to committees are not prescribed because doctors don't want complications. That's completely bad.""<br>Croatia, Caregiver (complex epilepsy), Participant 1<br><br>""Medication outages, when I go to the hospital and there is no medication, I run the risk of getting a seizure, to or from the hospital. There are extremes where I go even two months without the medication being available.""<br>Uganda, Male, Participant 1" | **Complementary:** Respondents who identified as belonging to a minority group were significantly more likely than non-minority respondents to report needin ongoing access to treatment (55% vs. 47%; OR = 6.2, p < 0.0001), and tailored healthcare (46% vs. 37%; OR = 7.3, p < 0.0001). | Across Domain 3 (Healthcare and wellbeing), survey respondents from minority backgrounds were more likely to report unmet needs across nearly all items. The most significant findings indicate consistent disparities in access to rare or complex epilepsy services, ongoing access to treatment,and receiving tailored healthcare. While interview participants did not explicitly self-identify as belonging to a minority group, we explored the experiences of those who described exclusion or rejection directly linked to their epilepsy. What unites these accounts is not ethnicity, migration status, or cultural identity, but the social marginalisation that arises specifically because of epilepsy. As such, the healthcare barriers described long wait times, lack of tailored care, medication shortages, and systemic inefficiencies are best understood as consequences of epilepsy-related exclusion. While this does not offer a complete picture of all minority group experiences (e.g., it does not capture the impact of language barriers or cultural dissonance that may affect some ethnic minority or immigrant groups), it does offer insight into how epilepsy itself can lead to 'minority-like' status in the healthcare system. |
| | | Quality of medical practice | "When they go to a paediatrician and mention that their child has epilepsy or intellectual disability, they often just throw their hands up and say they can’t help and give up from the start.”<br>Bosnia & Herzegovina, Caregiver (complex epilepsy), Participant 2<br><br>"The communication between the doctor and the patient is very one-sided. So when the doctor says he's in a bad mood that day, he sometimes fails to ask me any questions he has prepared for months."<br>South Korea, Caregiver (complex epilepsy), Participant 1 | No corresponding quantitative item. | No integration required. |

| | | | | | |
|---|---|---|---|---|---|
| | Cognitive, developmental and emotional needs | Cognitive, developmental and extreme care needs | "When her condition was severe before she was two years old, it had a great impact on her development. She was slower in development than her peers. Other children at that age might have already been able to walk and talk, but she couldn't...after entering junior high school, due to the frequent seizures she had before the age of two, which caused certain damage to her nervous system, she is relatively weaker in some subjects that require stronger thinking abilities, such as mathematics in science."<br>China, Caregiver (complex epilepsy), Participant 4<br><br>"ever since he's been having so many episodes of seizures, it has affected his speech. The only thing he's able to do sometimes is make murmurs or gestures that show he's complaining about something or that he needs you to attend to his needs."<br>The Gambia, Caregiver (complex epilepsy), Participant 3.<br><br>"We treated her with a lot of changes. She had frequent seizures, so we called 119 more than 60 times every six months."<br>South Korea, CGC, Participant 1<br><br>"She needs help with almost everything, both personal care and getting food and remembering her medication… to get dressed and all that kind of stuff."<br>Denmark, CG, Participant 1<br><br>"she picks her skin non-stop. You literally can't leave her for more than two minutes by yourself...They can't do anything themselves. I mean, the only thing they could do themselves is get their iPad and use it. Everything else they rely on people."<br>Australia, CGC, Participant 1 | No corresponding quantitative item. | No integration required. |

| | | | |
|---|---|---|---|
| | | Emotional and behavioural challenges | “...any time he has an epilepsy attack, it takes a number of days for him to be himself... His moods or reaction to actions are unpredictable.”<br>The Gambia, Caregiver (complex epilepsy), Participant 3<br><br>"“She gets very irritable when there are people around that she doesn’t know.”<br>Uganda, Caregiver (complex epilepsy), Participant 2<br><br>“He’s very attached to his routine, so it is hard for him to deal with something small that isn’t his routine...”<br>Argentina, Caregiver (complex epilepsy), Participant 4<br><br>“Her anxiety is so high because she remembers her cramps...She’s so scared of having another seizure, she refuses to step away from me at all.”<br>South Korea, Caregiver (complex epilepsy), Participant 1<br><br>“I always tell him to let me know. But now, he does not tell me as he remembers my scream that time and he doesn't want to [scared me]...”<br>Argentina, Caregiver (complex epilepsy), Participant 4<br><br>“The person feels nervous sometimes for fear that she will have seizures.”<br>Tunisia, Caregiver (complex epilepsy), Participant 1<br><br>"When she was a child, she was very afraid of going to the hospital. She would cry when she saw a white coat, and even refused to enter the consulting room. Especially when doing an electroencephalogram, those long wires made her very scared.”<br>China, Caregiver (complex epilepsy), Participant 4 |

| | | | | | |
|---|---|---|---|---|---|
| | | Gaps in specialist medical support | "[Discussing mental health] I don't think it's at the forefront of conversations. I'm just thinking of a few times, well, I was just thinking, has a paediatrician neurologist ever said are you taking care of yourself? They probably have, but haven't really been invested in the answer." Australia, Caregiver (complex epilepsy), Participant 3<br><br>"I have been through a lot – visiting various clinics and institutions in many countries, all related to his epilepsy and autism...I know many parents who have similar problems. When they go to a pediatrician and mention that their child has epilepsy or intellectual disability, they often just throw their hands up and say they can't help and give up from the start. Talking to people, I realized that this is truly a major issue in the approach to patients."" Bosnia & Herzegovina, Caregiver (complex epilepsy), Participant 2<br><br>"...I have so much printed stuff here. I've researched extensively about epilepsy...I also spoke with other mothers. I connected with parents from all over the world...I would get articles from them and learn as much as I could. I did all this so I could have informed conversations with doctors, to fight for what was best for her. Because it's important, there are so many different conditions, and sometimes doctors don't know everything about a particular one...it's crucial that caregivers and doctors can have proper discussions, working together to find the best approach for the child." Brazil, Caregiver (complex epilepsy), Participant 5<br><br>"No one is treating (PWE's) emotional state... in that area we are self-taught... I don't know who I have to speak to about that, maybe a child psychologist?" Spain, Caregiver (complex epilepsy), Participant 1 | **Complementary:** People affected by complex epilepsy were significantly more likely than those affected by non-complex epilepsy to report that they need help taking care of their other health problems along with their epilepsy (e.g., co-morbidity) (48% vs. 37%; OR = 1.6, $p < 0.0001$). People affected by epilepsy who view the condition as a disability were also significantly more likely than those who do not to report that they need help taking care of their other health problems along with their epilepsy (e.g., co-morbidity) (47% vs. 32%; OR = 2.0, $p < 0.0001$)." | The quantitative findings show that people with complex epilepsy and those who view their condition as a disability are more likely to need help managing other health problems. To explore these findings in the qualitative dataset, we reviewed all interviews with participants who self-identified as having a disability. Most were caregivers of individuals with complex epilepsy. While there were no direct references in the interviews to additional care reuired for co-existing conditions , we identified quotes in which caregivers described their experiences of trying to ensure all aspects of epilepsy including broader health needs were properly addressed. These participants described significant struggles navigating poorly coordinated care systems. Many spoke of the burden, frustration, and helplessness they experience when faced with HCPs who lack the knowledge or confidence to support co-existing needs. Some reported doing extensive personal research just to advocate effectively for their child, highlighting the lack of accessible, integrated care. Our interviews show that those with complex epilepsy and those that self-identify as having a disability are likely to have additional health challenges that are not adequately addressed, placing added responsibility on families already under significant strain. |
| | | Need for psychsocial and emotional support | "I talk to her more, even when she is sleeping, when she hears a familiar voice close by it calms her down." Uganda, Caregiver (complex epilepsy), Participant 2<br><br>"By loving and caring for her. It's always only who calms her down." India, Caregiver (complex epilepsy), Participant 2<br><br>"When she was a child, she was very afraid of going to the hospital... Especially when doing an electroencephalogram, those long wires made her very scared. Later, I changed the way I talked to her. I told her that these long wires were like turning her into a Rapunzel, so she gradually stopped being so afraid. When doing an MRI, the sound inside was very noisy, and she was very resistant at first. I told her that since she liked singing and music so much, she could imagine the sound of the MRI as the sound of different musical instruments and listen to it as a special kind of music. In this way, she gradually overcame her fear." China, Caregiver (complex epilepsy), Participant 4<br><br>"She is given medication for depression." Denmark, Caregiver (complex epilepsy), Participant 4 | No corresponding quantitative item. | No integration required. |

| | | | | | |
|---|---|---|---|---|---|
| | | Safety risks related to developmental needs | "If they get a banana, they could put the whole thing in their mouth and choke on it... that's for me, more of a safety concern than the seizures...SUDEP... it's always in the back of my mind... but choking is a bigger worry."<br>Australia, Caregiver (complex epilepsy), Participant 3<br><br>"he's very impulsive and he doesn't have a concept of like, safe surroundings. He would, he would cross the street without looking twice, comparatively, to to kids his age. He would jump in a pool without thinking about swimming like just. The risks that are that other kids inherently feel or know because we'd either warn them or they've learned it, he kind of he misses those for sure."<br>Canada, Caregiver (complex epilepsy), Participant 1 | No corresponding quantitative item. | No integration required. |
| | | Social vulnerability and relational risk | "She would even get in touch with anyone without limits. Another challenge when she gets older is that she can become a target for abusers."<br>Croatia, Caregiver (complex epilepsy), Participant 1 | No corresponding quantitative item. | No integration required. |
| | Burden of caregiving | Gaps in social support systems | "I don't have many organizations that can understand how I feel, or help me…"<br>South Korea, Caregiver (complex epilepsy), 1.<br><br>""When I have family parties, whether near or far, my attendance is limited, meaning that I stay for a very short time, since I cannot stay away from my daughter for long...As for her friend, she was limited to small relatives whom she met from time to time. Since she does not go to a specialized association. How I wish I could find an association that can frame such cases. on the one hand the child finds specialists who take care of him in the right way, and on the other hand the mother finds an outlet to renew her energy"<br>Tunisia, Caregiver (complex epilepsy), Participant 1<br><br>""I have felt the need to have is someone who can guide me, for me this is all new, and all you've got are associations... but there are no associations here in the Canaries, I am not a member of any epilepsy association... the neurologists don't work with any associations... they don't bring those families together, there's no meetings, no talks, nothing... I'm not in touch with families who have epilepsy, I'm scared, I mean I'm scared to compare cases, you know I would think this could be my son in the future, and so I feel like I also need some psychological help, which I can't find either.... And so I guess that in that sense I need an association"".<br>Spain, Caregiver (complex epilepsy), Participant 1<br><br>"some years ago I came across of a support group and the support group provide assistance to all kinds of patients not only epilepsy but any other condition, but that did not last long and their procedures are so tedious we could not proceed with that.""<br>The Gambia, Caregiver (complex epilepsy), Participant 3 | Respondents affected by complex epilepsy were significantly more likely than those with non-complex epilepsy to report that there needs to be inclusive social activities for caregivers and those they care for (54% vs. 41%; OR = 2.2, $p < 0.0001$). | Survey data showed that people affected by complex epilepsy were more likely to seek inclusive social activities for both caregivers and care recipients (54% vs. 41%; OR = 2.2, $p < 0.0001$). Qualitative accounts complement this by highlighting the social isolation, reduced access to peer networks, and limited community support faced by caregivers. These accounts reveal that inclusive activities are not only about recreation, but serve as a potential means to counteract exclusion, promote connection, and support overall wellbeing. Caregivers, particularly in complex epilepsy contexts, described significant withdrawal from friends and family, with calls for greater support from epilepsy organisations often linked to unmet psychological, emotional and practical needs. At the same time, some expressed hesitation to participate in peer groups due to discomfort or fear of confronting challenging realities, a theme present in both complex and non-complex experiences. |

| | | | | | |
|---|---|---|---|---|---|
| | | Hopes, fears, and uncertainty about the future | "An important topic to discuss is the employment of people with epilepsy and how to organize their lives after the passing of their parents or caregivers."<br>Bosnia & Herzegovina, Caregiver (non-complex), Participant 1<br><br>"I'm starting to see insecurities in him... he could basically have low self-esteem. Which could become depression. I worry a lot about when he sees there are certain things that he can't do, how he's going to fit in."<br>Spain, Caregiver (complex epilepsy), Participant 1 | No corresponding quantitative item. | No integration required. |
| | | **(Caregiver need)** Broader social and economic pressures | "we had to spend a lot of money. So, we reduced visiting healthcare facilities for seeking treatment"<br>India, Caregiver (complex epilepsy), Participant 1<br><br>"there should be special opportunities for carers to access credit facilities to enable them generate an income that's enough to care for their families."<br>Uganda, Caregiver (complex epilepsy), Participant 2<br><br>"Interviewer: what are [son's] needs to continue his treatment with the psychologist and the educational psychologist and the speech therapy?<br>Participant: Money basically, because everything depends on having private insurance. The State does not take care of that. That's the barrier, the barrier is purely and exclusively economic""<br>Argentina, Caregiver (complex epilepsy), Participant 4<br><br>"I have to take care of my child 24/7, which makes me lose the financial capacity as a mother. My child needs to be free from cramps to develop... so I prioritized it... That's the first thing I gave up my job. Naturally, mothers quit their jobs to care for their sick children...I hope there is a way to support such mothers to continue their economic activities while taking care of patients"<br>South Korea, Caregiver (complex epilepsy), Participant 1 | **Complementary:** Caregivers of people with complex epilepsy were significantly more likely than those caring for someone with non-complex epilepsy to report that they need money to cover the costs related to caring for someone (e.g., medicines, reduced work hours) (61% vs. 48%; OR = 2.2, $p < 0.0001$). | Quantitative findings show that caregivers of people with complex epilepsy face a greater financial burden compared to those caring for individuals with non-complex epilepsy (61% vs. 48%; OR = 2.2, $p < 0.0001$). This is complemented by qualitative accounts that vividly describe the economic strain, such as cutting back on healthcare visits, relying on private insurance, and losing income due to caregiving responsibilities. Although the qualitative data largely focuses on the impact of costs rather than direct expressions of financial need, the alignment is strengthened by participant reflections on the lack of state support and the need for economic opportunities. |

| | | | | | |
|---|---|---|---|---|---|
| | | **(Caregiver need)** Strain on family relationships and wellbeing | We were often away for a long time at (highly specialized Epilepsy Hospital) for several weeks. It was a difficult time for [PWE's] little sister. When was mom there, and when wasn't mom there? So, her little sister distanced herself from me for a while. So, it was a difficult time."<br>Denmark, CGC, Participant 4<br><br>"There has to be one of us always present... we do find challenges when the sister has to go to school and my husband goes to the market."<br>Uganda, Caregiver (complex epilepsy), Participant 2<br><br>" when we got the diagnosis, it was very difficult for me. I needed psychiatric support and medication, which I still take today. Later, when I went through my divorce—it has been a year now, after a 15-year marriage— that was also very difficult. So, I have both psychological and psychiatric support, with medication, and I'm doing okay. I am well cared for in this regard, but I cannot stop taking the medication or seeing professionals because otherwise, I wouldn't be well."<br>Brazil, Caregiver (complex epilepsy), Participant 5<br><br>"I suppose just that it's it's all connected and there's a chronic, chronic strain on your mental health. And yeah, just this, this undercurrent of of stress that you it either conquers you or you conquer it, but that that is something that needs to be dealt with...she picks her skin non-stop. You literally can't leave her. For more than two minutes by yourself... she's got impulsive behaviour and they have both have really high pain threshold and no concept of implications, consequences, No Fear. So it's it's a tough combination. We can't do anything on a whim. I can't leave the house by myself with both of them."<br>Australia, Caregiver (complex epilepsy), Participant 3 | No corresponding quantitative item. | No integration required. |
